# Maternal immune gene expression trajectories and dynamic genetic regulation throughout pregnancy

**DOI:** 10.64898/2026.09.22.26363644

**Authors:** Sophie Hoffman, Katie L Burnham, Emma Cook, Céleste E Cohen, Ulla Sovio, D. Stephen Charnock-Jones, Gordon C S Smith, Emma E Davenport

## Abstract

Pregnancy involves profound changes to maternal immunity throughout gestation to simultaneously tolerate the fetus and protect both mother and baby from infection. Dysregulation of this process can lead to pregnancy complications and adverse outcomes. However, how the maternal peripheral immune system adapts throughout an uncomplicated pregnancy remains poorly understood, limiting our ability to identify and interpret alterations associated with pregnancy complications.

Here, we longitudinally profiled maternal whole-blood transcriptomes of 1,305 nulliparous women at 12, 20, 28 and 36 weeks of gestation (n=3,776 samples) from the Pregnancy Outcome Prediction Study 2 (POPs2). Differential expression analysis revealed that the maternal immune system underwent its most pronounced remodeling in early pregnancy (between 12 and 20 weeks gestation). Co-expressed gene modules demonstrated immune-dominant programs were highly active in early stages, shifting towards increased cellular and metabolic activity in later pregnancy, reflecting maternal adaptation to increasing physiological demands.

Through *cis*-eQTL mapping we found that genetic regulatory effects increasingly diverged from the non-pregnant state as pregnancy advanced. Colocalising with GWAS revealed putative mechanisms underlying genetic predisposition to pregnancy complications, with 58% of these colocalizations detectable only in the context of pregnancy. Finally, we identified eQTL that exhibit significant interactions with gestational age, implicating dynamic regulation of innate immune and metabolic pathways. These findings show that pregnancy is not only a state of changing gene expression, but also a dynamic regulatory environment that reshapes the effects of genetic variation. By characterizing these adaptations across gestation, this work provides a reference for interpreting maternal immune variation and genetic risk across gestation.

## Introduction

Pregnancy represents a unique and dynamic immunological state in which the maternal immune system must balance the competing demands of tolerating the semi-allogeneic fetus while maintaining the capacity to respond to infection. This balance is achieved through tightly regulated changes in immune function across gestation^1^. Disruption of these processes has been implicated in a range of adverse pregnancy outcomes, including preeclampsia and preterm birth^2,3^. While progress has been made in characterising immune adaptation at the maternal-fetal interface^4–7^, less is understood about how these changes manifest in the maternal peripheral immune system.

Peripheral blood provides a tractable window into maternal immunological shifts in pregnancy and is an accessible and minimally invasive source of biomarkers. However, efforts to characterise immune gene expression in peripheral blood during pregnancy have been limited in cohort size and temporal coverage^8–10^, constraining the ability to capture both coordinated immune adaptation across gestation and its inter-individual variability. Recent large-scale studies of maternal physiology, including population-level analyses of clinical laboratory measurements across hundreds of thousands of pregnancies^11^, have highlighted the value of large-scale longitudinal datasets for understanding systemic changes during gestation. However, comparable comprehensive temporal profiling of maternal immune gene expression, which is necessary to resolve these changes at the molecular level and interpret deviations associated with complications, are lacking.

Beyond average changes in gene expression, understanding how inter-individual variability influences maternal immunity in pregnancy is critical. Periods of increased or reduced transcriptomic variability may reflect temporal shifts in regulatory constraint, yet such dynamics have not been systematically explored in pregnancy. Moreover, the extent to which genetic variation influences immune gene expression during pregnancy and how this regulation may change over time, remains poorly defined. This gap limits the interpretation of genetic associations with pregnancy-related traits.

Genome-wide association studies have identified numerous loci associated with pregnancy outcomes^12–15^. These genetic associations can act through gene expression via expression quantitative trait loci (eQTL), which are often context-specific^16^. Existing eQTL resources derived from non-pregnant individuals are unlikely to capture these context-dependent effects, limiting their utility for interpreting mechanisms underlying genetic risk of pregnancy outcomes.

We address these gaps by generating and analysing a large-scale longitudinal dataset of whole-blood transcriptomes and genotypes from pregnant women sampled across gestation. By modelling gene expression, its variance, and co-expression module trajectories over time, we characterize coordinated temporal changes in immune functions and identify periods of altered regulatory constraint. We map expression quantitative trait loci and, by integrating them with genetic association studies, provide novel insights into putative molecular mechanisms underlying pregnancy outcomes.

Together, this work provides a comprehensive reference of maternal peripheral immune variation across pregnancy and establishes a framework for integrating longitudinal transcriptomic and genetic data in a natural human perturbation. These insights advance our understanding of the maternal immune trajectory in pregnancy and provide a foundation for the development of clinically actionable biomarkers and therapeutic strategies to improve maternal and fetal health.

## Results

### The POPs2 RNA-seq cohort

We recruited nulliparous pregnant women through the Pregnancy Outcome Prediction Study 2 (POPs2) and serially characterised the maternal peripheral immune system using whole blood bulk RNA-seq on 1,305 individuals at up to 4 time-points (12, 20, 28, 36 weeks gestational age [wkGA], **Table 1**, **Figure 1A**, **Supplementary Figure 1**). The majority of participants had uncomplicated pregnancies, with 43 cases of gestational diabetes (4.3%), 61 cases of pre-eclampsia (6.4%); 18 cases of spontaneous preterm delivery (2.0%), and 68 cases where the infant was small for their gestational age (7.1%, **Table 2**). This cohort therefore enabled us to establish typical gene expression patterns throughout pregnancy.

**Figure 1:**
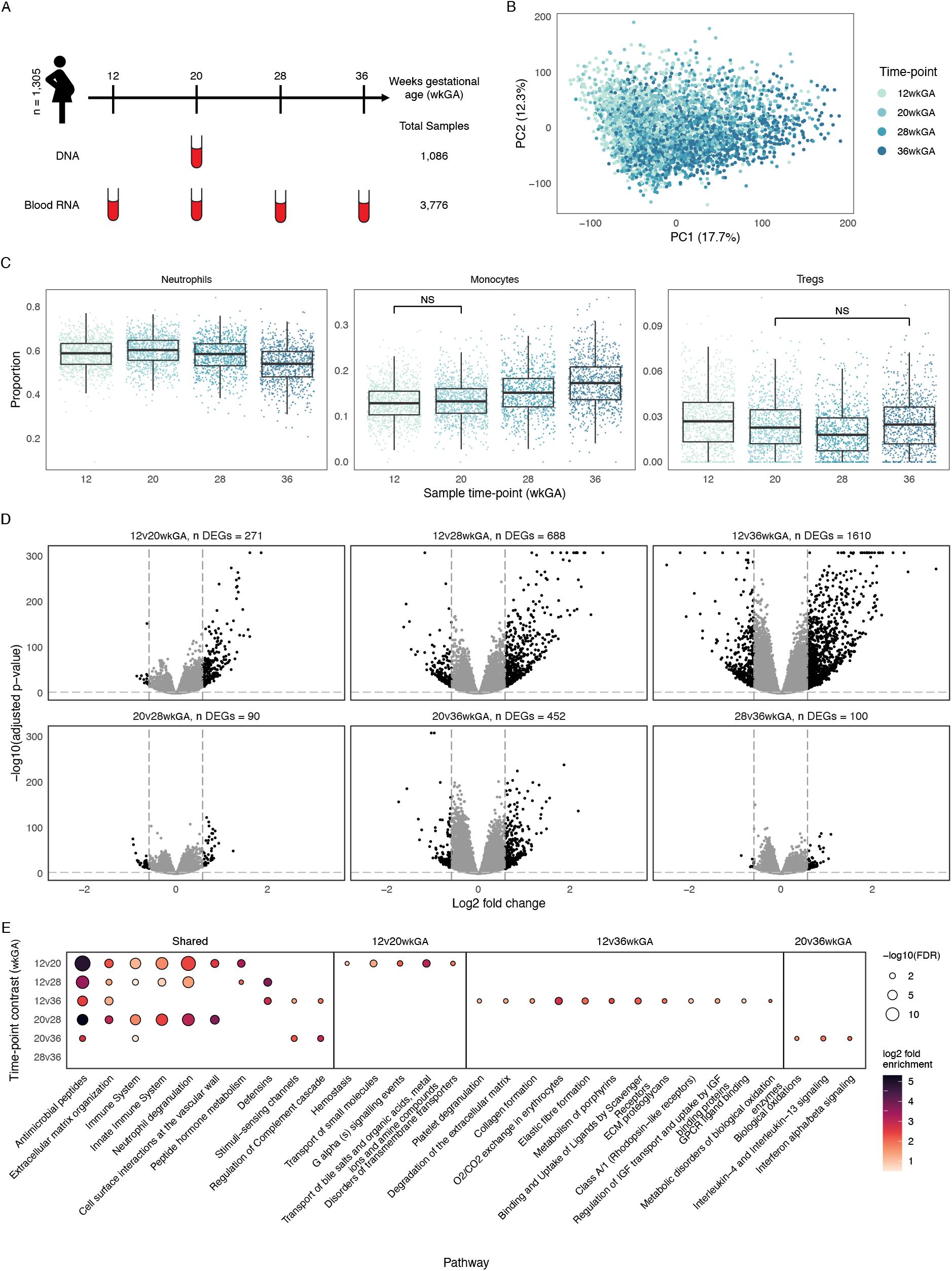
Changes in gene expression and cell proportions over time. A) Study design. B) PCA of full RNA-seq cohort colored by sample time-point. C) Boxplots of CIBERSORTx imputed cell proportions at each pregnancy time-point in Neutrophils, Monocytes and Regulatory T cells. Cell proportions differ significantly (p < 0.05) between time-points except where marked as “NS”. D) Volcano plots of log2 fold change of gene expression vs. -log10 adjusted p-values for all pregnancy time-point pairs. Differentially expressed genes (DEGs) are shown in black and are defined as having fold change greater than 1.5 and adjusted p-value less than 0.01. Each plot is annotated with the number of DEGs identified at that time-point contrast. E) XGR pathway enrichment (Reactome ontology) of DEGs across time-point contrasts. All pathways shown are significantly enriched in at least one time-point contrast, and only significantly enriched pathways are represented as dots (FDR < 0.05). Dot size represents -log10(FDR) and color represents log2-fold enrichment. Pathways grouped by sharing across time-point contrasts.

**Table 1.**
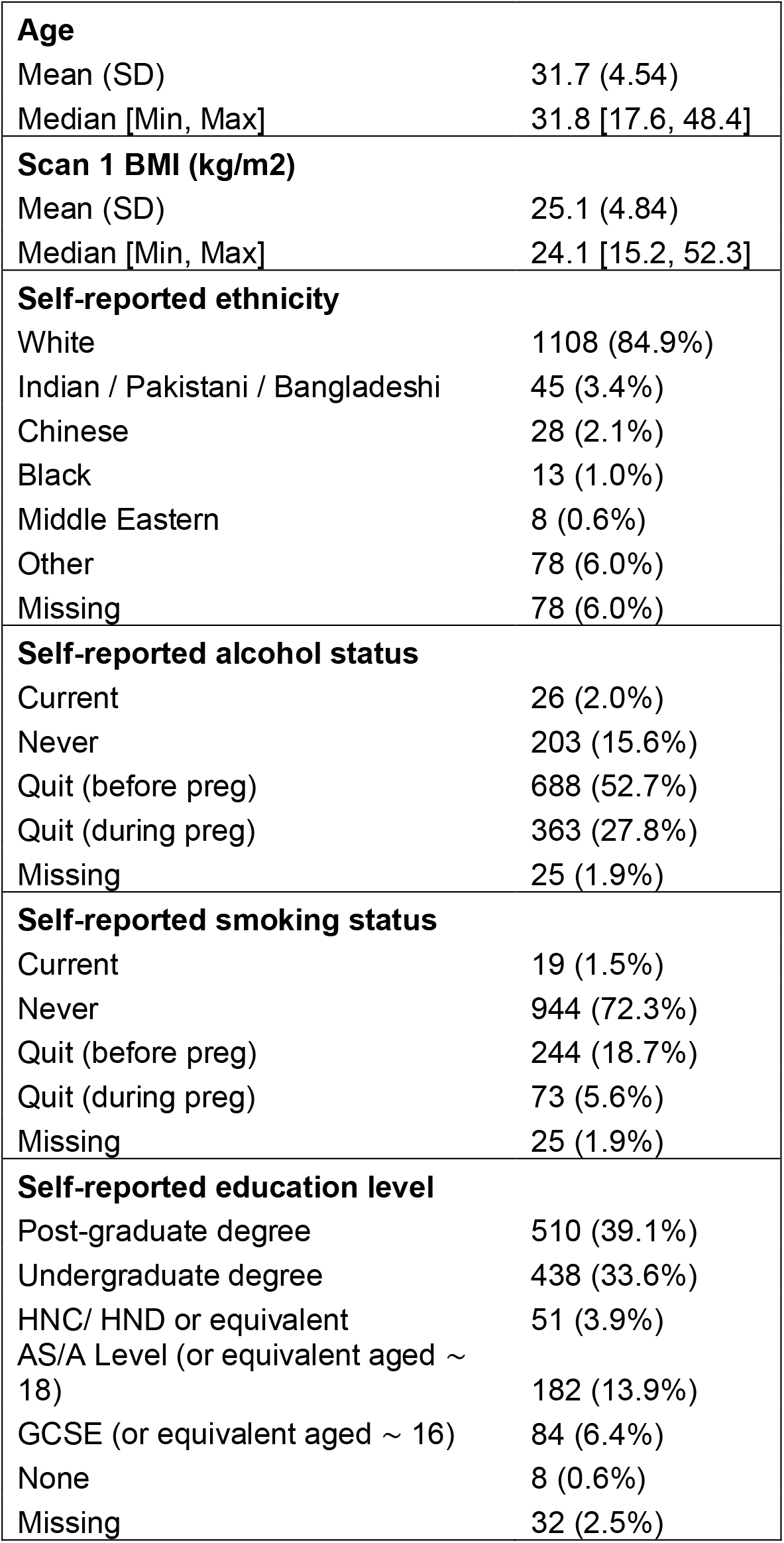
Characteristics of the POPs2 RNA-seq cohort.

**Table 2:** Numbers and rates of pregnancy outcomes in the POPs2 RNA-seq cohort.

|  |  |
| --- | --- |
| **Diabetes** |  |
| Gestational diabetes mellitus | 41 (3.1%) |
| Pre-existing | 6 (0.5%) |
| None | 907 (69.5%) |
| Missing | 351 (26.9%) |
| **Pre-eclampsia (PE)** |  |
| PE, severe | 42 (3.2%) |
| PE, not severe | 19 (1.5%) |
| No PE | 893 (68.4%) |
| Missing | 351 (26.9%) |
| **Spontaneous preterm birth** |  |
| Yes | 18 (1.4%) |
| No | 935 (71.6%) |
| Missing | 352 (27.0%) |
| **Induction of labor** |  |
| Yes | 385 (29.5%) |
| No | 565 (43.3%) |
| Missing | 355 (27.2%) |
| **Any neonatal morbidity** |  |
| Yes | 88 (6.7%) |
| No | 866 (66.4%) |
| Missing | 351 (26.9%) |
| **Gestational age at birth, weeks** |  |
| Preterm: 24-<33 | 6 (0.5%) |
| Preterm: 33-<37 | 31 (2.4%) |
| Term >= 37 | 916 (70.2%) |
| Missing | 352 (27.0%) |
| **Birth weight centile** |  |
| Severe small for gestational age (SGA, < 3rd) | 16 (1.2%) |
| SGA (< 10th) | 52 (4.0%) |
| Not SGA | 885 (67.8%) |
| Missing | 352 (27.0%) |
| **Mode of delivery** |  |
| Spontaneous vaginal | 450 (34.5%) |
| Assisted vaginal | 201 (15.4%) |
| Intrapartum cesarean | 148 (11.3%) |
| Pre-labor cesarean | 144 (11.0%) |
| Missing | 362 (27.7%) |

### Gene expression profiles demonstrate controlled temporal regulation of gene expression in pregnancy

#### Gene expression and cell proportions change over time in pregnancy

We found that global blood gene expression changes occur throughout the course of pregnancy, as indicated by a clear gradient on principal components from 12 to 36wkGA. (**Figure 1B**). We also observed differences in 18 imputed immune cell proportions between the time-points, with innate and adaptive immune cells showing distinct patterns of change in abundance throughout pregnancy (**Figure 1C**, **Supplementary Figures 2-4**, **Supplementary Table 1**). We validated the imputed cell proportion trajectories against measured leukocyte fractions from Bar et al. (2025)^11^. Although agreement in weekly mean cell proportions was modest (except for monocytes), the major leukocyte populations exhibited similar temporal trajectories across datasets, with differences in timing likely due to temporal alignment by conception (POPs2) versus delivery (Bar et al., **Supplementary Figure 5**).

To identify the specific genes modulated during pregnancy, we performed differential expression analysis between all time-point pairs. This identified 1,740 unique differentially expressed genes (DEGs), with differential expression occurring in all time-point contrasts (**Figure 1D**, **Supplementary Table 2**). The 12 and 36 week time-points were the most transcriptomically different, with 1,610 genes differentially expressed, whereas 20 and 28wkGA were the most similar time-points, reflected by the fewest DEGs (n = 90, **Supplementary Figure 6**). On comparing adjacent time-points we observed that the maternal immune system changes most between 12 and 20wkGA (n DEGs = 271), indicating a large transcriptomic shift in early pregnancy. Conversely, differential expression analysis across pregnancy outcome groups demonstrated limited signal (**Supplementary Figure 7**, **Supplementary Table 3**). Differences in cell composition are known to influence bulk transcriptomic profiles. However, differential expression results were highly consistent between models including vs. excluding imputed cell proportions, with strong correlations of log2-fold change values across all time-point contrasts (minimum Pearson’s r=0.91), indicating cellular composition was not a major driver of the observed differential expression across time-points (**Supplementary Figure 8**). Pathway enrichment analysis of DEGs revealed changes in both shared and unique immune-related pathways across five time-point contrasts (**Figure 1E**). For example, antimicrobial peptide-related gene expression changes between five time-point pairs, and extracellular matrix organization and immune system-related gene expression changes between four time-point pairs.

We compared our findings to the largest publicly available pregnancy transcriptomics study (Gomez-Lopez et al. 2019^9^, Affymetrix [HTA 2.0] microarray, n samples used = 174), applying the same analytical methods and significance thresholds and considering only the 12,814 genes detected in both cohorts. Out of 48 genes differentially expressed at any time-point in the Gomez-Lopez et al. (2019)^9^ dataset, we re-capitulate 89.58% (**Supplementary Figure 9**). The additional differentially expressed genes we observed in POPs2 were likely due to both increased power and greater sensitivity of RNA-seq compared to microarray.

#### Gene co-expression networks capture distinct biological processes with different patterns of change over time

To investigate temporally stable gene co-expression, we constructed consensus gene co-expression modules using WGCNA^17^ by restricting module membership to genes that were consistently co-expressed across all four time-points. As expected with this stringent approach, the 15 identified modules contained relatively few genes, with 2,403 of the 18,826 expressed genes assigned to modules. Module sizes ranged from 55 to 330 genes. (**Supplementary Figure 10**, **Supplementary Table 4**). Module eigengenes, defined as the first principal component of each module, summarise module activity. As all module eigengenes were positively correlated with the expression of their member genes, increases in eigengene values reflect coordinated upregulation of genes within the module, and decreases reflect coordinated downregulation. We fitted linear mixed-effects models of eigengene values vs time to characterise module trajectories and found various patterns of change in module activity over time (**Figure 2**). Module eigengenes can have different trajectories across clinical subgroups. On testing for differences by complication status, we found module trajectories showed only nominally significant differences across clinical subgroups (**Supplementary Figure 12**).

**Figure 2:**
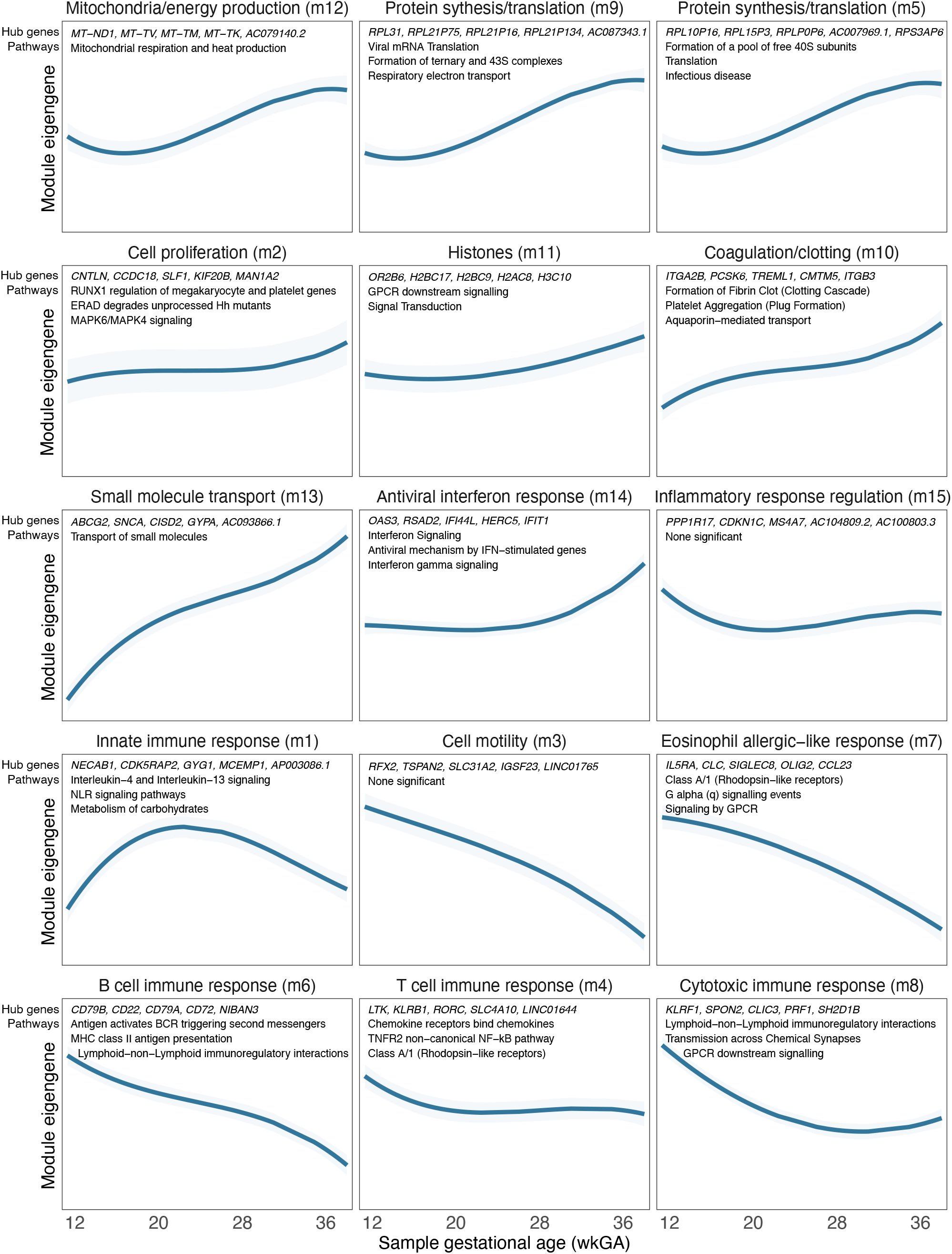
Temporal changes in gene co-expression networks. Temporal module eigengene profiles where curves represent predicted eigengene values over time and shaded areas represent 95% confidence interval of predicted values. Each module is annotated with its top 5 hub genes (most interconnected as measured by kWithin metric) and top 3 enriched Reactome pathways (measured by fold enrichment). Reactome pathways with long names have been abbreviated in the plot title, full pathway names are available in Supplementary Table 5. Each module has been named to summarize module annotations including top 10 hub genes, all enriched pathways, all enriched regulons, and correlated and enriched cell types. Full annotation information for each module can be found in Supplementary Table 5. Modules ordered by similarity of eigengene values as displayed in Supplementary Figure 11.

After annotating modules with biological functions (**Supplementary Table 5**), we compared module eigengenes to related clinical measures from a large-scale dataset of blood laboratory tests from pregnant women described in Bar et al. (2025)^11^. Associations between weekly mean module eigengene values and the corresponding weekly mean lab test values were assessed using Pearson correlation. For example, Module 10 is enriched for coagulation-related genes and is also strongly correlated with multiple coagulation-related lab tests (**Supplementary Figure 13A**), supporting a functional increase in coagulation-related gene expression over time in pregnancy. Module 13, enriched for small molecule transport-related genes, is strongly correlated with various red blood cell-related lab tests, reflecting the importance of small molecule transport processes in red blood cell physiology (**Supplementary Figure 13B**).

#### Changes in gene expression variance over time in pregnancy indicate dynamic gene regulation

We further investigated the extent to which gene expression variance changed over time (heteroskedasticity), hypothesising that genes critical at different stages of gestation would display tightly controlled expression only at specific time-points. We found that 2,296 unique genes displayed heteroskedasticity and had at least one significant change in expression variance between time-points (**Figure 3A**, **Supplementary Table 6**). The most changes in variance occurred between 12 and 36wkGA (n genes = 1,577), and the least between 12 and 20wkGA (n genes = 31). The majority of significant genes in the 12 to 28wkGA contrast (86.19% of significant genes) showed a reduction in variance, whereas from 20 to 36wkGA a greater proportion showed increased variance (80.11%). This pattern of reduced gene variance in mid-pregnancy could suggest tighter gene regulation at this stage.

**Figure 3:**
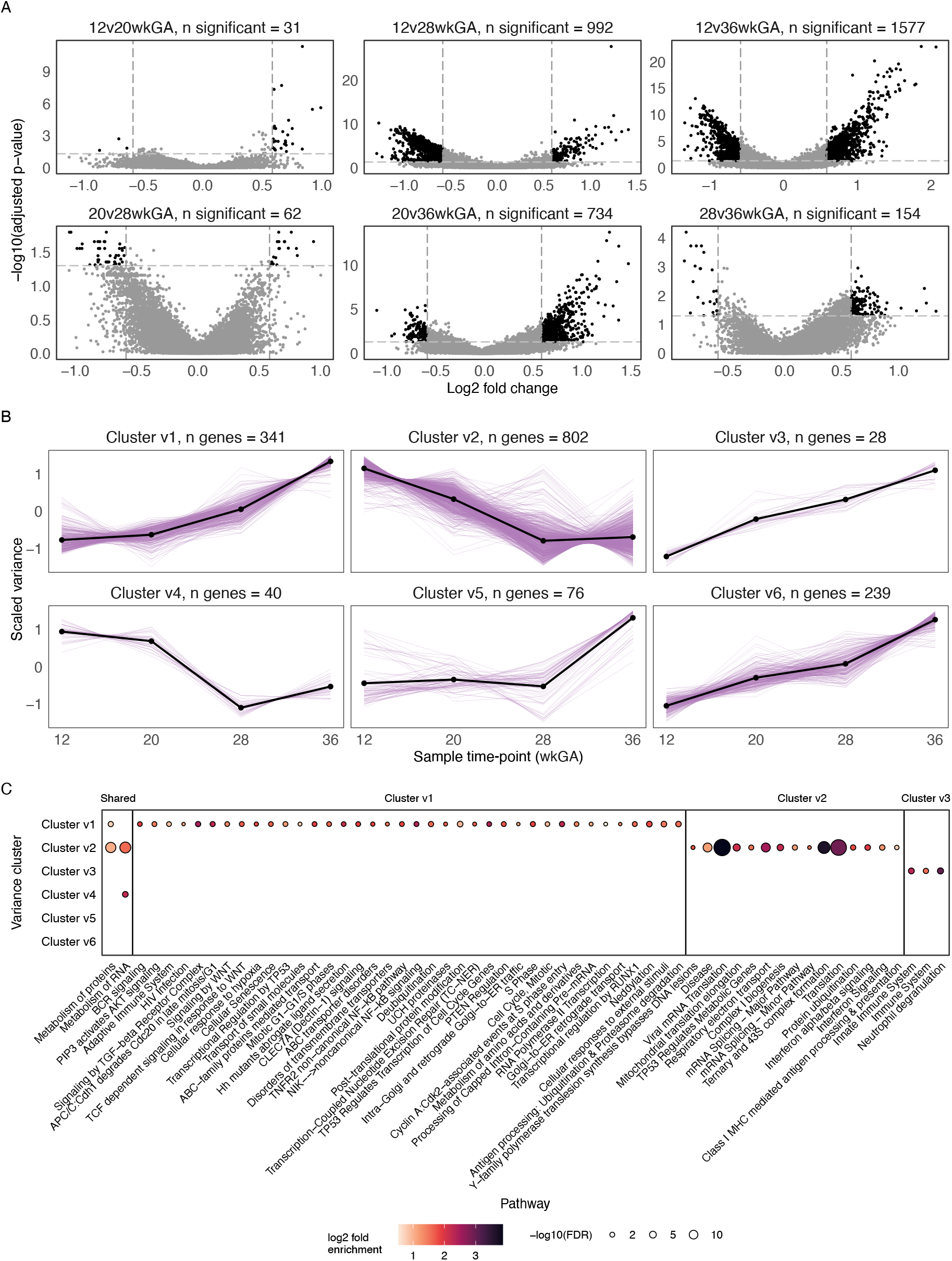
Temporal changes in gene expression variance. A) Volcano plots of log2 fold change in gene expression variance vs. -log10 adjusted p-values for all pregnancy time-point pairs. Differentially variable genes (DVGs) between time-points are shown in black, defined as having a fold change greater than 1.5 and adjusted p-value less than 0.05. Genes with positive fold change increase in variance from the earlier to the later time-point. Genes with negative fold change decrease in variance from the earlier to later time-point. Each plot is annotated with the number of DVGs identified at that time-point contrast. B) Scaled mean variance per gene at each time-point for genes in each cluster. Each plot is annotated with the number of genes in the cluster. Mean scaled variance across all genes in each cluster are represented by the black points and lines. C) XGR pathway enrichment (Reactome ontology) of differentially variable genes across time-point contrasts. All pathways shown are significantly enriched in at least one time-point contrast, and only significantly enriched pathways are represented as dots (FDR < 0.05). Dot size represents -log10(FDR) and color represents log2 fold enrichment.

Groups of genes that display similar temporal variability patterns could represent co-regulated gene programs, where the level of variance may reflect the degree of regulatory constraint within these programs. To investigate this, we defined six clusters of heteroskedastic genes with different patterns of variance over time (**Figure 3B**, **Supplementary Table 7**) and identified enriched Reactome pathways in each cluster (**Figure 3C**).

Both cluster v1 and v2 are enriched for pathways that reflect immune activation but differ in emphasis and variance pattern over time. Genes in cluster v1 have no change in variance from 12 to 20wkGA but increase in variance from 20 to 36wkGA. This cluster is enriched for 39 pathways related to cell cycle progression, cell signaling, cellular stress responses, metabolism and immune activation, collectively suggesting a role in immune cell signaling, activation and proliferation. Cluster v2 genes decrease in variance from 12 to 28wkGA and then show little change from 28 to 36wkGA. This cluster is enriched for 17 pathways related to translation, energy metabolism and interferon-mediated immune signaling, consistent with a role in immune cell activation and proliferation. Taken together, these patterns suggest that immune cell activation has less variable proliferation and signaling response in early pregnancy, whereas interferon-driven translational and metabolism focused activation become less variable at the end of pregnancy, highlighting possible temporal specificity of regulation of immune cell activation mechanisms.

### Genetic regulation of gene expression in pregnancy

Having identified substantial inter-individual variation in gene expression between individuals, we then aimed to understand the relationship between genetic variation and gene expression in the context of pregnancy. We mapped *cis*-eQTL in the 1,083 individuals with both gene expression and genotyping data using a linear mixed model, and fine-mapped 25,779 eQTL signals across 14,268 eGenes across the autosomes and X chromosome.

#### Pregnancy eQTL nominate mechanisms mediating genetic drivers of pregnancy traits

To gain insight into the molecular mechanisms through which genetic variants influence risk of pregnancy complications, we performed statistical colocalisation analyses between our pregnancy eQTL and publicly available GWAS results. We identified eQTL colocalizing with risk loci for gestational diabetes^12^ (n=43 locus-eGene pairs), pre-eclampsia^13,14^ (n=24), gestational duration^15^ (n=8), and preterm birth^15^ (n=2) (**Supplementary Table 8**), with the number of colocalizations correlating with the number of GWAS loci for each trait (Pearson’s r=0.97, **Supplementary Figure 14**).

Many of the colocalising eGenes were biologically plausible potential mediators of the phenotypic outcomes. In gestational diabetes, 36 of the 40 unique colocalising eGenes had previously established links to diabetes, insulin signalling, pancreatic function, or obesity. We further observed colocalization between an eQTL for *OPRL1* and a gestational duration GWAS locus on chromosome 20 (PPH4 = 0.99, **Supplementary Figure 15**), consistent with existing evidence that OPRL1 signaling regulates uterine contractility^18^ and suggesting a potential maternal mechanism influencing labour timing. We also identified a colocalization between a *VEGFA* eQTL and a pre-eclampsia GWAS locus on chromosome 6 (PPH4 = 0.99, **Supplementary Figure 16**). VEGFA is a key regulator of angiogenesis and endothelial function, and its expression is responsive to hypoxic signalling, which is implicated in pre-eclampsia^19^. Finally, an eQTL for *MSMO1* colocalized with a preterm birth locus on chromosome 4 (PPH4 = 0.98, **Supplementary Figure 17**), connecting the role of MSMO1 in cholesterol biosynthesis with prior evidence that disrupted maternal cholesterol levels are linked to risk of preterm birth^20^.

Given that regulation of gene expression can be context-specific, we hypothesised that the consequences of a subset of genetic risk loci would only be revealed using eQTL mapped during pregnancy. Indeed, colocalization using published summary statistics from the INTERVAL^21^ blood donor cohort demonstrated that pregnancy transcriptomic context provides substantial additional insight into related GWAS mechanisms. Of the 77 unique colocalizations identified with our pregnancy eQTL, 58% (n = 45) were not detected with INTERVAL eQTL despite greater statistical power, therefore revealing candidate mechanisms that are unique to the context of pregnancy (**Figure 4A**, **Supplementary Table 8**). For example, an eQTL for *NOTCH2*, which has previously been reported to have altered expression levels in type 2 diabetes^22^, colocalizes exclusively in POPs2 with a gestational diabetes GWAS hit on chromosome 1 (PPH4 = 0.96, **Figure 4B**). The remaining 42% of colocalizations with POPs2 (n=32) also colocalized with INTERVAL (**Figure 4A**), indicating that some GWAS loci are mediated by regulatory mechanisms active across adult life, whereas others are only revealed under pregnancy-specific physiological conditions.

**Figure 4:**
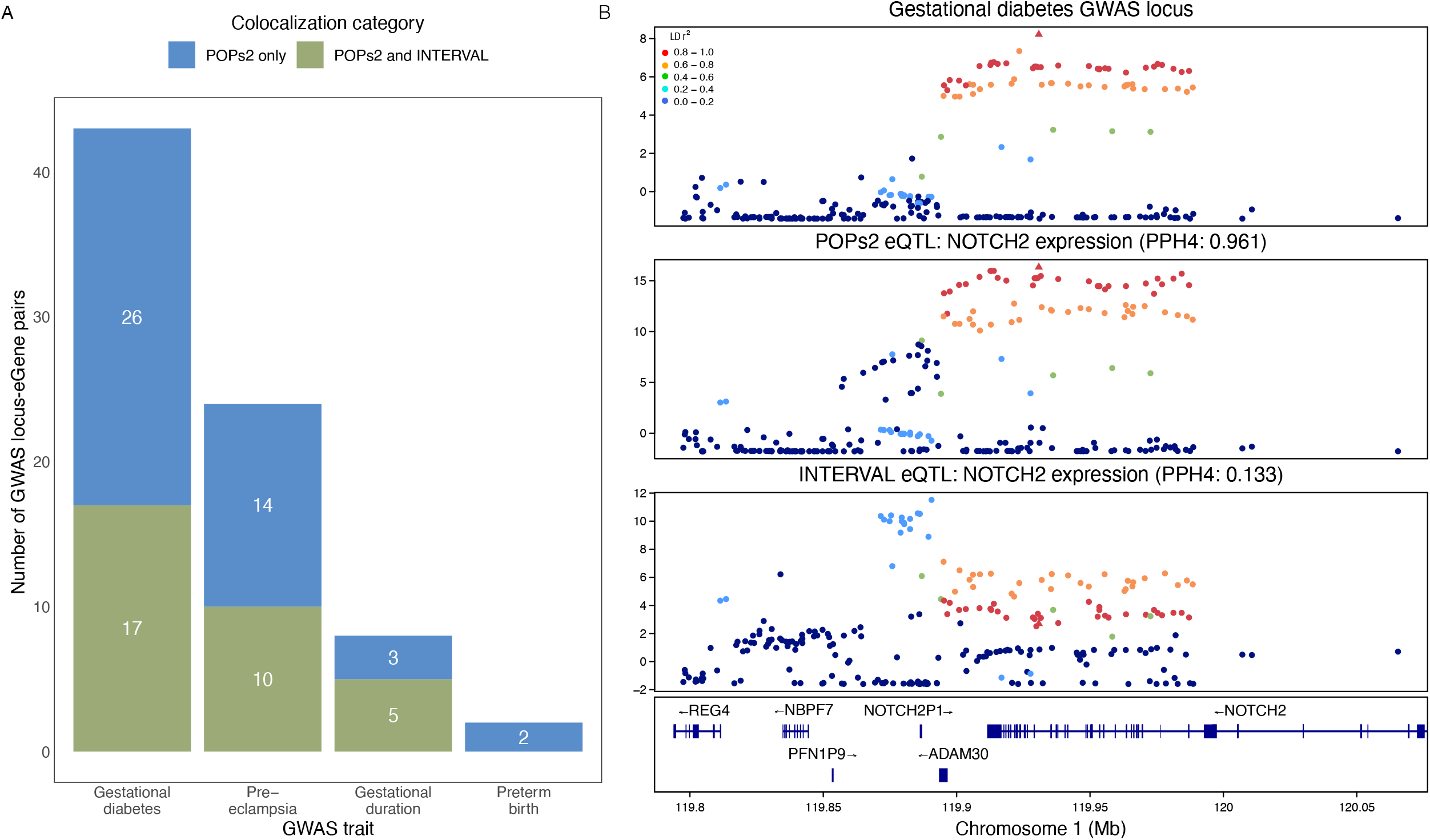
Colocalisation of eQTL with GWAS. A) Stacked bar chart summarising the number of significant colocalisations (PPH4 > 0.8) with each GWAS trait, coloured by eQTL dataset(s) where colocalisation was detected. B) Regional association plots showing chromosome position vs. log approximate Bayes factor (lABF) from SuSiE. The lABF quantifies the strength of evidence that a variant is associated with the trait. The triangular point represents the joint lead SNP between GWAS and pregnancy eQTL signals and points are colored by LD with that SNP. PPH4 of colocalization between eQTL and GWAS signal noted in subtitle. Gestational diabetes GWAS locus (top) and NOTCH2 eQTL in POPs2 (middle) and INTERVAL blood donor cohort (bottom).

#### eQTL mapped in pregnancy exhibit context-specific effects

In order to understand why some colocalisations were exclusive to the POPs2 dataset, we comprehensively evaluated the pregnancy-specificity of the eQTL we detected using mashR. For this comparison, we re-mapped eQTL in just females of reproductive age from INTERVAL (age <45, n=445) to limit differences driven by age and sex. We subsetted the POPs2 cohort to individuals with RNA-seq available at all four time-points (n=448) and mapped eQTL per time-point to enable a time-point specific, power-matched comparison to INTERVAL using mashr (**Figure 5A**). eQTL significant at each time-point in pregnancy were categorized as shared with non-pregnant signals (up to 86.4%) or context-dependent (up to 19.1%), with 11.5%-16.4% of these being pregnancy-magnified (**Figure 5B**). Eight colocalizations involving eGenes with pregnancy-magnified signals were exclusive to pregnancy (*GRB10, KIAA1191, NOTCH2, NPC1, TBC1D22B, NRBF2, TSHZ3,* and *MSMO1*). An additional five loci (*CDC123*, *EHBP1L1*, *RUVBL2*, *JMJD1C*, and *PECAM1*) colocalized with both pregnancy and INTERVAL eQTL but involved eGenes with pregnancy-magnified signals, indicating that pregnancy-specific modulation of otherwise constitutive regulatory mechanisms may be critical for interpreting these GWAS associations.

**Figure 5:**
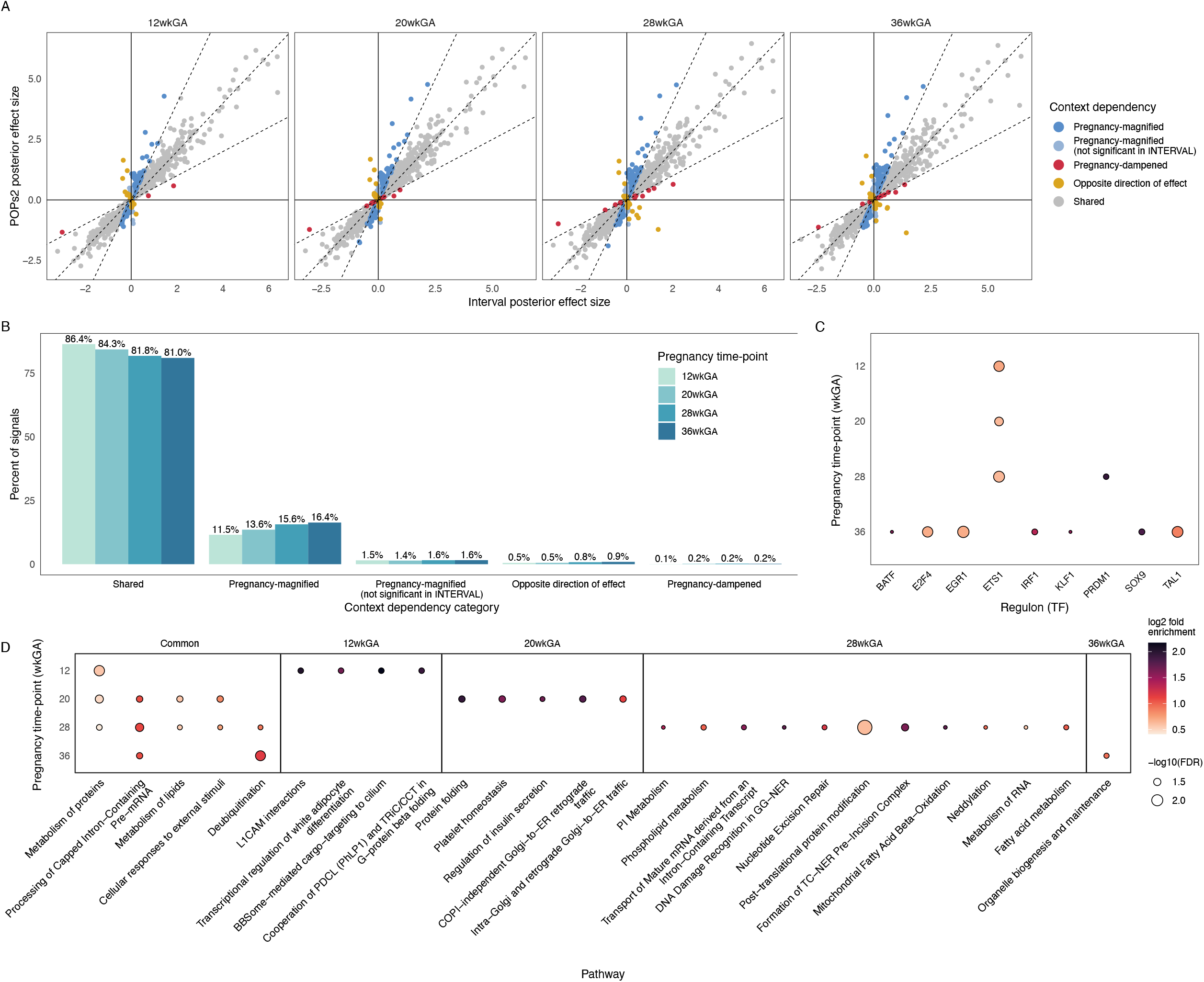
Context-specific eQTL in pregnancy. INTERVAL eQTL are compared to pregnancy eQTL mapped separately at each time-point. A) Scatterplot of posterior effect size estimates by mashr for significant lead SNP-gene pairs from POPs2 also tested in INTERVAL. eQTL significant in pregnancy are categorised as “shared” if mashr effect size is in the same direction as and within a factor of 0.5 of the INTERVAL effect size, as denoted by the dashed lines. Those with bigger effects in the same direction in POPs2 or only significant in POPs2 are classed “pregnancy-magnified” whereas larger effects in INTERVAL are “pregnancy-dampened”. Effects are “opposite direction of effect” if significant in both datasets but with opposite directions of effect. B) Barchart quantifying eQTL context-specificity at each time-point. C) Dotplot of enriched regulons among pregnancy-magnified eGenes across time-points. Dot size represents -log10(FDR) and color represents log2 fold enrichment. Only regulons with FDR < 0.05 shown. D) XGR pathway enrichment (Reactome ontology) of pregnancy-magnified eGenes at each time-point. All pathways shown are significantly enriched among pregnancy-magnified eGenes for at least one time-point, and only significantly enriched pathways are represented as dots (FDR < 0.05). Dot size represents -log10(FDR) and color represents log2 fold enrichment. Pathways grouped by sharing across time-points.

Beyond contextualising the colocalisation results, the mashR analysis provides insight into how genetic regulation itself evolves across gestation. Quantification of eQTL context-specificity revealed a progressive regulatory shift whereby the proportion of shared signals decreases over time, while the proportion of pregnancy-magnified signals increases (**Figure 5B**). This shift indicates that the landscape of genetic regulation of gene expression is most similar to the non-pregnant state in early pregnancy and becomes increasingly different by 36wkGA.

Pregnancy-magnified eGenes were enriched for the target genes of specific transcription factors (DoRothEA^23^ regulons), suggesting that the expression of these eGenes could be modulated by context-specific regulators (**Figure 5C**). For example, the ETS1 regulon, which regulates cytokine and chemokine production and contributes to lymphoid and vascular processes^24,25^, was enriched at multiple time-points (12, 20, and 28wkGA), suggesting a more sustained role in maternal immune and vascular regulation across early and mid-pregnancy. Pregnancy-magnified eGenes were also enriched for Reactome pathways (**Figure 4D**). Pathways shared across time-points include transcription and protein and lipid metabolism. Interestingly, regulation of insulin secretion is enriched among pregnancy-magnified eGenes at 20wkGA. Changes in insulin sensitivity throughout pregnancy are mediated by placental hormones. These insulin-related eQTL signals emerge just prior to routine GDM screening at 24–28wkGA, highlighting that genetic regulation of insulin-related pathways is most pronounced prior to the detection of pregnancy-induced insulin resistance. This finding exemplifies the type of dynamic, temporally restricted regulatory effects that can be uncovered through longitudinal eQTL mapping.

#### eQTL display dynamic effects over time in pregnancy

To comprehensively characterise eQTL effects that differ across time-points, we tested 23,859 conditionally independent eQTL signals for interactions with pregnancy time-point. We identified 1,193 significant interactions in 1,050 unique genes. Time interaction eGenes were enriched for pathways related to innate immune function, degranulation, and metabolic processes (**Supplementary Figure 18A**), indicating that these biological processes are under dynamic genetic regulation in pregnancy. Time interaction eGenes were also overrepresented among androgen receptor (FDR=0.018) and TAL1 targets (FDR=0.015, **Supplementary Figure 18B**), implicating these transcription factors (TFs) as potential upstream regulators of dynamic temporal eQTL effects. 59.5% (n = 710) of these time interaction eQTL also have interactions with neutrophil and/or monocyte proportions, indicating that shifts in blood cell composition may underlie the observed temporal genetic effects (**Supplementary Figure 19A**). To more directly assess this relationship, we tested whether time interactions persisted after accounting for cell proportion interactions and found that up to 60% of eQTL with both time and cell proportion interactions showed a time effect independent of cell composition (**Supplementary Figure 19B-C**). Together with the remaining 40.5% (n = 483) of time interactions that do not overlap with cell proportion interactions (**Supplementary Figure 19A**), these results suggest that cell composition contributes to temporal eQTL effects, but does not fully account for them, with the remaining signal likely reflecting either changes in smaller cell populations or additional regulatory mechanisms.

To further uncover mechanisms underlying dynamic temporal eQTL effects and capture directionality, we grouped these eQTL into five clusters based on standardized effect sizes across gestation, representing distinct temporal patterns of genetic regulation (**Figure 6A-B**). These clusters were characterised using pathway and regulon enrichment analyses to provide insight into the biological processes underlying each temporal pattern (**Figure 6C**, **Supplementary Figure 20**). For example, cluster t4 comprised eQTL with decreasing effect sizes across gestation and was enriched for immune signaling pathways. These eGenes were overrepresented among SMAD3 (FDR=2.2×10^-2^) and STAT1 (FDR=6.98×10^-5^) targets (**Supplementary Figure 20**), consistent with decreases in inferred activity of SMAD3 and STAT1 regulons across gestation (**Supplementary Figure 21**, **Supplementary Table 9**), with a small subset of these eQTL overlapping corresponding TF binding sites (**Supplementary Table 10**). This evidence supports a model in which reduction of SMAD3 and STAT1 activity attenuates cluster t4 eQTL effects over time. However, across clusters, we did not observe consistent evidence linking transcription factor binding disruption at eSNPs to changes in expression of downstream target genes, suggesting that temporal eQTL effects arise from combinations of regulatory mechanisms and cell type-specific effects that cannot be fully resolved in bulk data. These results show that changes in the maternal blood environment across gestation reshape genetic regulation of immune processes in distinct temporal patterns and provide a framework for linking dynamic genetic effects to regulatory mechanisms that change over time.

**Figure 6:**
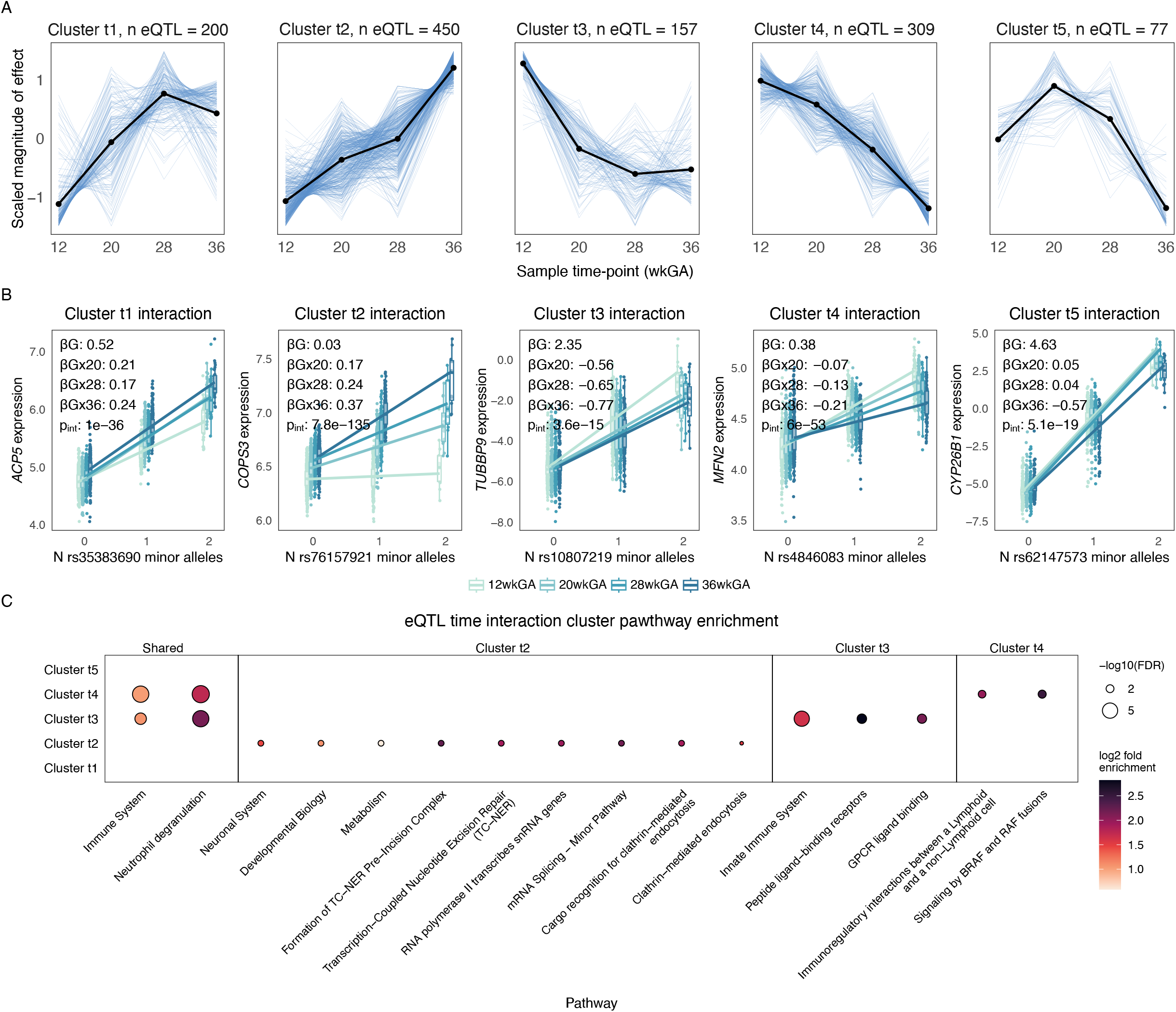
eQTL interactions with time. Summary of all eQTL time interaction clusters. For each cluster A) scaled β values at each time-point are shown for all eQTL in cluster, with the mean trajectory across eQTLs indicated by black points and lines and annotated by number of eQTL in cluster. B) an example eQTL from each cluster shown in plots of n minor alleles vs. partial residuals (after regressing out covariates included in the eQTL model) for the interaction model colored by time-point. βG denotes the genotype effect size at 12 weeks, βGX20, βGX28, and βGX36 denote interaction effect size differences at 20, 28, and 36wkGA, respectively, and pint indicates the p-value for the genotype-time interaction. C) XGR pathway enrichment (Reactome ontology) of eGenes in each cluster. All pathways shown are significantly enriched among eGenes in at least one cluster, and only significantly enriched pathways are represented as dots (FDR < 0.05). Dot size represents -log10(FDR) and color represents log2 fold enrichment. Pathways grouped by sharing across clusters.

## Discussion

Pregnancy involves dynamic, trimester-specific changes in maternal blood gene expression reflecting tightly regulated immunological remodelling across gestation. We present a large-scale longitudinal transcriptomic resource comprising 3,776 RNA-seq samples from 1,083 genotyped individuals at four time-points to define the temporal dynamics of maternal immune gene expression and genetic regulation in pregnancy.

### Gene expression

CIBERSORTx inferred cell proportions reveal dynamic, stage-specific immune adaptations across gestation, consistent with a coordinated shift in maternal immune function. Early pregnancy is characterised by elevated innate and humoral responses, including peaks in neutrophils, NK cells, mast cells and B cell subsets, potentially reflecting inflammatory and antigen-recognition processes associated with implantation. Late pregnancy demonstrates an increase in monocytes, macrophages, dendritic cells and T cells, indicative of enhanced immune surveillance and maintenance of homeostasis under a state of high physiological demand. These findings recapitulate and extend prior observations of patterns of change in immune cell composition^8,26–28^, while highlighting finer temporal dynamics such as a mid-gestational reduction in regulatory T cells underscoring the value of high-resolution longitudinal sampling. However, the compositional nature of inferred proportions limits interpretation of absolute cell population changes.

We found 1,740 genes enriched for immune-related pathways that were differentially expressed between pairs of pregnancy time-points. On comparing adjacent time-points we observed that the maternal immune system changes most between 12 and 20wkGA indicating a large transcriptional shift in early pregnancy, and least in the middle of pregnancy between 20 and 28wkGA. These patterns indicate that early pregnancy represents a critical window of immune remodelling, while mid-gestation is comparatively transcriptiomically consistent, a finding that can inform more efficient sampling strategies in future longitudinal pregnancy studies. Notably, changes in gene expression were not primarily driven by shifts in inferred cell proportions, suggesting substantial reprogramming of cell states in pregnancy, and motivating future single-cell approaches to resolve these dynamics at higher resolution.

Gene modules revealed coordinated immune and core cellular programmes underlying gestational adaptation, including established patterns such as increased coagulation. Early pregnancy was characterised by heightened immune activity, spanning inflammatory regulation, and both innate and adaptive responses, whereas late pregnancy showed greater engagement of processes including energy production and protein synthesis. Notably, innate immune activity peaked in mid pregnancy, likely reflecting a state of heightened immune surveillance rather than active antiviral or inflammatory activity between implantation and labor. Together, these patterns indicate a shift from immune-dominant programmes to meeting increased cellular and physiological demand as pregnancy progresses.

### eQTL

Observed temporal changes in the regulation of the maternal immune system in modules and variance clusters further motivates investigation of whether this variation is driven by genetics through eQTL analyses. Context-specificity of genetic regulation refines the interpretation of colocalization results by demonstrating how dynamic modulation of gene expression during pregnancy can strengthen genetic associations, even at loci with baseline regulatory activity outside pregnancy. Using reproductive-aged females from the INTERVAL cohort as a comparator suggests that many signals are pregnancy-magnified, although differences between the cohorts and limited sample size constrain interpretation, underscoring the need for longitudinal sampling of the same individuals before and during pregnancy to more directly resolve context-specific genetic regulation.

A recent GWAS study demonstrated changes in genetic effects over time in pregnancy^29^, highlighting growing interest in the temporal dependence of genetic regulation in this context. Following the identification of pregnancy-specific eQTL effects and evidence of variation in genetic regulation across gestation, we systematically characterised effects that change throughout pregnancy. To do this, we introduce a time-interaction framework for eQTL analysis and interpret context-specific signals through pathway and regulon enrichment, overlap with cell proportion interactions and characterization of temporal eQTL clusters, providing a structured approach to disentangle changes in genetic effects across gestation. Previous bulk RNA-seq eQTL studies have interpreted cell proportion interactions as putative cell type-specific eQTL effects^30–32^. The large number of cell proportion interactions we observe suggest that a substantial fraction of our eQTL may reflect cell type-specific regulatory effects that cannot be resolved in bulk eQTL, motivating future single-cell eQTL studies to further resolve cell type-specific regulatory effects. Together with the observed temporal remodelling of the maternal peripheral immune landscape from RNA-seq alone, time-interaction eQTLs indicate that these changes in the gestational environment further shape the genetic regulation of transcriptional and immune processes.

### Pregnancy outcomes

Although we assessed differences in gene expression and module trajectories across pregnancy outcome groups, associated signal was limited by the low number of complicated pregnancies in this sample of the POPs2 cohort. While some analyses yielded a small number of statistically significant signals, others showed only suggestive (nominally significant) associations, suggesting that the POPs2 cohort primarily captures transcriptomic dynamics of uncomplicated pregnancy. Therefore, investigation of transcriptomic associations with complications is primarily restricted to colocalisation with GWAS loci rather than uncovering outcome-specific transcriptomic signatures. This highlights the need for future studies to recruit cohorts enriched for pregnancy outcomes of interest to increase power for detecting transcriptomic signals that will both elucidate the mechanisms underlying these conditions and enable progression towards predictive modelling of pregnancy complications in order to improve clinical outcomes. Additionally, while both maternal and fetal genetics contribute to disorders such as pre-eclampsia^33–35^, our analyses capture only maternal genetic effects on maternal gene expression, and do not include fetal genetic influences or placenta-mediated regulatory mechanisms. Future studies extending maternal sampling to delivery and incorporating post-partum paired cord blood and placental sampling will be required to disentangle distinct genetic and transcriptional contributions to pregnancy complications.

### Conclusion

This study establishes a framework for large-scale longitudinal RNA-seq and eQTL analysis throughout a perturbation. By integrating temporal gene expression, inferred cell proportions, gene co-expression, and time-interaction eQTL modelling, we demonstrate how coordinated transcriptomic and genetic variation can be decomposed into interpretable temporal programmes to capture the full spectrum of transcriptional dynamics in pregnancy. This framework provides a template for future studies of longitudinal human perturbations including, for example, disease progression or drug response.

Beyond methodological contribution, we provide a comprehensive, publicly accessible resource of maternal genomics and transcriptional variation across gestation, forming a reference baseline for healthy pregnancy. The results of these analyses are made available through an interactive web portal (https://pops2.sanger.ac.uk/) to enable exploration by the broader community (link). Clinically, these data define the expected range and timing of immune remodelling in uncomplicated pregnancy and will aid in interpreting divergent immune states in pregnancy complications and infections in future studies. By linking dynamic temporal transcriptional changes with genetic regulation, this resource provides a foundation for future studies aimed at understanding and ultimately improving maternal and fetal health.

## Methods

### Sample collection

The Pregnancy Outcome Prediction Study 2 (POPs2^36^) recruited a prospective cohort of nulliparous women from the Rosie Hospital in Cambridge. The inclusion criteria were that all women had a viable singleton fetus at the time of the dating ultrasound where the estimated gestational age was <18 weeks and the fetus had no obvious anomalies. Exclusion criteria were limited to maternal age <16 and inability to provide consent. Ethical approval for the study was obtained from East of England - Essex Research Ethics Committee (reference number 19/EE/0331). All study participants gave written informed consent.

This study collected blood samples for RNA-sequencing and genotyping from the first 1,326 participants throughout gestation. The study originally commenced in January 2020 but was suspended from 20th March-3rd August 2020 due to the COVID-19 pandemic. 127 women were recruited in the initial recruitment window (97 x 1st time-point only, 30 x 1+2), but were unable to complete the study due to COVID19 restrictions. Their outcome data were collected and samples retained. The characteristics of the women included in the current study are summarised in **Table 1**.

All estimates of gestational age were based on the dating ultrasound and the vast majority of dating was performed using a first trimester crown rump length. All cohort members had blood obtained at 12, 20, 28 and 36wkGA appointments and the 20wkGA phlebotomy also included a sample for genotyping. The actual gestational age at sampling was tightly clustered around the scheduled visits, with median (IQR) gestational ages of 12+6 (weeks + days; IQR 5 days), 20+2 (IQR 3 days), 28+2 (IQR 4 days) and 36+2 (IQR 4 days), respectively.

### Clinical data curation

Pregnancy outcome data were obtained by individual review of each participant’s electronic medical record (EMR, EPIC). Gestational diabetes mellitus (GDM) was defined as a documented diagnosis on the EMR treated with either behavioural modification (diet and exercise) and/or drugs (metformin or insulin). Women were screened for GDM risk factors using the NICE NG3 Guideline and high risk women had a 75g fasting oral glucose tolerance test^37^, typically 24-28wkGA. Spontaneous preterm birth was defined as delivery prior to 37wkGA where the participant was documented as being in labour (irrespective of the eventual mode of delivery) but excluding cases where labour was induced. Small for gestational age was defined as a birth weight less than the 10^th^ percentile by sex and gestational age. Preeclampsia was defined based on the 2013 ACOG classification^38^.

### Genotyping dataset generation

We collected 1-2ml of maternal blood from 1,113 individuals at the 20 week time-point into Nunc 1.8ml cryovials. DNA extraction and genotyping was performed by Yourgene Health (UK). DNA was extracted from 0.3ml of blood using an in-house phase separation method and the genotyping performed using Affymetrix UK Biobank arrays on an Axiom GeneTitan instrument. Genotypes were called using the Axiom Genotyping Best Practices Workflow on the full batch of samples, with 8 samples excluded prior to final genotype calling for failing default call rate (n=6) and dish QC (n=2) thresholds.

### Genotyping QC

We performed genotyping QC using PLINK v1.90b6.21^39^ following the methods described for the UK Biobank dataset^40^ on the cohort of 1,105 individuals with called genotypes.

Briefly, variants with high missingness (>5%), low minor allele frequency (MAF <1%) and variants that were not in Hardy Weinberg equilibrium (HWE, p<1×10^-9^) were excluded. We used PLINK to infer genetic sex and KING v 2.3.2^41^ to infer relatedness and population ancestry. Using this information, we excluded samples of inferred male sex (n=5), one sample from each pair of duplicates (n=2), and for related individuals the sample with fewer paired RNA-seq samples (n=3). We additionally removed individuals with elevated missing data rates (missingness >2%) and outliers for population-specific heterozygosity rate without long runs of homozygosity (n=8). The resulting genotyping cohort (with at least one paired RNA-seq sample) comprised 1,083 samples.

### Imputation

We checked the filtered genotyping dataset for strand, alleles, position, ref/alt assignments, and frequency differences versus the Haplotype Reference Consortium (HRC) reference panel (HRC.r1-1.GRCh37.wgs.mac5.sites.tab, http://www.well.ox.ac.uk/~wrayner/tools/). We then performed phasing and imputation against the HRC with the Sanger Imputation Service. We chose this reference panel because our data consist majorly of samples from individuals of European ancestry (82.21%). We filtered the imputed dataset to variants with an info score >0.70. We used the HRC reference panel to annotate variants with rsIDs, and genomic positions were lifted over from hg19 to hg38 to align with the RNA-seq build using pyliftover v 0.4.1 (https://pypi.org/project/pyliftover/). We excluded variants with high missingness (>5%), low MAF (<1%) and variants that were not in HWE (p<1×10^-5^) using PLINK (v1.90b6.27).

We detected and resolved mismatches between serial RNA-seq (see below) and genotyping samples from the same individual using MBV from QTLtools v 1.2^42^.

### RNA dataset generation

#### RNA extraction and sequencing

RNA was extracted from the PAXgene blood samples using the Perkin Elmer chemagic system (RNA Tissue10 Kit H96) in 96-well plate format, following the technical support recommendations for using the minimum lysis buffer and elution buffer options to maximise the end sample concentration (performed by Source Bioscience). The RNA samples were quantified via a UV spectrophotometric method involving a Thermo NanoDrop One C and qualified using electrophoretic separation on the Agilent Fragment Analyser. Following a pilot run in which RNA yields were lower than expected, the protocol was modified by increasing the lysate input and reducing the final elution volume to increase RNA yield and concentration.

Globin depletion was performed using a KAPA RiboErase Globin Kit (Roche). Stranded cDNA libraries were prepared for RNA-sequencing by the Wellcome Sanger Institute’s DNA Pipelines team using NEB Ultra II RNA Library Prep kits for Illumina with NEBNext Poly(A) mRNA Magnetic isolation. For the first 564 samples, 14 PCR cycles were performed but due to a high failure rate this was increased to 16 cycles for the remainder of the cohort. Equimolar pools of up to 95 samples were then pooled and sequenced on S4 XP flow cells using a NovaSeq 6000 system with 100bp paired-end reads.

#### RNA-seq QC

Bulk RNA-seq reads were aligned and quantified using multi-sample 2-pass STAR 2.7.8a^43^ and featureCounts v2.0.2^44^ by the Human Genetics Informatics (HGI) team at the Wellcome Sanger institute using their Nextflow RNA-seq pipeline (https://github.com/wtsi-hgi/rna_seq/tree/master?tab=readme-ov-file). RNA sequencing and mapping quality were evaluated using FastQC, geneBody_coverage and TIN from RSeQC v 4.0.1^45^ and RNA-SeQC v 2.3.5^46^. We checked the total number of reads, GC content, mapping rate, duplication rate, coverage and globin rates per sample and between sequencing lanes and library prep batches.

We first filtered out low-quality samples based on percent of reads uniquely mapping to the reference <70%. Next, we used principal component analysis (PCA) to identify quality metrics correlating with outliers in the dataset that would indicate low-quality samples. We removed samples with percent of reads mapping to exons <65% or with duplicate rate >90%. Finally, we identified and removed samples that were outliers for the number of genes with at least one read mapping. This resulted in a final cohort of 3,776 RNA-seq samples from 1,305 individuals.

We used CrosscheckFingerprints from GATK4 v4.3.0.0^47^ to identify sets of RNA-seq samples coming from the same individual. Comparison of CrosscheckFingerprints results to expected matches from sampling records enabled detection and resolution of sample swaps between individuals in the serial RNA-seq samples, together with MBV analysis (see above). This also identified two individuals each with 5 samples who had miscarried after an initial twelve week visit and were re-recruited.

#### Personalized HLA mapping of RNA-seq reads

We performed personalized HLA mapping in order to increase the accuracy of HLA gene expression quantification. We called HLA alleles from genotyping samples using HIBAG v1.30.2^48^, and from RNA-seq samples using arcasHLA reference version 3.52.0^49^. HLA types of individuals were determined based on agreement between genotyping and all RNA-seq samples. Next, we generated personalized reference sequences for each gene in the MHC based on each individual’s HLA types (https://github.com/davenportlab/HLApm). Reads originally mapped to the HLA region and unmapped reads were re-mapped to the personalized references. Reads mapping to the personalized references were quantified and re-introduced into the RNA-seq count matrix for use in downstream analysis.

### RNA-seq analysis

#### Gene filtering and count matrix normalization

We retained genes expressed at a CPM equivalent to >10 counts from the mean library size (10/24,100,299=0.4149326 CPM) in at least 5% of the samples (18,826 genes). Library sizes were trimmed means of M (TMM) normalized using calcNormFactors from edgeR v4.0.16^50^ and the count matrix was CPM normalized and log-transformed using voomWithDreamWeights from variancePartition v1.32.5^51^ without covariates.

#### Cell type proportion imputation and time course analysis

We used the CIBERSORTx^52^ cell fractions module to impute proportions of 22 immune cell types from the RNA-seq dataset with the following parameters: LM22 signature matrix, B-mode correction, 100 permutations. We performed paired Wilcoxon signed-rank tests to compare cell proportions between time-points and adjusted p-values using Bonferroni correction. R v 4.4.1^53^ was used for statistical analyses unless otherwise stated.

The cell proportions that had significant differences across pregnancy time-points were modeled over continuous time to better delineate their trajectory. Covariates were selected following variancePartition analysis of clinical (sample time-point, individual, education level, self-reported ethnicity, diabetes status, alcohol status, smoking status, delivery month, age, BMI) and technical (sequencing batch) covariates on imputed cell proportion values to determine which covariates contribute to variation in imputed cell proportions (**Supplementary Figure 22**). The model was fit with lmer from lmerTest v 3.1-3^54^ using bs from splines v 2.3.1^53^, following the methods of Gisby et al. (2022)^55^.

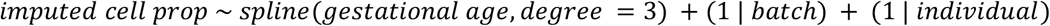

Summary statistics of measured cell fractions per week of pregnancy reported by Bar et al. (2025)^11^ were downloaded in order to compare to continuous imputed cell proportions. Time-points were subsetted to 11-37wkGA. Eosinophils, monocytes and neutrophils were compared to the same cell types in POPs2. Lymphocytes were compared to the sum of all B cells, T cells and NK cells in POPs2. In order to correlate weekly mean cell counts reported by Bar et al. (2025) and POPs2, gestational age in POPs2 was floored and weekly group means were calculated to create weekly intervals.

### Differential expression analysis

We used dream (variancePartition) to fit a differential expression model with no intercept so that each time-point coefficient was estimated directly, enabling all pairwise comparisons to be conducted. Time-point was modeled as a discrete variable (12, 20, 28, 36wkGA). We selected additional covariates following variancePartition analysis of clinical (sample time-point, individual, education level, self-reported ethnicity, diabetes status, alcohol status, smoking status, delivery month - capturing seasonality, age, BMI) and technical (sequencing batch) covariates to determine how much variation in gene expression each covariate explained (**Supplementary Figure 23**). Only covariates explaining substantial variation in gene expression (individual, sequencing batch) were included in the final model, which was fit for each gene with dream:

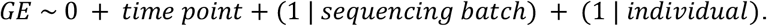

To assess contribution of cell composition on bulk RNA-seq profiles, we performed variancePartition of inferred cell proportions and compared logFC values from models including vs excluding inferred cell proportions (**Supplementary Figure 8**). These explained little additional variance and had minimal impact on differential expression estimates, so were not included as covariates in the final dream model.

We applied contrasts to identify differentially expressed genes (DEGs) between each pair of time-points independently, defining DEGs as genes with adjusted p-value <0.01 and fold change >1.5. We then performed pathway enrichment analysis on the differentially expressed gene sets at each time-point contrast using xEnricherGenes from XGR v1.1.9^56^ with Reactome pathways, and xEnrichConciser to reduce the list of differentially expressed pathways to non-redundant terms.

We validated our differential gene expression analysis using publicly available gene expression data from Gomez-Lopez et al. (2019)^9^. We included only samples taken within a 2 week window on either side of the POPs2 time-points (e.g. samples from 10-14wkGA were considered as comparable to POPs2 12wkGA time-point samples). This resulted in a dataset of 174 samples from 49 individuals aligned to our four time-points (12wkGA n = 34, 20wkGA n = 43, 28wkGA n = 56, 36wkGA n = 41).

We downloaded the Gomez-Lopez et al. (2019) gene expression data from the Gene Expression Omnibus. Microarray probes were mapped to Ensembl gene IDs using hta20transcriptcluster.db, retaining only uniquely mapped probes. Where multiple probes mapped to the same gene, expression values were averaged to obtain gene-level expression. Genes were then restricted to those present in the POPs2 dataset, and differential expression analysis was performed using the same methods as for POPs2. We subsetted the POPs2 and Gomez-Lopez et al. (2019) datasets to the intersection of genes in both datasets. Correlations of logFC estimates between the two datasets were assessed for each time-point contrast.

Separate differential expression analyses were run to contrast samples from individuals with vs. without each pregnancy complication. For each complication, differential expression was assessed at each time-point and in a combined model including samples from all time-points. Models included a term for the presence of the complication and controlled for covariates determined to contribute to variation in gene expression in the differential expression analysis between time-points:

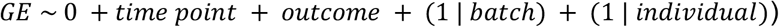

Differentially expressed genes were defined as those with an absolute value of FC >1.5 and adjusted p-value <0.05.

### Gene co-expression module definition

We performed Weighted Gene Correlation Network Analysis (WGCNA^17^) consensus module analysis to define co-expressed gene sets whilst accounting for repeat sampling from the same individual. First, we defined gene networks on data subsets from each of the 4 time-point groups separately, as detailed below. Then, we formed a consensus network from all 4 time-point specific networks and defined consensus modules. Modules were defined using the signed hybrid network type, which modifies any negative correlations between genes to a correlation of 0 in order to group only positively correlated genes into modules (for enhanced interpretability: groups of genes changing in the same direction over time).

Gene networks were defined on each time-point as follows. We corrected each time-point expression matrix for confounding technical variation using sva_network from sva v3.50.0^57^ with n.pc=20. We calculated biweight midcorrelation matrices from each gene expression matrix using bicor from WGCNA v1.72-5^17^. Mean-correlation bias in the correlation matrices was corrected using normalize_correlation from SpQN v1.14.0^58^ with parameters ngrp=10, size_grp=2000, ref_grp=10. We performed soft thresholding on the normalized correlation matrices using pickSoftThreshold.fromSimilarity from WGCNA, updating negative correlation values to 0 as required for signed hybrid network construction. Using a soft threshold of 13 (**Supplementary Figure 24**), we generated signed hybrid adjacency matrices to define the similarity between all genes in each time-point dataset, then generated a topological overlap metric (TOM) matrices from the adjacency matrices as described by Langfelder and Horvath (2008)^17^.

Separate TOM matrices were scaled to the 95th percentile to control for the differing statistical properties across time-point datasets, in order to reduce bias in consensus modules as described in the WGCNA consensus module tutorial. The consensus TOM was calculated by taking the component-wise minimum of the individual TOMs. Consensus modules were generated by clustering the consensus TOM matrix with hclust from stats and cutreeDynamic from dynamicTreeCut v1.63.1^59^. Parameters were optimized to generate modules with a minimum of 70 genes for optimal interpretability using parameters deepSplit=4, minClusterSize=50 (**Supplementary Figures 10,25**). Module eigengenes were calculated using the multiSetMEs function from WGCNA.

Module biological functions were inferred by annotating each module gene sets as follows: (1) pathway enrichment analysis with XGR and Reactome pathways (adjusted p-value threshold <0.05); (2) enrichment with hypergeometric test (adjusted p-value <0.05) for cell type-specific marker genes from xCell v1.1.0^60^, subsetted to the cell types in CIBERSORTx deconvolution; (3) positive Spearman correlation (Rho >0.3, adjusted p-value <0.05) between CIBERSORTx imputed cell proportions and module eigengene values at the 12 week time-point; and (4) enrichment with Fisher’s exact test of regulon gene sets from DoRoTHEA^23^, defined using decoupleR v2.8.0^61^ and parameters organism=human, levels=A,B,C, and restricted to genes expressed in the POPs2 dataset.

### Gene co-expression module analysis

Repeated measures ANOVA was performed to test for association between eigengene value and discrete time-point, and p-values were FDR-adjusted. The first sample for each of two individuals who had miscarriages and re-enrolled in the study (resulting in two 12 week samples) were removed before this analysis.

Due to significant differences in eigengene expression in all modules across time-points (**Supplementary Figure 26**), linear mixed-effects models of module eigengenes were fitted to identify continuous patterns of change in module expression over time. Covariates were selected following variancePartition analysis of clinical (sample time-point, individual, education level, self-reported ethnicity, diabetes status, alcohol status, smoking status, delivery month, age, BMI) and technical (sequencing batch) covariates on module eigengene values (**Supplementary Figure 27**). The model below was fitted using bs from splines.

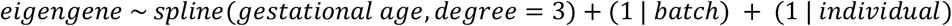

Modules were clustered based on eigengene expression in order to systematically group modules with similar expression patterns for visualization (**Supplementary Figure 11**).

Module eigengenes were also tested for differential trajectories across outcome groups by adding an interaction term with outcome as described by Gisby et al. (2022)^55^.

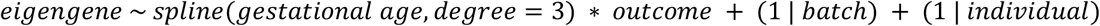

An ANOVA test was used to determine the significance of the interaction between the trajectory of gestational age and the outcome (for each outcome separately) in order to identify module eigengenes with significantly different trajectories over time across outcome groups, and p-values were FDR adjusted.

To compare temporal patterns of module eigengenes with related laboratory blood tests, weekly summary statistics for coagulation and red blood cell measures were obtained from Bar et al. (2025) for 11–37wkGA (corresponding to the minimum and maximum gestational ages observed in POPs2). Gestational age in POPs2 was floored to whole weeks and samples were grouped by week to match the weekly means reported by Bar et al. Weekly mean module eigengene values were then calculated for each gestational week and compared with the corresponding weekly mean laboratory values. Pearson correlations between weekly module eigengene values and weekly lab test values were computed.

### Gene expression variance analysis

For variance analysis, only individuals with all four RNA-seq time-points were included, enabling a paired design controlling for inter-individual variation. Batch correction of gene expression was performed by obtaining residuals from linear regression of gene expression values on sequencing batch as a random effect using lm from stats. The Levene test of heteroskedasticity was performed between all time-point pairs using leveneTest from the car package v3.1-2^62^ to identify genes with changes in variance over time, and p-values were Benjamini & Hochberg adjusted within time-point pairs.

To group together genes with similar patterns of variance over time, genes were filtered to those with evidence of heteroskedasticity (adjusted p-value <0.05, abs(FC) >1.5 in at least one contrast). Gene variance was calculated at each time-point using var from stats, and variance ratios were calculated as the variance in expression of a gene at the later time-point divided by the variance in expression of the gene at the earlier time-point for each time-point pair. Variance ratios were then used to calculate log_2_ fold change (log2FC) values from the earlier to later time-point. For time-point pairs where the heteroskedasticity test was non-significant, log_2_FC values were set to zero. Log_2_FC values were scaled across genes within each time-point comparison group using StandardScaler from scikit-learn v1.5.1^63^ to avoid biased weighting of specific time-points in subsequent clustering. Hierarchical clustering was applied to the scaled log2FC values from all pairwise comparisons using AgglomerativeClustering from scikit-learn and number of clusters, k, set to 3–16 clusters. For each clustering, sum of squared distances of genes within the cluster and the minimum number of genes in a cluster were calculated. Six clusters were chosen to maximize the size of the smallest cluster while minimizing within cluster sums of squared differences (**Supplementary Figure 28**). Pathway enrichment of genes in each cluster was performed using xEnricherGenes and xEnrichConciser from XGR (Reactome ontology), where the background included all 18,826 genes expressed.

### eQTL mapping

eQTL mapping was performed with a linear mixed model using 3,394 RNA-seq samples that had a paired genotyping sample from the same individual. The top 6 genotyping PCs were included in all eQTL models to account for population stratification. The number of genotyping PCs to include was determined based on the amount of variation captured (**Supplementary Figure 29**). Imputed neutrophil and monocyte proportions centered around 0 were included (**Supplementary Figure 30**). PEER factors calculated from the gene expression data using peertools v20120508^64^ were included in the eQTL model in order to control for unknown sources of variation in gene expression. Fixed effect covariates were held out in the calculation of PEER factors, including binary time-point variables, centered imputed neutrophil and monocyte proportions, 6 genotype PCs and a covariate for the mean (using the add-mean parameter). PEER can not handle random effects, so individual and batch were not held out of the PEER factor calculation but were also included in the eQTL models. 50 PEER factors were calculated, and the number of PEER factors to include in eQTL models determined by mapping eQTL on chromosome 1 with varying numbers of PEER factors (0-50, increasing by 5). 30 PEER factors were included because this number maximized eGene discovery (**Supplementary Figure 31**).

SNP-gene pairs were tested for eQTL for SNPs within 1Mb on either side of the transcription start site of expressed genes. HLA-DRB3, 4 and 5 were excluded from eQTL mapping as not all individuals have these genes, depending on their HLA-DRB1 haplotype.

### Main effects eQTL mapping

First, the main effects model was fit using lmer from lme4 v1.1-35.2^65^. Time-point was included as a factor, rather than a continuous variable to allow for non-linear effects, which was appropriate as time-points were non-overlapping (**Supplementary Figure 32**).

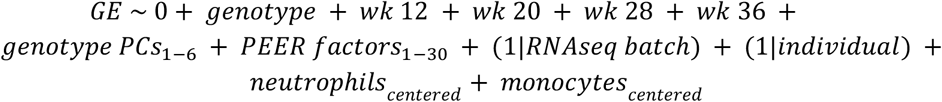

Models that failed to converge were re-fitted up to 50 times restarting from the original value and up to 50 times from slightly perturbed values or until they converged as recommended by the lme4 package. Nominal p-values were determined by comparing the null model (main effects model without genotype effect) to the main effects model using anova from stats.

Gene-specific nominal significance thresholds were determined based on the effective number of tests for each gene as estimated by eigenMT^66^ and Benjamini-Hochberg correction across genes, as described by Burnham et al. (2024)^32^. SNP-gene pairs with nominal p-values below the corresponding gene-specific threshold were considered significant.

### Assessing context-specificity of pregnancy eQTL

POPs2 eQTL results were compared to eQTL mapped in a non-pregnant blood donor cohort from the same UK population as part of the INTERVAL study^21^ to identify pregnancy-specific genetic regulation of gene expression. In order to facilitate the most direct comparison without confounding effects of age, sex, and pregnancy time-point, the INTERVAL dataset was subsetted to females of reproductive age for eQTL mapping, and the POPs2 cohort was subsetted to individuals with RNA-seq samples at all four time-points to be re-mapped at each time-point separately.

### Mapping eQTL in females of reproductive age in the INTERVAL cohort

The INTERVAL RNA-seq dataset was subsetted to females of reproductive age (<45 years old, n=445, **Supplementary Figure 33A**). Chromosome X was lifted over from hg19 to hg38 as described previously for the autosomes, and the genotyping dataset was subsetted to females of reproductive age and re-filtered to exclude variants with high missingness (>5%) and low MAF (<5%) using PLINK. A MAF threshold of 5% was used to minimize the FDR for a dataset of this size as described by Huang et al. (2019)^67^. Using the same procedures described above for the POPs2 data, the INTERVAL raw count matrix was filtered and normalized, eQTL covariates were selected (**Supplementary Figure 33B-D)**, the eQTL model was fitted, and multiple testing correction was performed.

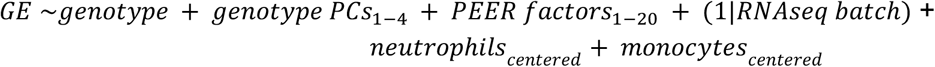

### Mapping eQTL in POPs2 per time-point

In order to maintain comparable power between datasets and investigate the effect of each pregnancy time-point independently, POPs2 eQTL were re-mapped separately per time-point in a subset of the cohort with samples at all 4 time-points (n individuals=448). SNPs tested were filtered to exclude variants with low MAF (<5%) in this data subset to minimize the FDR^67^. eQTL covariates were selected and the eQTL model was run and corrected for multiple testing as described for the full POPs2 dataset above, including re-selecting the number of PEER factors (**Supplementary Figure 34**).

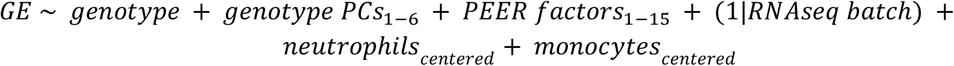

### Comparison of signals identified in pregnancy vs non-pregnant eQTL

For each time-point specific eQTL dataset, lead SNP-gene pairs also present in INTERVAL were extracted and the effect alleles aligned between datasets. Context-specificity of eQTLs was assessed using the mash framework from mashr v0.2.79^68^. First, a strong test subset was defined consisting of the union of significant lead SNP-gene pairs across all pregnancy time-points. A random test subset was defined as 1% of all SNP-gene pairs tested in both the POPs2 and INTERVAL cohorts.

To account for correlations across the five datasets (four pregnancy time-points and the INTERVAL cohort), the correlation structure was estimated from the random subset using estimate_null_correlation_simple and incorporated into the mash analysis. Data-driven covariance matrices were calculated from the strong test subset using cov_pca with five principal components, reflecting the major axes of variation across datasets, and further deconvoluted with cov_ed. Canonical covariance matrices were calculated from the random subset using cov_canonical. Both data-driven and canonical covariance matrices were included in the mash model to capture both complex, data-specific patterns of sharing and simple, interpretable patterns. Finally, posterior effect sizes for the strong test subset were computed using the fitted mash model learned from the random test pairs.

The resulting mashr posterior effect sizes were used to characterize context specificity of results. The lead SNP-gene pairs for each time-point were considered independently. eQTL were characterized as “not significant” if the mashr local false sign rate for the POPs2 result was > 0.05. Remaining effects were were categorized as (1) “shared” if effect sizes had the same sign and were within a factor of 0.5 of each other or (2) “opposite direction of effect” if their effect sizes had different signs but were both significant, or (3) “pregnancy-magnified” if the effect was only significant in POPs2 (not INTERVAL) or had a larger effect size in POPs2, or (4) as “pregnancy-dampened” if the effect size was larger in INTERVAL.

### Characterization of pregnancy-magnified signals

Pathway enrichment of pregnancy-magnified eGenes was performed using XGR as described above. Regulon enrichment of pregnancy-magnified eGenes was performed using DoRothEA regulons as described above. In both pathway and regulon enrichment analyses, for each time-point, the enrichment background consisted of all eGenes whose lead SNP-gene pair was also tested in the INTERVAL eQTL analysis (all genes included in the mashR comparison for that time-point). For visualization, fold enrichment of regulon membership was calculated in the same way as in XGR where

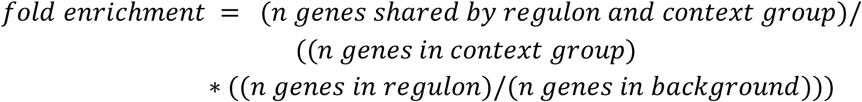

This metric quantifies how strongly pregnancy-magnified eGenes are enriched in each regulon by comparing observed gene overlap to gene overlap expected by chance.

### Fine mapping and colocalization

The Flanders pipeline (https://github.com/Biostatistics-Unit-HT/Flanders) was used for fine-mapping and colocalization across multiple sets of summary statistics. These included POPs2 main-effect eQTL, INTERVAL eQTL from the full cohort^21^ and the best-powered publicly available GWAS in majority European populations of gestational diabetes mellitus^12^, pre-eclampsia^13,14^, preterm delivery^15^ and gestational duration^15^.

For all studies, the Flanders input parameters were set to a hole size of 200,000 and a credible set threshold of 0.95. Flanders requires two significance thresholds to define genomic regions for fine-mapping. A primary significance threshold (p_thresh1) determines whether a region is selected for fine-mapping, and a secondary inclusion threshold (p_thresh2) filters the set of variants included and therefore determines the size of the fine-mapped region. We implemented an update to the Flanders pipeline allowing for the input of per-gene significance thresholds where appropriate (i.e. for eQTL studies, github.com/wtsi-hgi/Flanders/tree/per-gene-p-value). In eQTL datasets where per-gene significance thresholds from eigenMT were available, p_thresh1 was defined as the gene-specific significance threshold from eigenMT. The corresponding variant inclusion threshold, p_thresh2, was determined using a scaled approach based on p_thresh1 with an upper bound to capture the full signal while constraining the region size:

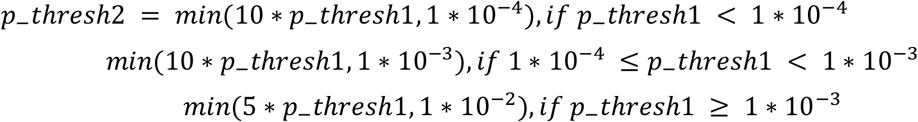

Due to limited power of GWAS for pregnancy traits, for GWAS studies, p_thresh1 was set to 1×10^-5^, corresponding to genome-wide suggestive significance, and p_thresh2 was set to 1×10^-4^.

Flanders was run with parameters run_liftover=FALSE, chromosomes=1-23 to include the X chromosome and susie_max_iter=1,000.

eQTL signals were fine-mapped within the Flanders pipeline using SuSiE^69^. Loci were excluded from fine-mapping if they overlapped the HLA region (n=197), exceeded the size threshold (n=144), or did not yield any credible sets (n=8).

Colocalizations with posterior probability for hypothesis 4 (PPH4) >0.8 were considered significant. Two pre-eclampsia GWAS were included for colocalization. In two instances there were colocalizations between the two GWAS for PE and a pregnancy eQTL (chr16:28516005 and chr15:90887387). To avoid counting the same colocalization signal twice, the FinnGen colocalizations were excluded.

Regional association plots of colocalizations were created using locuszoomr v0.3.8 (https://github.com/myles-lewis/locuszoomr) using Ensembl 99 EnsDb for Homo sapiens (AH78783) from AnnotationHub v 3.10.0^70^. Plot points were colored by LD (Plink r2) with the joint lead SNP between the POPs2 and GWAS colocalization. The joint lead SNP was selected from the group of overlapping SNPs present in both the POPs2 eQTL and GWAS SuSiE credible sets. Each SNP was ranked by decreasing lABF (log approximate Bayes factor, quantifying the strength of evidence that a variant is associated with the trait) in both datasets, and the joint rank was calculated as the sum of the eQTL and GWAS rank. The SNP with the lowest joint rank was selected as the joint lead SNP.

### Conditional eQTL mapping

In genes where Flanders fine-mapped signals in only one locus, all signals were considered to be conditionally independent. Where signals were discovered in more than one locus in the same gene, we performed an additional filter for conditional independence. For these genes, we re-mapped each eQTL signal conditioning on all other Flanders fine-mapped signals from the same gene. eQTL that remained significant despite the inclusion of other signals were considered conditionally independent. In genes where no signals were conditionally independent after controlling for all others, only the top signal (lowest p-value) was considered conditionally independent (n=100). All conditionally independent signals were taken forward for eQTL interaction mapping.

### Mapping eQTL interaction models

eQTL interaction models for genotype-by-time-point, genotype-by-imputed neutrophil proportion and genotype-by-imputed monocyte proportion were fit for conditionally independent eQTL signals. For genes with multiple conditionally independent eQTL signals, the other conditionally independent signal SNPs for each gene were included as covariates to isolate the interaction effect of the tested signal. SNP-gene pairs were only tested for an interaction if there was more than one minor allele homozygous individual in each group (time-point or half of imputed cell proportion distribution). If a gene contained multiple conditionally independent SNPs, all other signal SNPs were controlled for in the model. Significance was determined by comparing the model with vs without the interaction term using anova from stats. For each of the three interaction models fit, p-values were Benjamini–Hochberg adjusted within each model. SNP-gene pairs with adjusted p-values <0.05 were considered to have significant interaction effects.

### eQTL interaction with time

eQTL interaction models with time-point were fit with the same covariates as the main effects model, additionally controlling for the remaining conditionally independent eQTL signal SNPs for that gene, as identified in the conditional analysis.

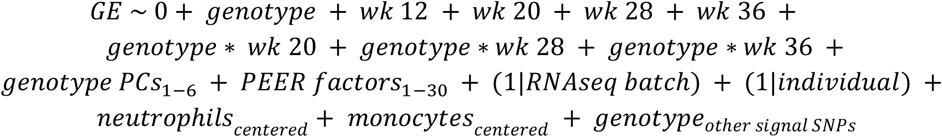

The slope (beta value) at each time-point was extracted from significant time interactions. To prepare these genes for clustering, beta values were transformed to emphasize the magnitude of temporal changes in effect size rather than the direction by multiplying time-point beta values by the sign of the main effect beta. Following transformation, beta values were scaled on a per gene, or row wise, basis so that clustering would focus on the pattern of change in beta values over time within each gene, regardless of differences in magnitude between genes. For each gene, values were standardized to have a mean of zero and a standard deviation of one using the z score transformation from the scipy.stats module in SciPy v1.11.0^71^, with the standard deviation calculated using a degrees of freedom parameter of one. Hierarchical clustering was applied to genes with significant time-point interactions based on their transformed beta values at each time-point, using AgglomerativeClustering from scikit-learn with the number of clusters (k) ranging from 3-16. For each clustering resolution, the within-cluster sum of squared distances and the smallest cluster size were recorded. The optimal number of clusters was chosen to minimize the within-cluster sum of squared distances while keeping the size of the smallest cluster greater than 50 eQTL, as no strong elbow was observed in the sums of squared distances. The final clustering used k=5 (**Supplementary Figure 35**). Enrichment analyses were performed with a background of all conditionally independent eQTL (corresponding to the full set of eQTL tested for interactions) on 6 groups: all significant interaction eQTL and each eQTL interaction cluster. Pathway enrichment was performed on clusters using xEnricherGenes from XGR. Regulon enrichment was performed on DoRothEA regulon gene groups from get_dorothea from decoupleR as described in the WGCNA module analysis section above. Regulon activity scores for SMAD3 and STAT1 regulons were calculated using run_ulm from decoupleR with minsize=5. For each regulon, changes in regulon activity scores over time were modeled using formula *score* ∼ *time point* + (1 | *individual*). Estimated marginal means were extracted for each time-point using emmeans from emmeans v1.10.0^72^ and all pairwise differences between time-points were tested for significance using pairs from graphics v4.4.1^53^ and Benjamini-Hochberg adjusted. The position of eSNPs in SMAD3 and STAT1 TF binding motifs was determined using motifbreakR from motifbreakR v4.16.0^73^ using pwmList=motifbreakR_motif, threshold=0.85, verbose=TRUE and show.neutral=TRUE.

eQTL interaction with imputed cell proportions

Interaction models with neutrophil and monocyte proportions were fit using the models below.

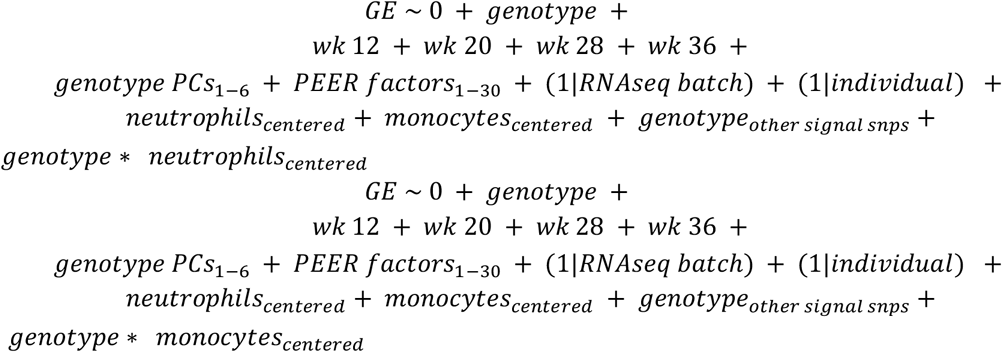

To further assess whether time-interaction eQTL were independent of cell composition effects, we fit extended interaction models to include both genotype-by-cell proportion and genotype-by-time interaction terms.

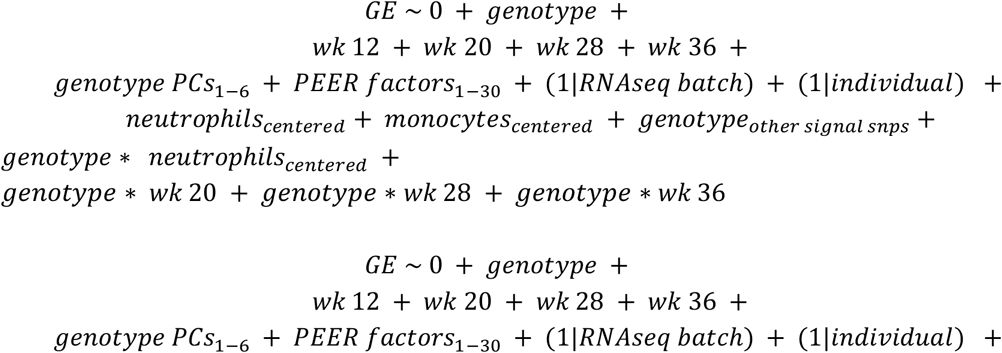

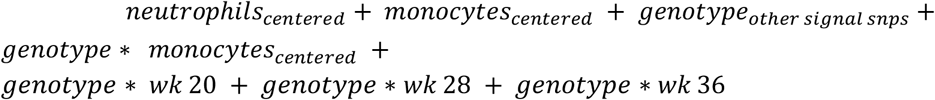

## Contributions

Conceptualization: E.E.D., D.S.C-J., G.C.S.S.; Data curation: K.L.B., E.C., S.H., U.S.; Formal analysis: K.L.B., C.E.C., S.H.; Funding acquisition: E.E.D., D.S.C-J., G.C.S.S.; Investigation: K.L.B., E.C.; Methodology: All authors; Project administration: K.L.B., E.C., U.S.; Supervision: E.E.D., D.S.C-J., G.C.S.S.; Visualization: S.H.; Writing – original draft: K.L.B., S.H.; Writing – review & editing: All authors.

## Conflicts of interest (last five years)

The authors declare the following competing interests: G.C.S.S and D.S.C-J have received research support from Roche Diagnostics Ltd for studies of diagnostics and screening for fetal growth restriction and preeclampsia. G.C.S.S has been a member of a Data Monitoring Committee for GSK trials of RSV vaccination in pregnancy. G.C.S.S is currently a member of a Data Monitoring Committee for RSV vaccination in pregnancy (Moderna) and chairs a Data Monitoring Committee for a hyperemesis gravidarum therapeutic trial (NG Biopharmaceuticals). G.C.S.S and D.S.C-J have received nonfinancial support from Illumina. G.C.S.S and D.S.C-J have received support from Pfizer (outside the scope of this work). G.C.S.S and D.S.C-J are paid consultants to Medicines360. D.S.C-J is a paid consultant to GondolaBio and G.C.S.S is a paid consultant to Exeltis. The remaining authors report no conflict of interest.

## Funding

This research was funded by the Wellcome Trust [Grant numbers 215524/Z/19/Z, 220540/Z/20/A and 206194]. For the purpose of Open Access, the author has applied a CC BY public copyright licence to any Author Accepted Manuscript version arising from this submission.

## Acknowledgements

We thank the Wellcome Sanger Institute’s Sequencing Operations team for support with sample processing and sequencing library preparation. We thank the Human Genetics Informatics team for their support with data preparation and software development, specifically Sendu Bala, Vivek Iyer, Matiss Ozols, Iaroslav Popov, Michael Woolnough, and Gennadii Zakharov. The Flanders pipeline was developed by the Biostatistics and Genome Analysis Units at Human Technopole, in particular thanks to Bruno Ariano, Edoardo Giacopuzzi, Arianna Landini, Sodbo Sharapov, and Nicola Pirastu. We are grateful to the POPs2 study participants and thank all the midwifery, research and laboratory staff for support with sample collection, with particular thanks to Amy Sutton-Cole, Katrina Holmes and Josephine Gill.

## Additional information

Supplementary Information is available for this paper.

## Data availability

The RNA-Seq dataset generated in this study are available under managed access with accession number EGAS00001006379 (will be made public upon publication). The genotyping dataset generated in this study are available under managed access with accession number EGAS00001006379 (will be made public upon publication). Source data for Figs. 1 - 6 and Supplementary Figs. 1-35 are available on Zenodo (10.5281/zenodo.22249714). Publication of individual patient data and clinical characteristics is precluded by the ethics approval. Results are browsable in web portal format (https://pops2.sanger.ac.uk/).

## Code availability

All custom code used for the current study will be publicly available from: https://github.com/davenportlab/POPs2_publication/

## Correspondence and requests for materials

These should be addressed to Emma Davenport and Gordon C S Smith

## Notes

### Author Declarations

East of England - Essex Research Ethics Committee gave ethical approval for this work (reference number 19/EE/0331).

