## Supplementary figures for "Maternal immune gene expression trajectories and dynamic genetic regulation throughout pregnancy"

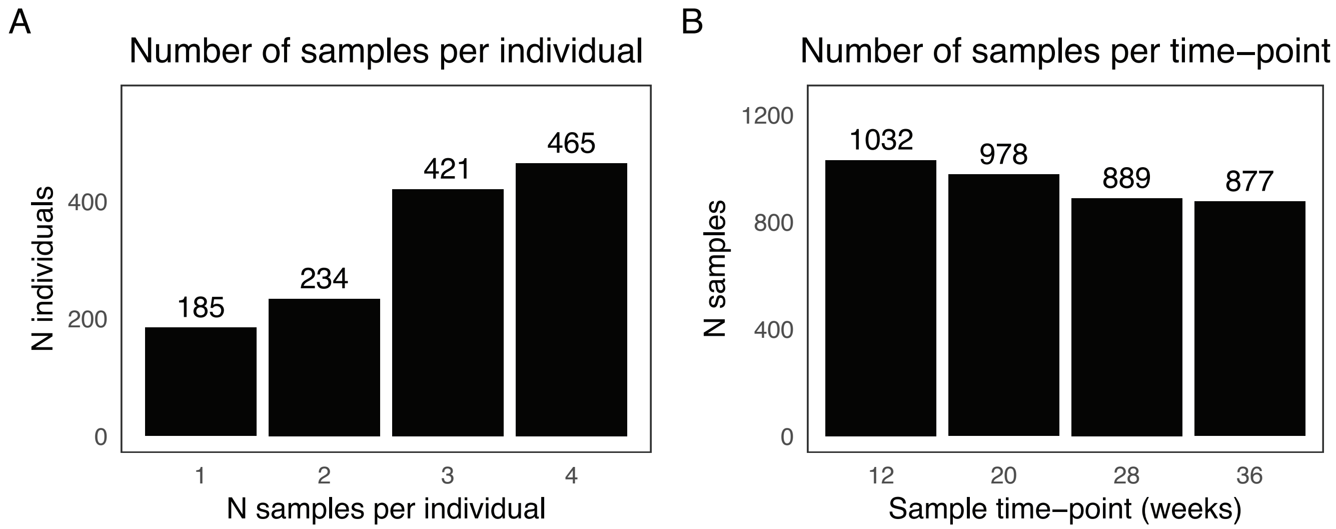


Supplementary Figure 1.
A. Bar plot of number of RNA-seq samples per individual. B. Bar plot of Number of RNA-seq samples per time-point.

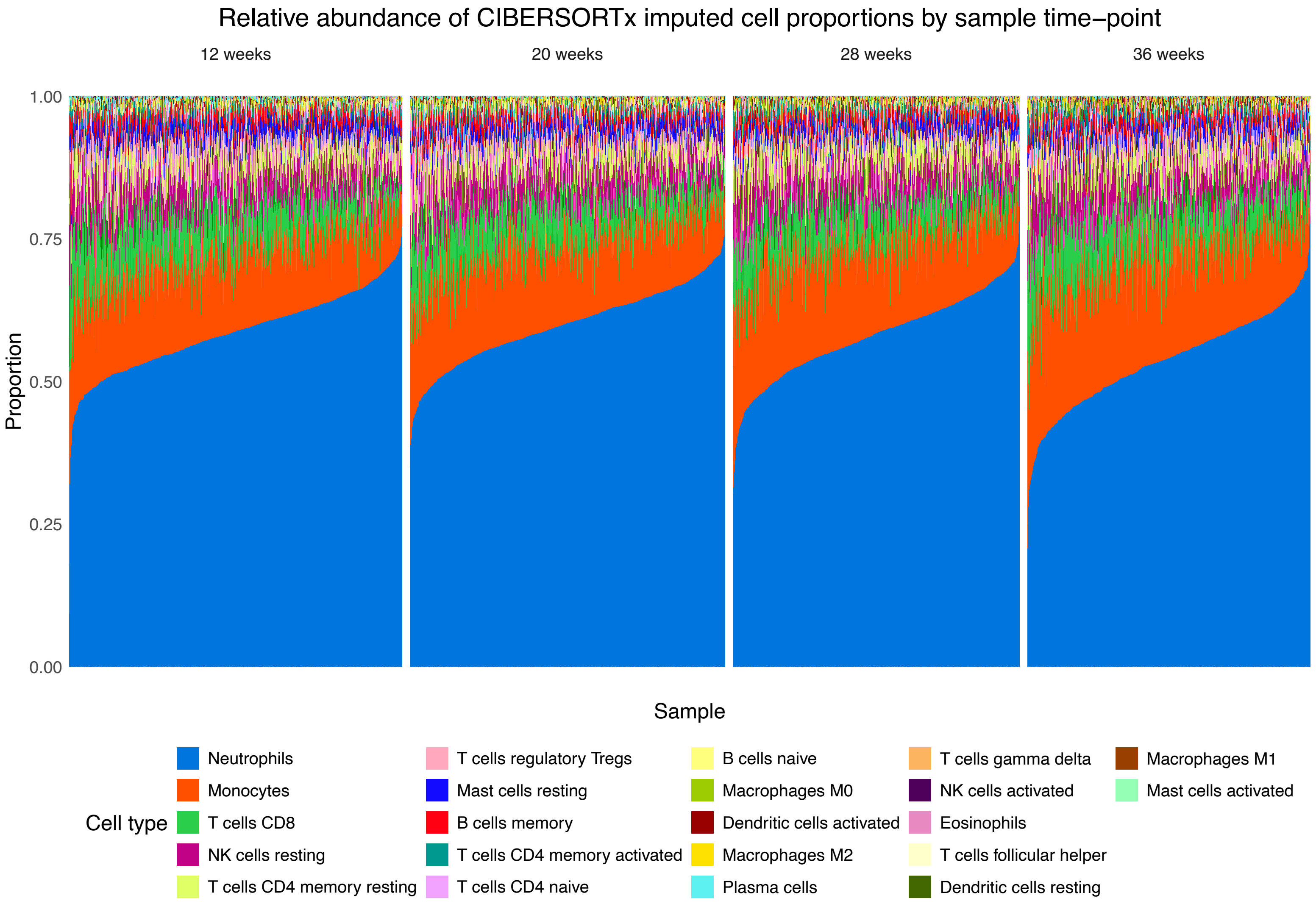


Supplementary Figure 2.
Stacked bar chart showing relative abundance of each of the LM22 cells type in bulk RNA-seq samples by time-point.

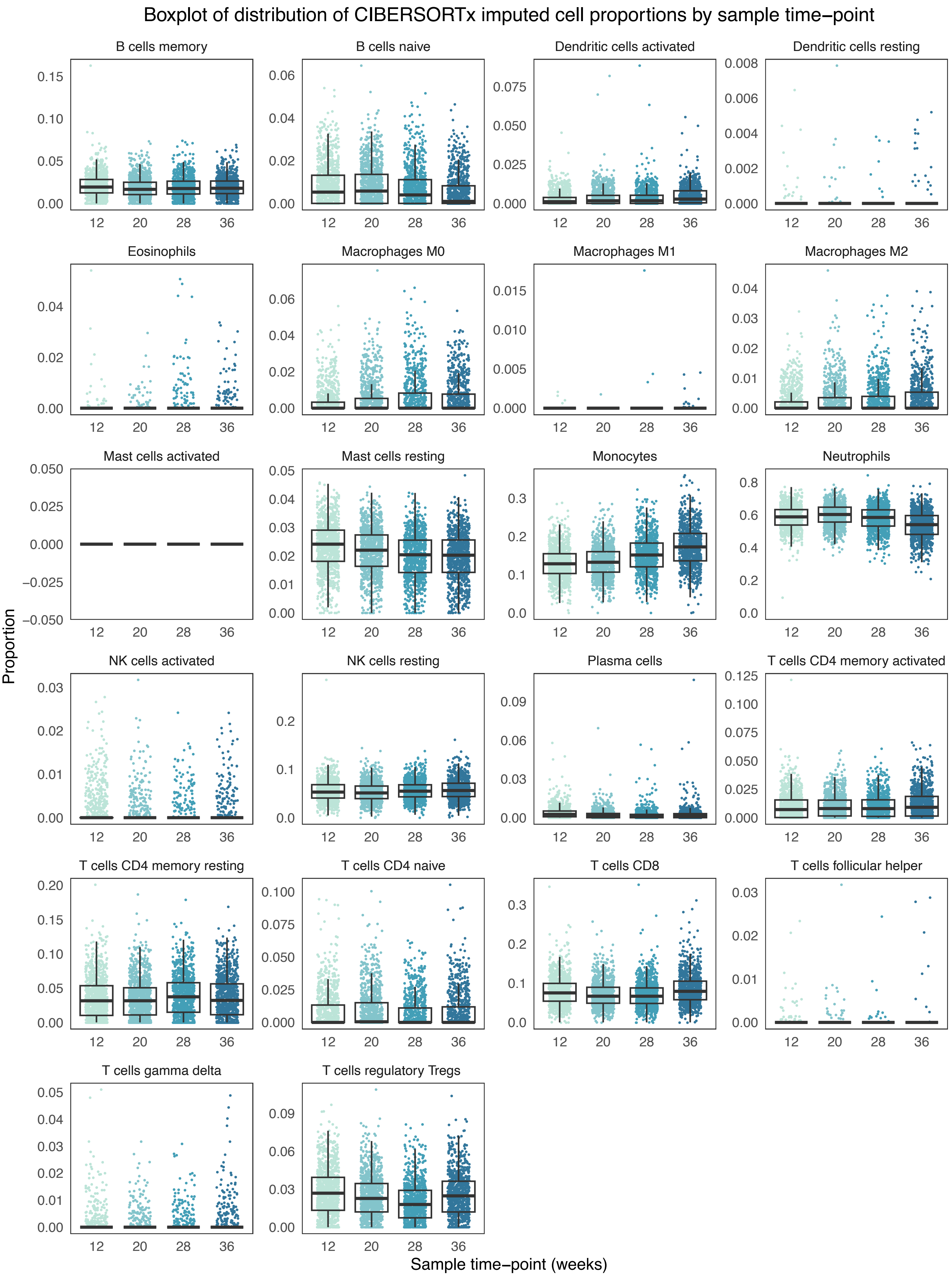


Supplementary Figure 3.

Boxplot of distribution of CIBERSORTx imputed cell proportions across sample time-points and cell types.

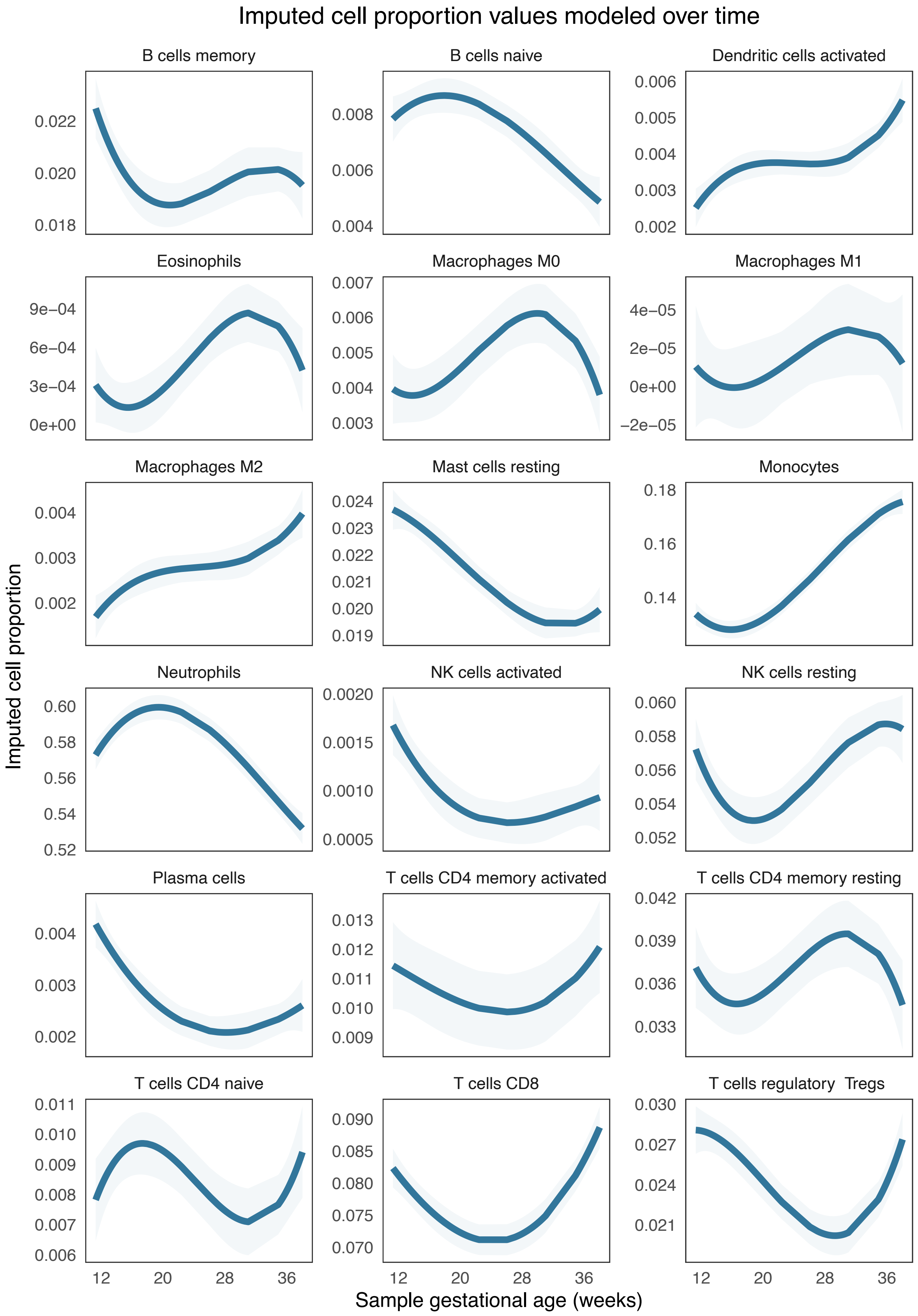


Supplementary Figure 4.

Continuous trajectories of cell proportions where curves represent predicted cell proportion values over time and shaded areas represent 95% confidence interval of predicted values. Only cell types that show significant differences in imputed cell proportions over time are shown

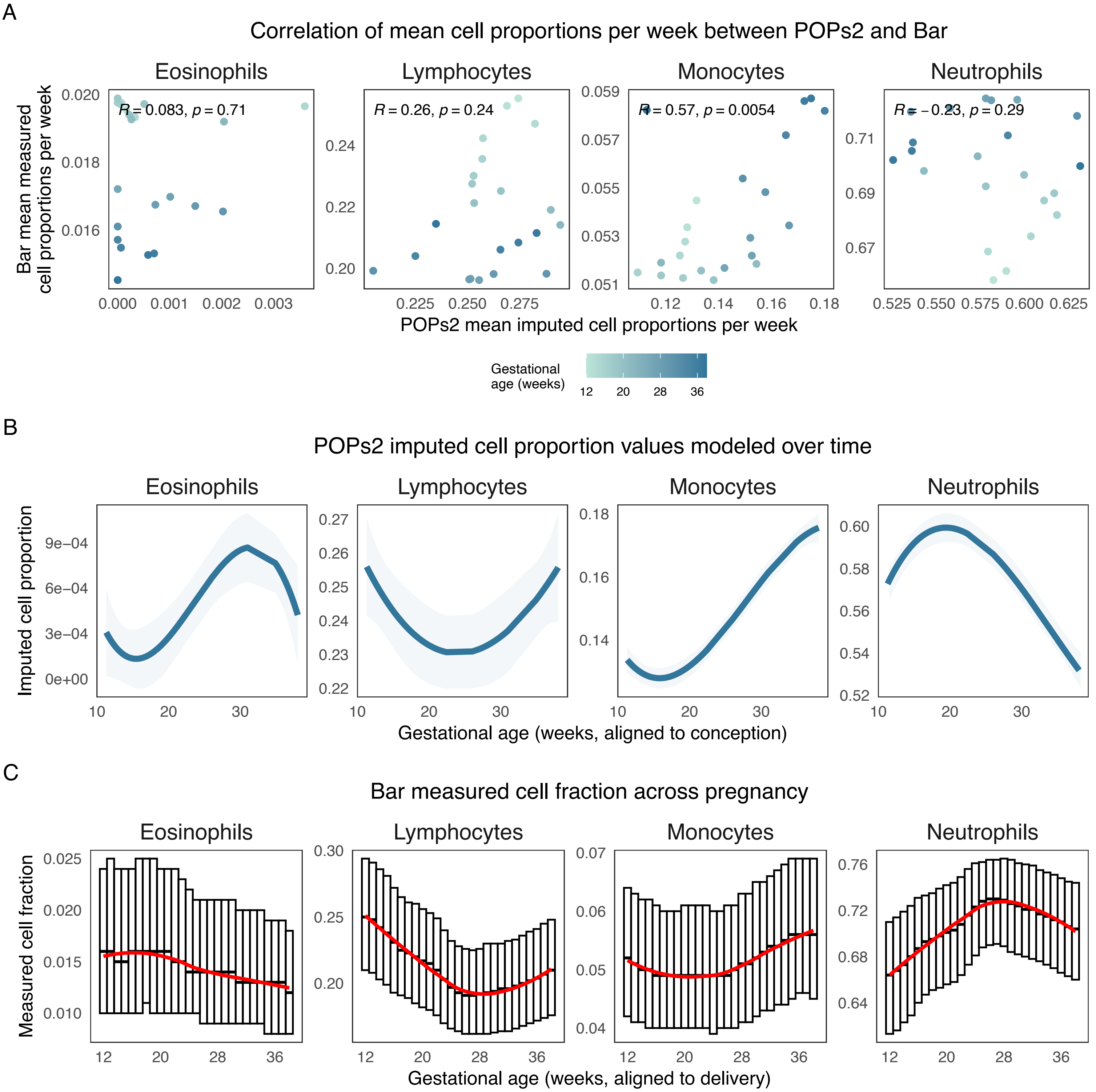


Supplementary Figure 5.
A. Scatterplot of correlation between weekly means of POPs2 imputed cell proportions and Bar et al. (2025) measured cell fractions. Points colored by gestational age. Plots annotated with correlation coefficient and its significance by cell type. B. Continuous trajectories of imputed cell proportions for cell types reported by Bar et al. (2025), where curves represent predicted cell proportion values over time and shaded areas represent 95% confidence interval of predicted values. The lymphocyte cell type is the sum of all B cell subtypes, T cell subtypes and NK cells. C. Weekly median and IQR of measured cell counts from Bar et al. (2025), with curves fitted to median values displayed in red.

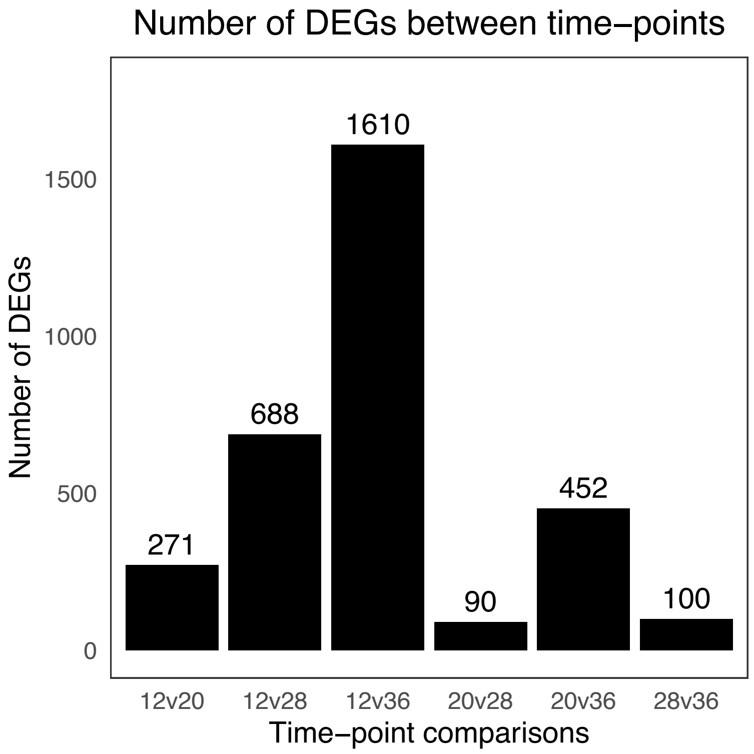


Supplementary Figure 6.
Bar chart of quantification of differentially expressed genes between each combination of time-points.

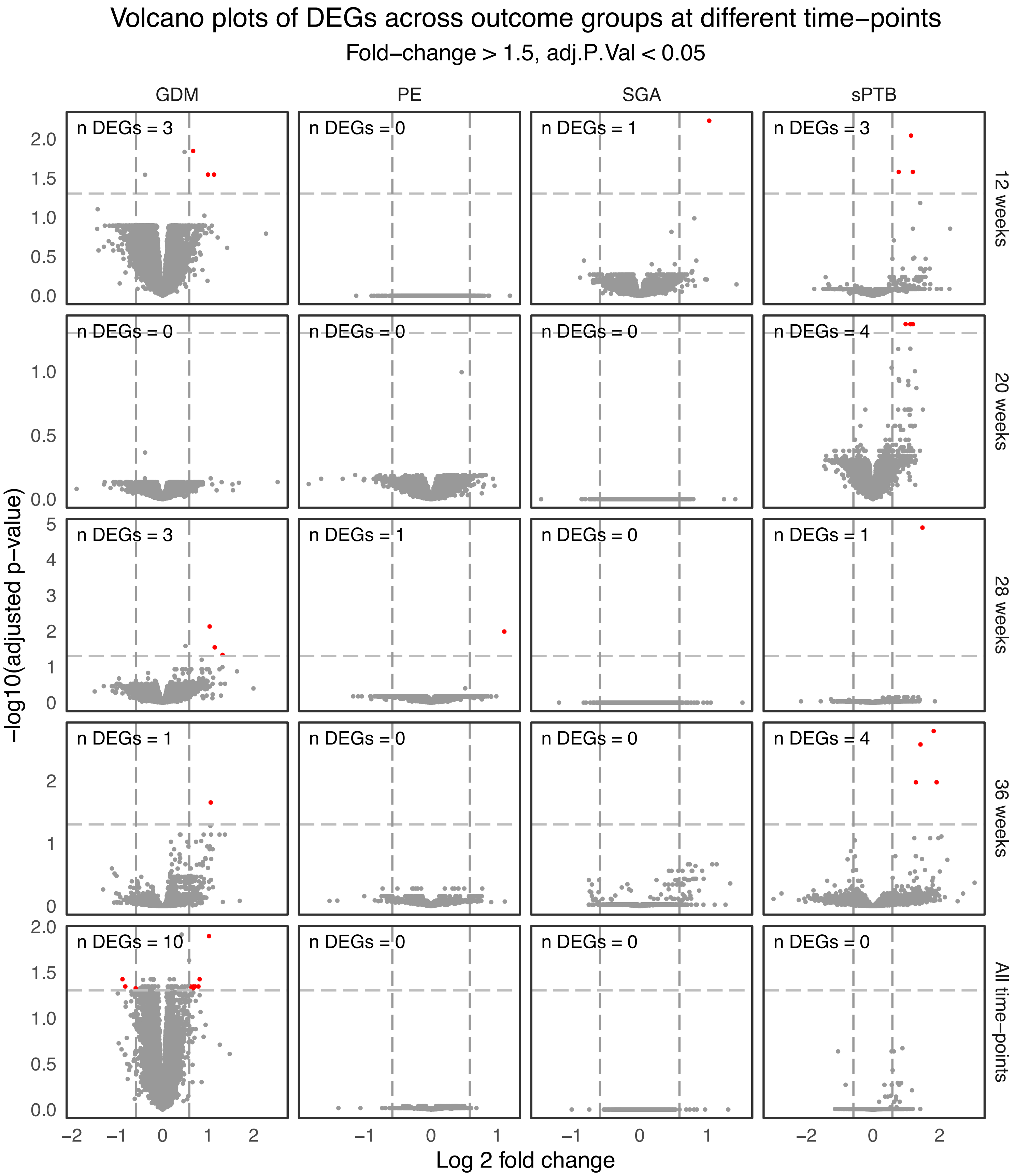


Supplementary Figure 7
Volcano plots of log 2 fold change vs. -log 10 adjusted p-values. Fold change cutoff = 1.5, adjusted p-value cutoff = 0.05. Subplots specify outcome (x) vs. time-point of samples (y) in differential expression analysis.

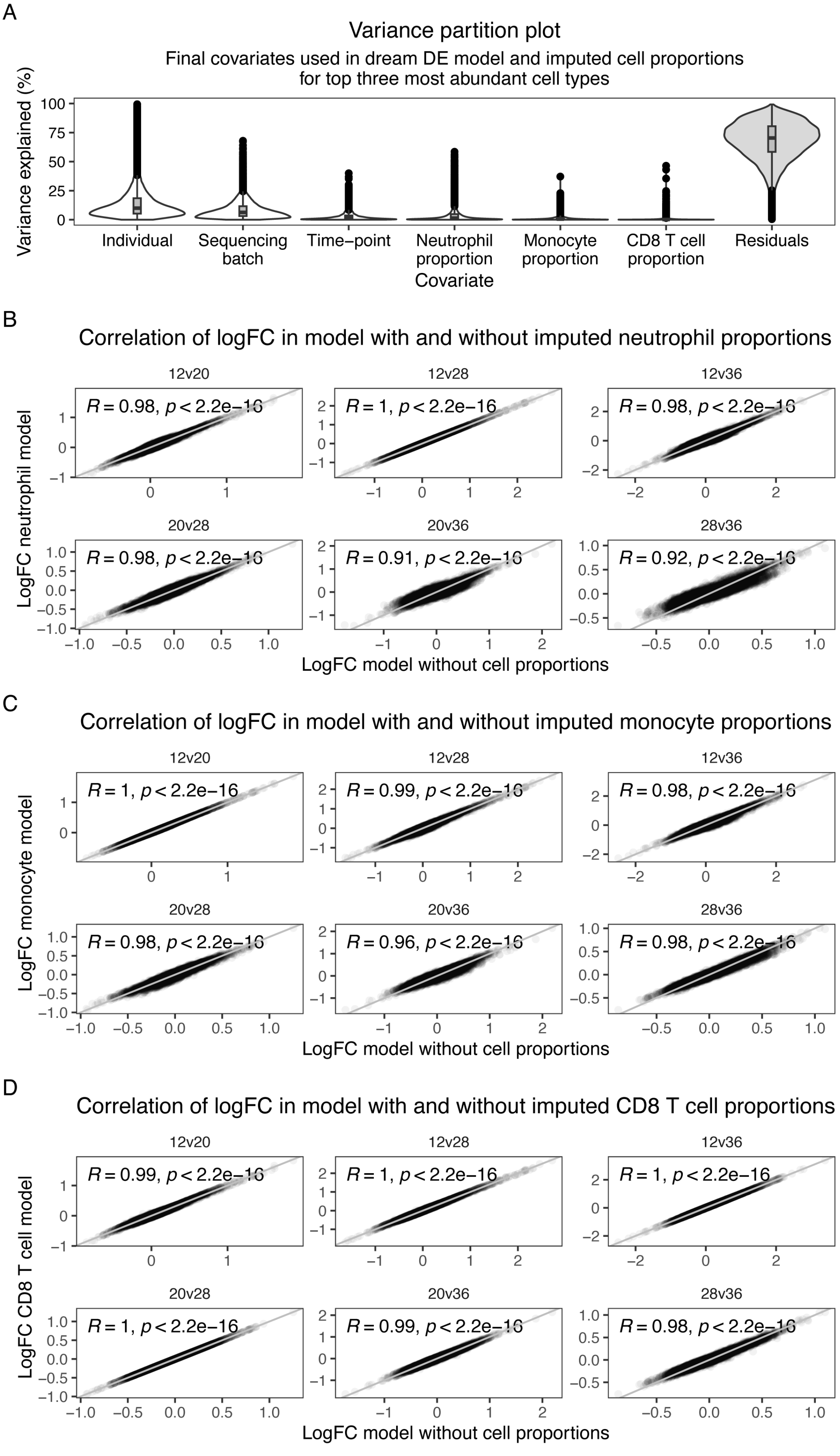


Sipplementary Figure 8.
A. Proportion of gene expression variance explained (y) by covariates used in gene expression model and imputed cell proportions for the three most abundant cell types – neutrophils, monocytes, CD8 T cells (x). Scatterplots of logFC in model including vs. excluding imputed cell proportions in B. Neutrophils, C. Monocytes and D. CD8 T cells.

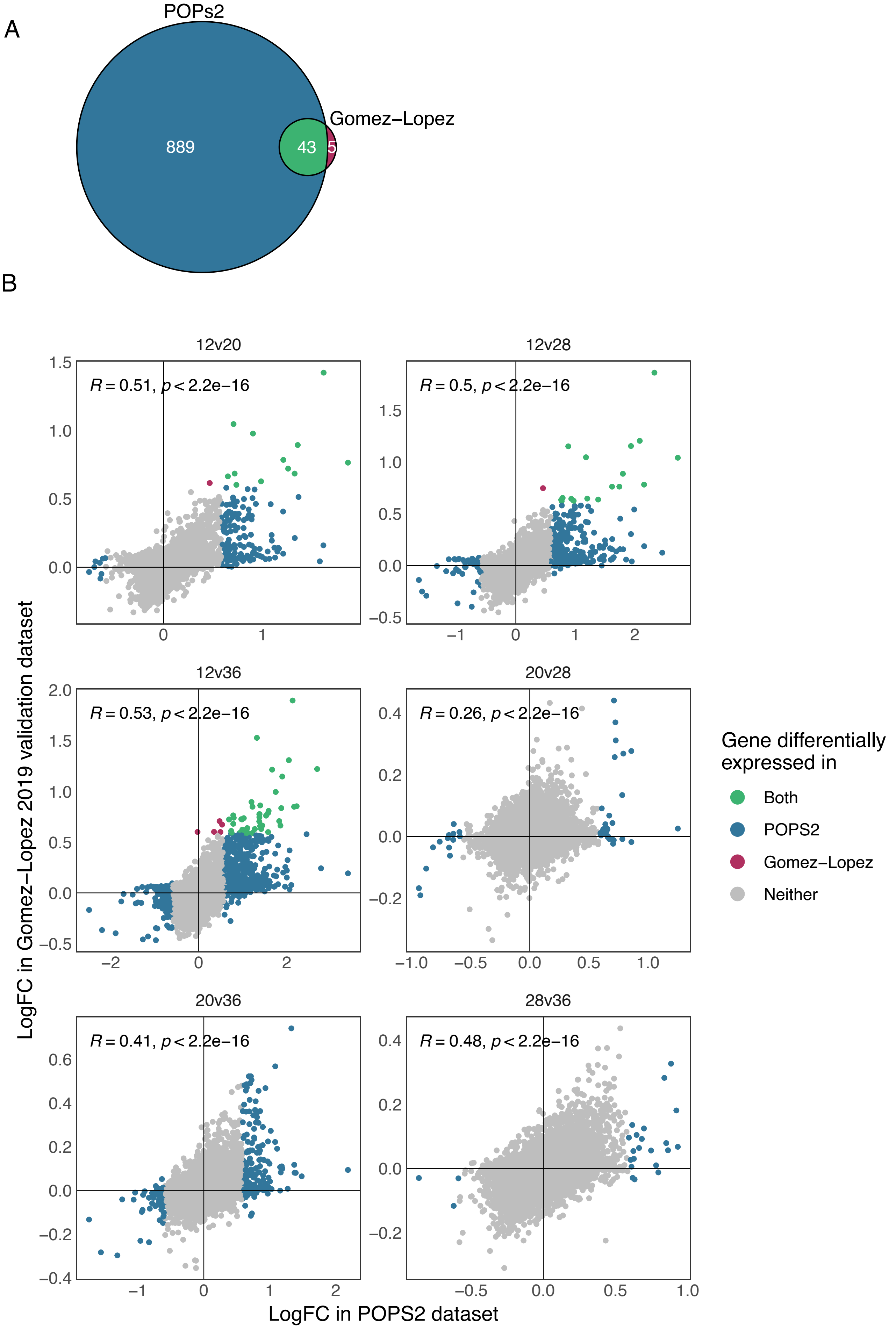


Supplementary Figure 9.
A. Venn diagram displaying the overlap between the number of (unique) differentially expressed genes in POPs2 and Gomez-Lopez et al. (2019). B. Scatterplot of logFC of genes in POPS2 vs. Gomez-Lopez et al. (2019) colored by dataset of significance.

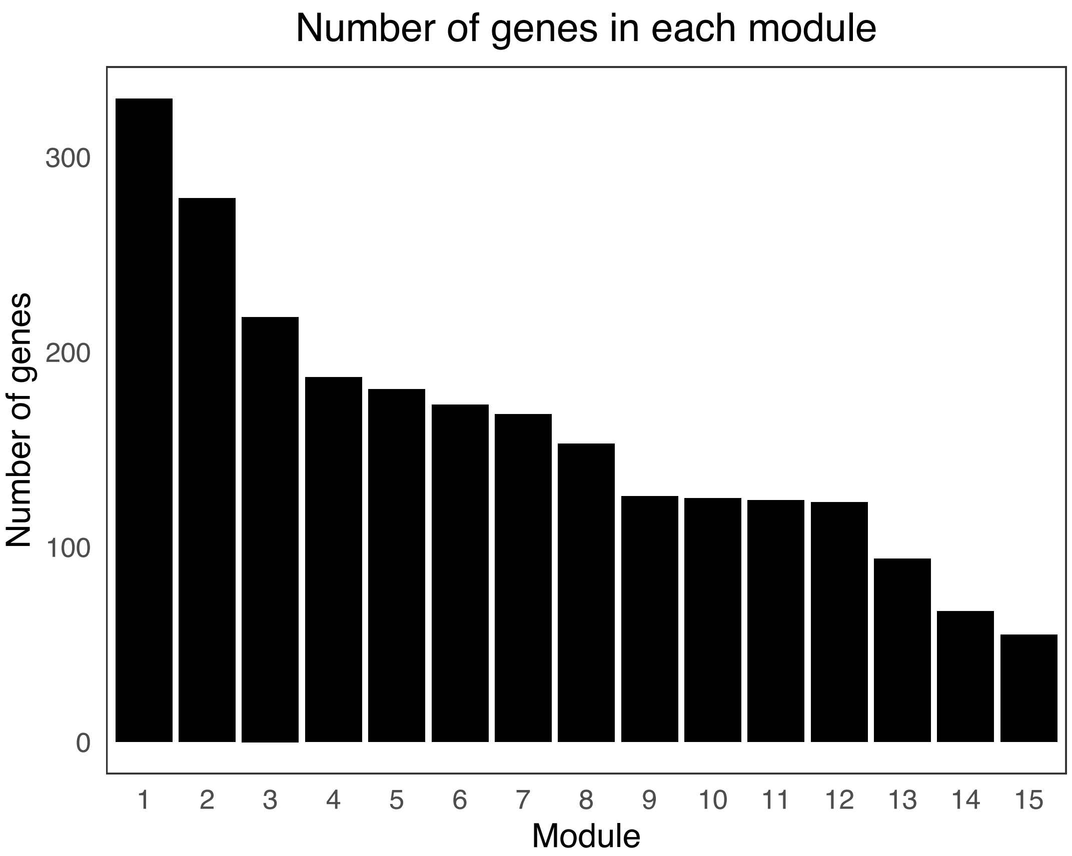


Supplementary Figure 10.
Barchart of number of genes in each WGCNA module.

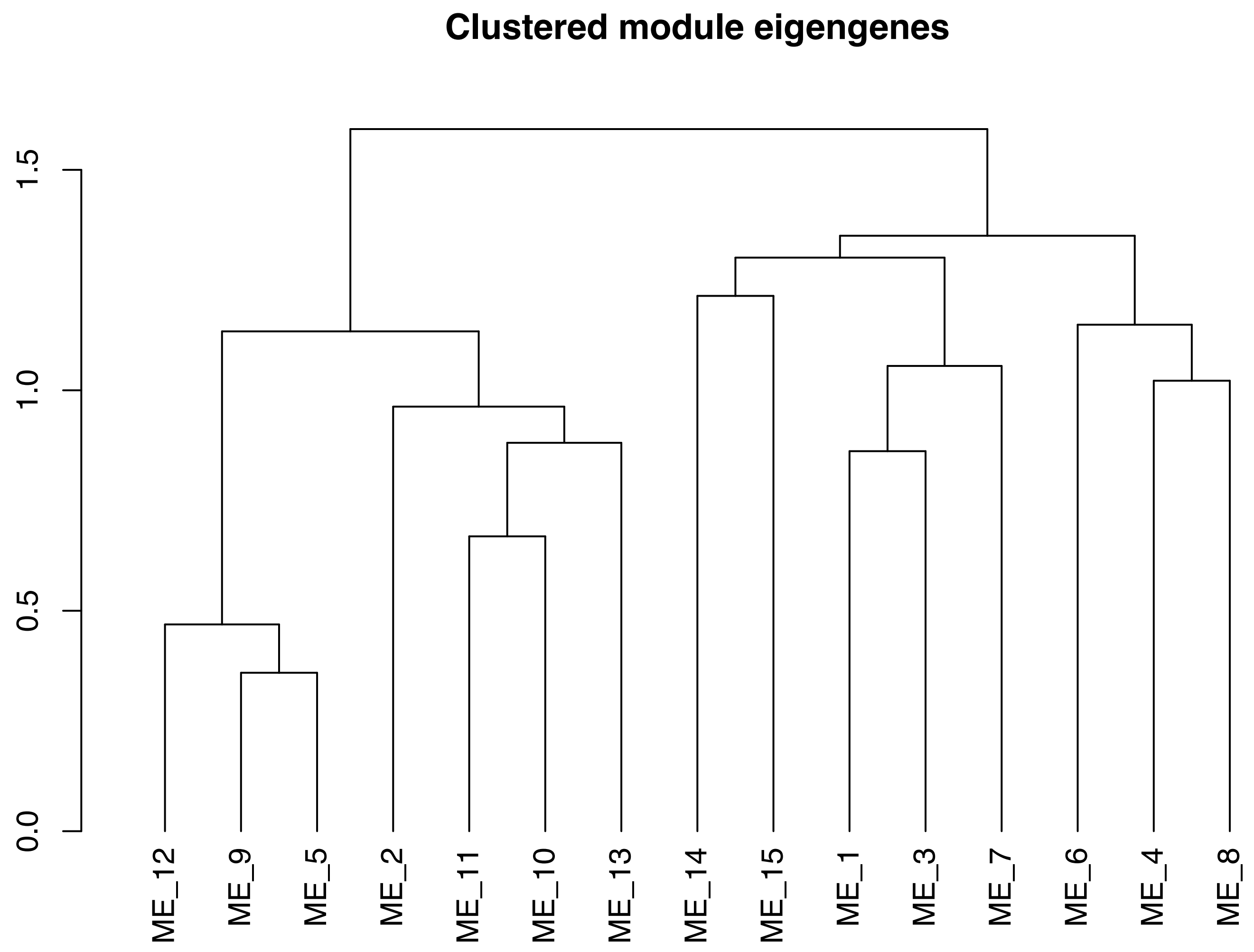


Supplementary Figure 11.
Modules clustered by eigengene values.

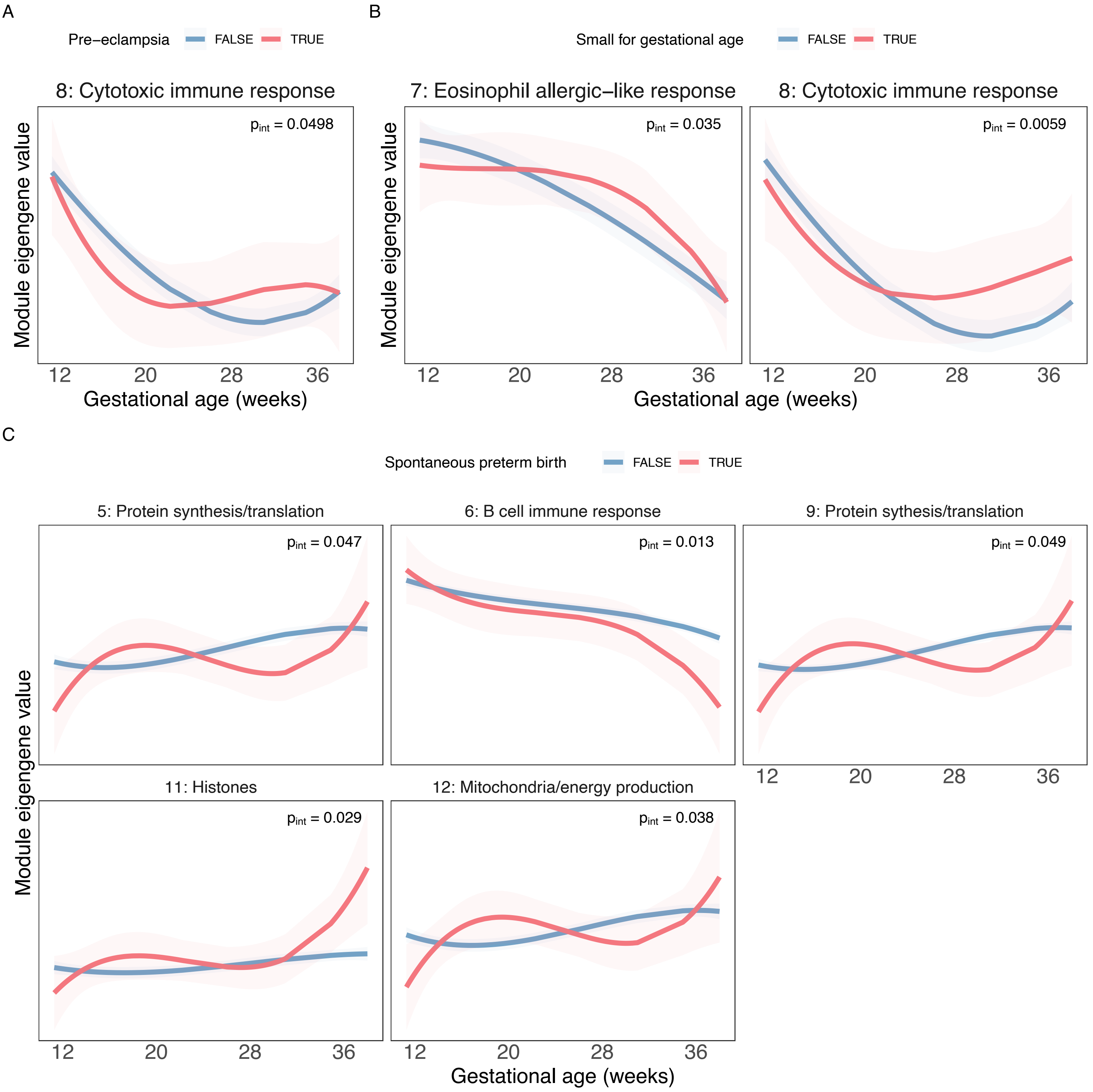


Supplementary Figure 12.
Modules with significantly different trajectories across clinical outcome groups, where curves represent predicted module eigengene values over time and shaded areas represent 95\% confidence interval of predicted values, colored by outcome group. A. Pre-eclampsia B. Small for gestational age. C. Spontaneous preterm birth.

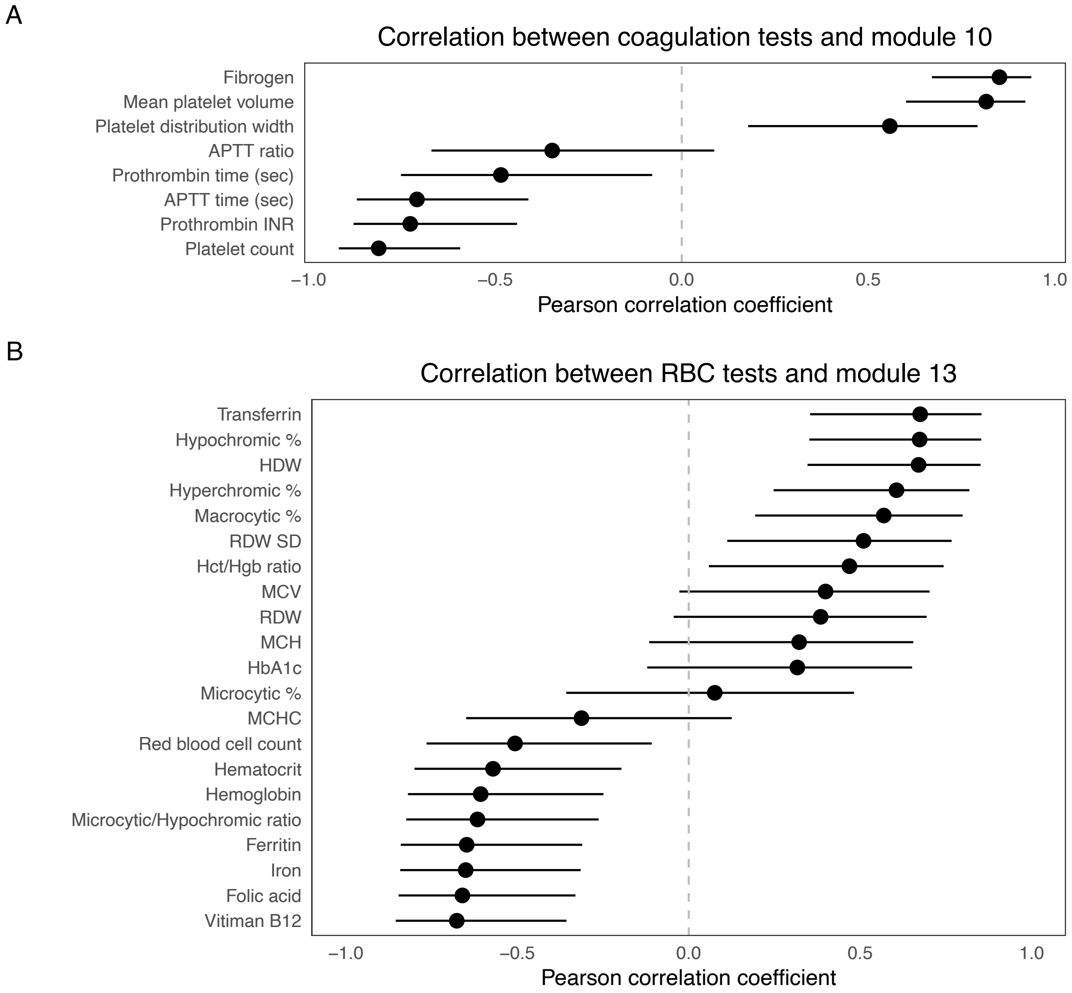


Supplementary Figure 13.
Pearson's correlation and 95% confidence interval for correlation between Module eigengene values in POPs2 and lab tests from Bar et al. (2025). A. Module 10 and coagulation-related tests. APTT=Activated Partial Thromboplastin Time, INR=International Normalized Ratio. B. Module 13 and red blood cell-related tests. HDW=Hemoglobin Distribution Width, RDW=Red Cell Distribution Width, SD=Standard Deviation, Hct=Hematocrit, Hgb=Hemoglobin, MCV=Mean Corpuscular Volume, MCH=Mean Corpuscular Hemoglobin, HbA1c=glycated hemoglobin, MCHC=Mean Corpuscular Hemoglobin Concentration.

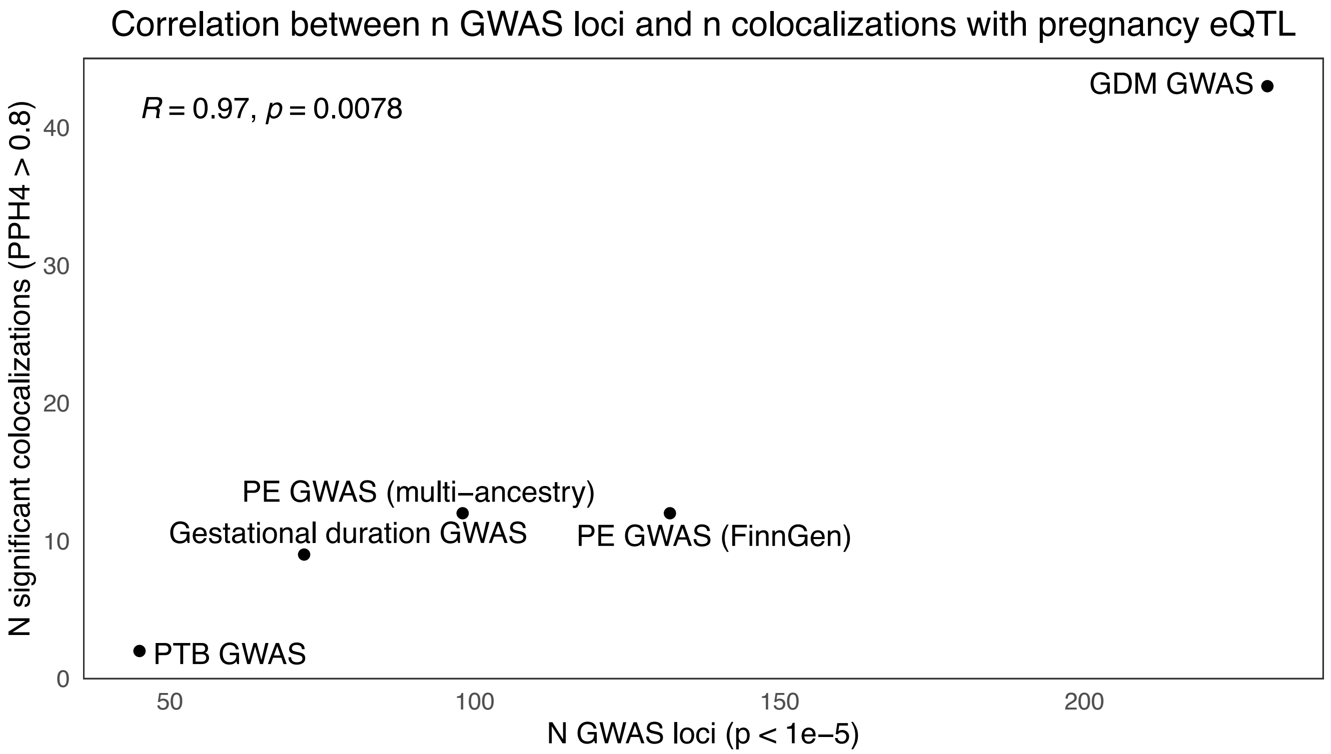


Supplementary Figure 14.
Pearson's correlation between number of GWAS loci and colocalizations in each GWAS dataset.

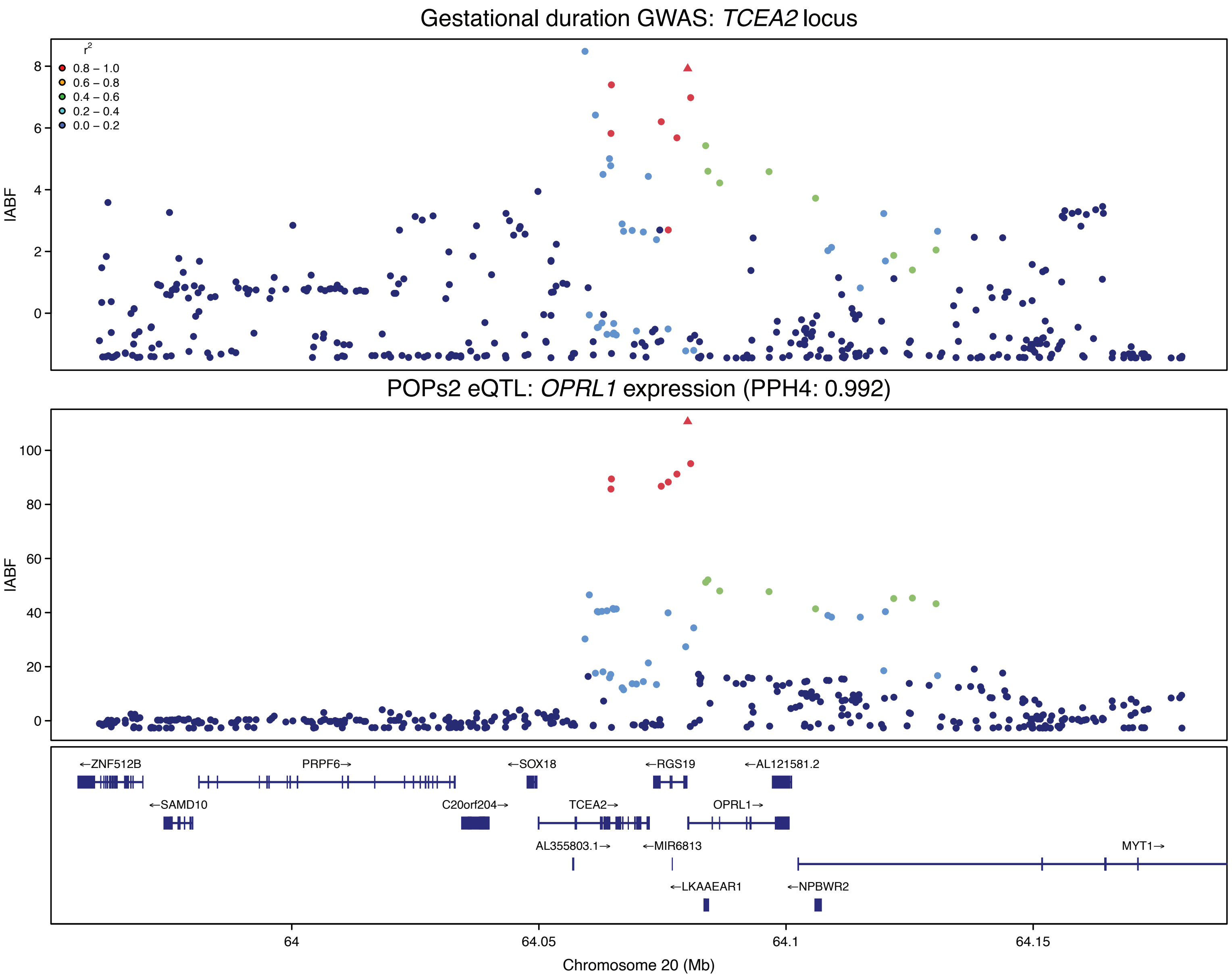


Supplementary Figure 15.

Regional association plots showing chromosome position vs. log approximate Bayes factor (lABF) from SuSiE. The lABF quantifies the strength of evidence that a variant is associated with the trait. The triangular point represents the joint lead SNP between GWAS and pregnancy eQTL signals and points are colored by LD with that SNP. PPH4 of colocalization between eQTL and GWAS signal noted in subtitle. Gestational duration GWAS locus and OPRL1 eQTL in POPs2.

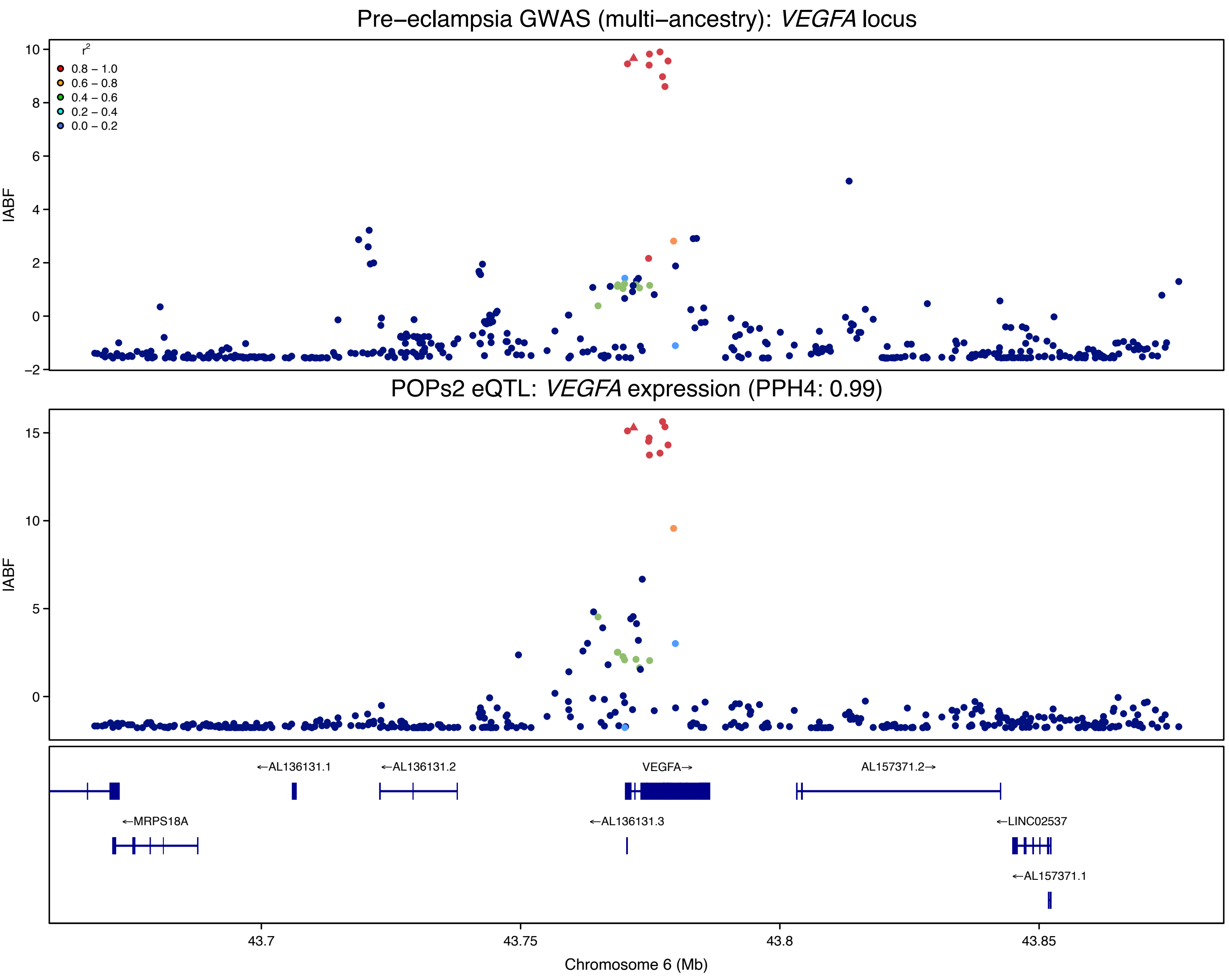


Supplementary Figure 16.
Regional association plots showing chromosome position vs. log approximate Bayes factor (lABF) from SuSiE. The lABF quantifies the strength of evidence that a variant is associated with the trait. The triangular point represents the joint lead SNP between GWAS and pregnancy eQTL signals and points are colored by LD with that SNP. PPH4 of colocalization between eQTL and GWAS signal noted in subtitle. Pre-eclampsia GWAS locus and VEGFA eQTL in POPs2.

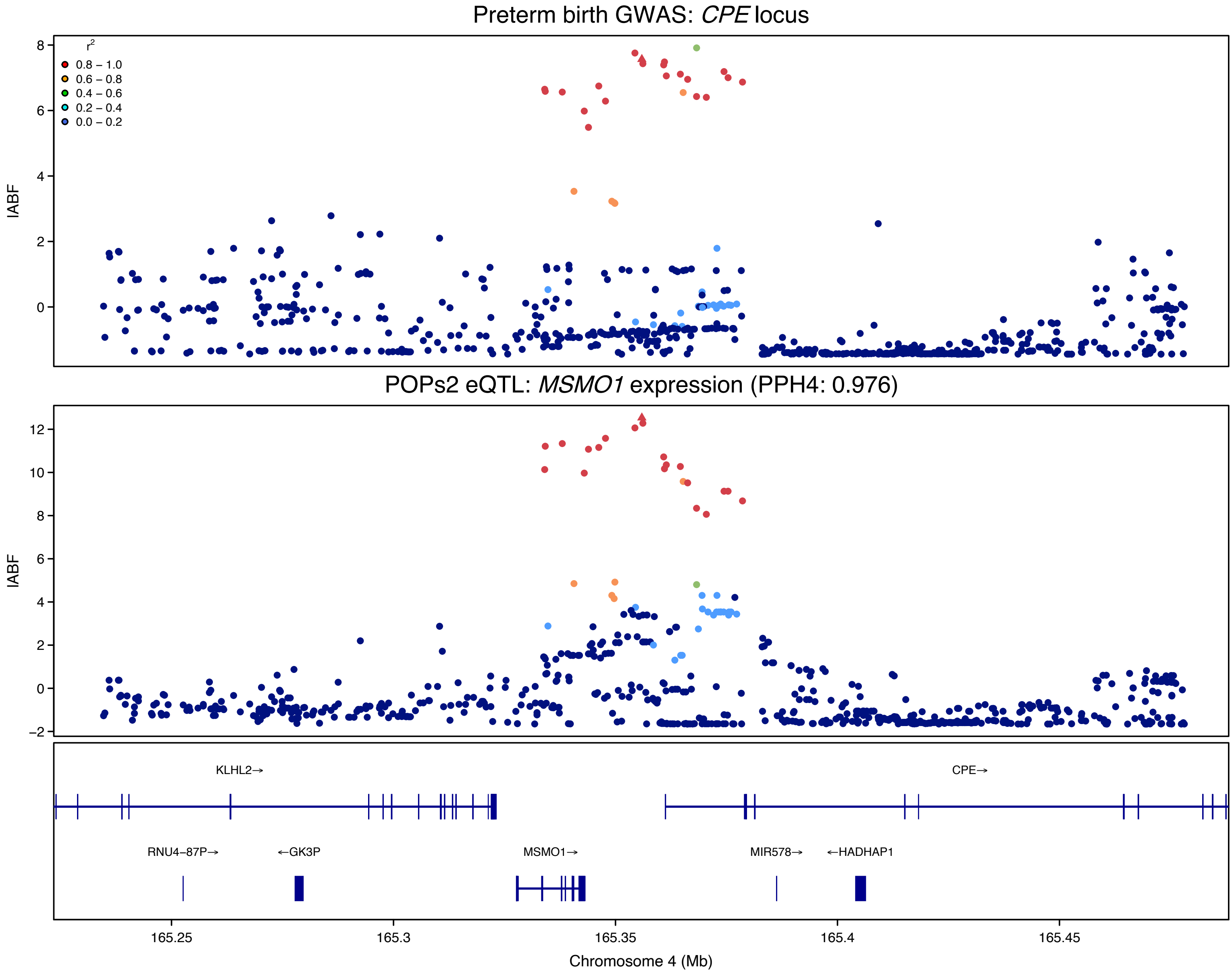


Supplementary Figure 17.
Regional association plots showing chromosome position vs. log approximate Bayes factor (lABF) from SuSiE. The lABF quantifies the strength of evidence that a variant is associated with the trait. The triangular point represents the joint lead SNP between GWAS and pregnancy eQTL signals and points are colored by LD with that SNP. PPH4 of colocalization between eQTL and GWAS signal noted in subtitle. Preterm birth GWAS locus and MSM01 eQTL in POPs2.

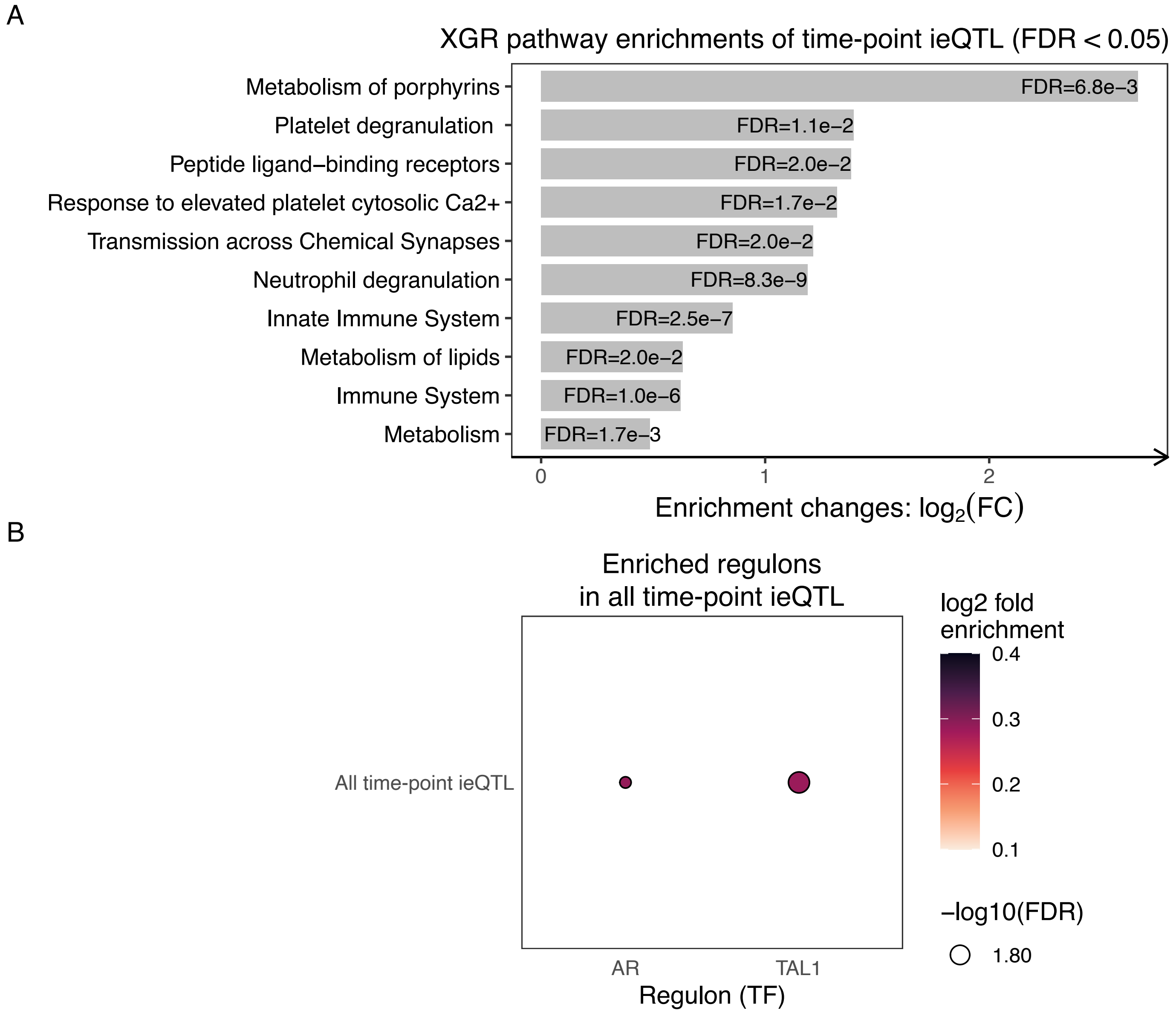


Supplementary Figure 18.
A. xEnrichBarplot showing concise enriched Reactome pathways among time interaction eGenes (FDR < 0.05). Rows represent Reactome pathways, log2 fold enrichment is displayed on the x-axis, and bars are annotated with corresponding FDR values. B. Dotplot of enriched regulons among time interaction eGenes across time-points. Dot size indicates significance, fill color indicates the fold enrichment or how strongly time interaction eGenes are enriched in each regulon. Only regulons with FDR < 0.05 shown.

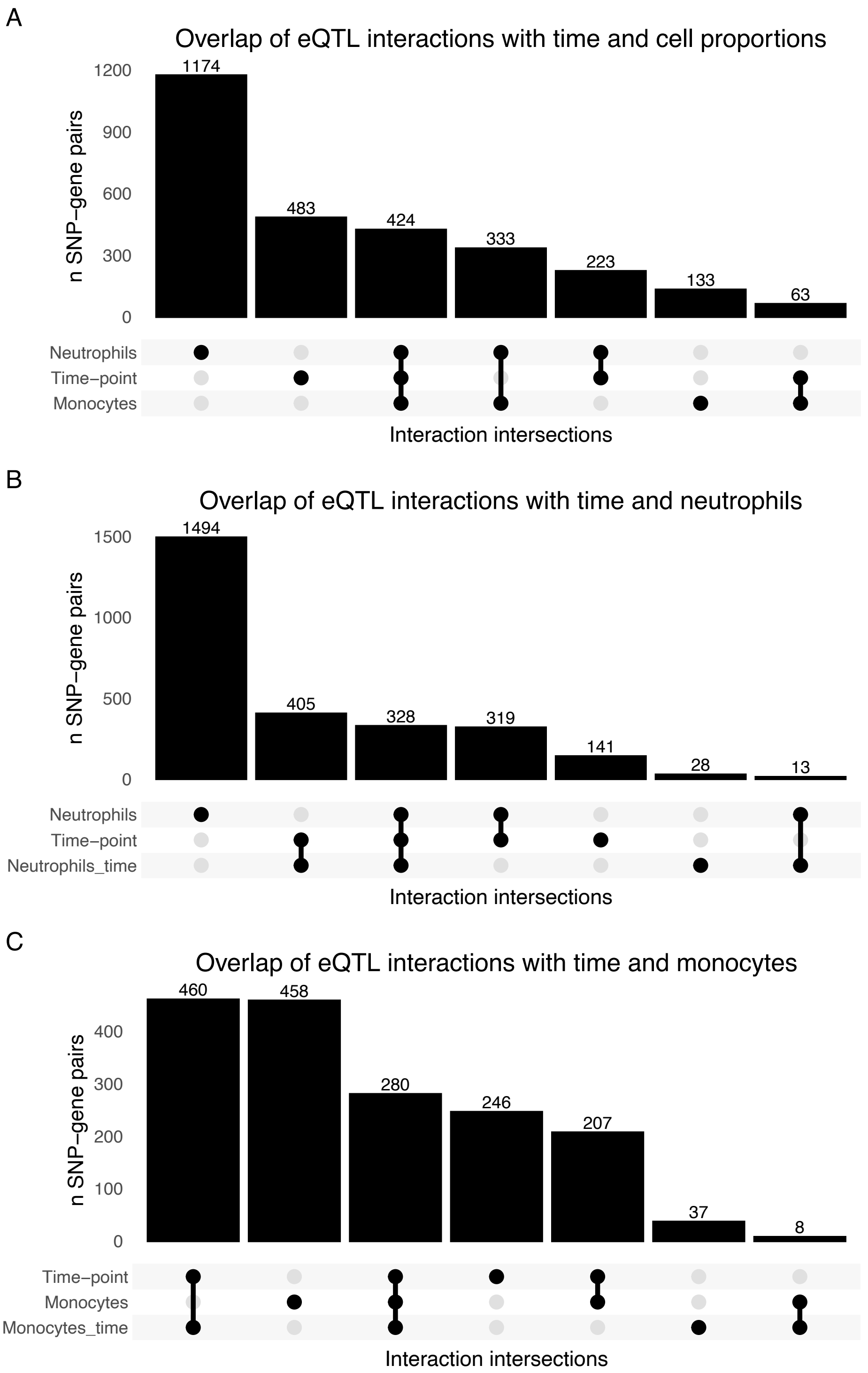


Supplementary Figure 19.
Upset plots showing overlap of eQTL with interaction effects. A. Overlap of eQTL with time-point, neutrophil, and monocyte proportion interactions. B. Overlap of time-point, neutrophil, and neutrophil-adjusted time-point eQTL interactions. C. Overlap of time-point, monocyte, and monocyte-adjusted time-point eQTL interactions.

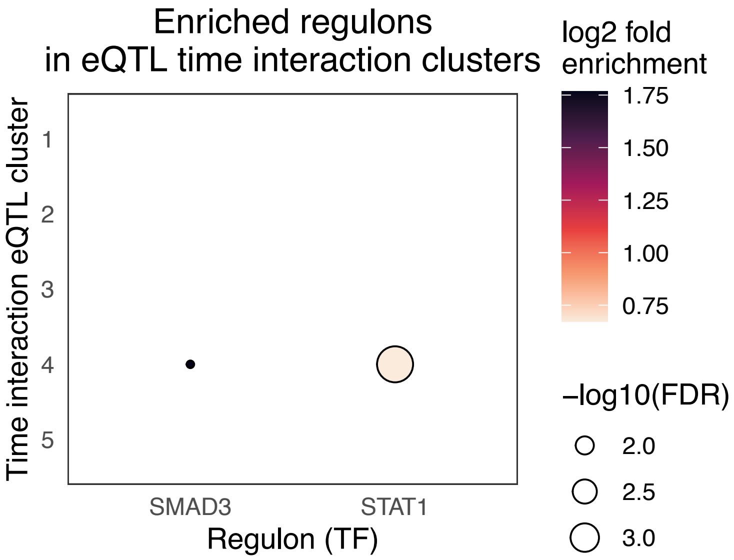


Supplementary Figure 20.
Dotplot of enriched regulons among time interaction cluster eGenes across time-points. Dot size indicates significance, fill color indicates the fold enrichment or how strongly time interaction cluster eGenes are enriched in each regulon. Only regulons with FDR < 0.05 shown.

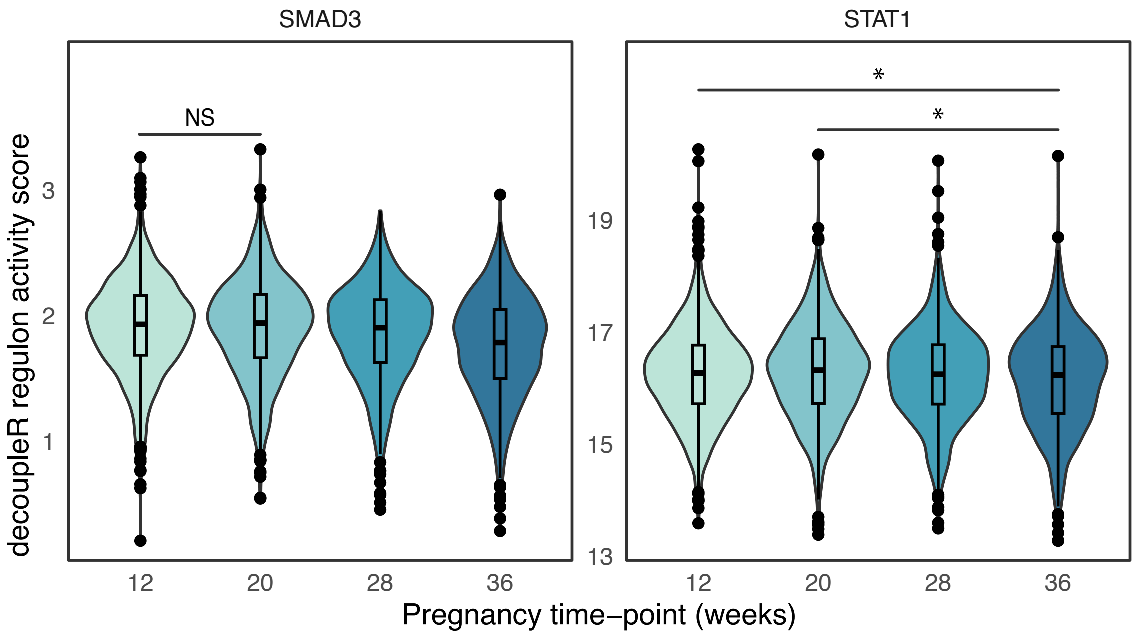


Supplementary Figure 21.
Violin plots of regulon activity scores by time-point showing change in activity over time in SMAD3 and STAT1. In SMAD3, all pairs of time-points have significant ($p < 0.05$) changes in regulon activity but the pair marked NS. In STAT1, only the time-point pairs denoted with * have significant ($p < 0.05$) changes in regulon activity.

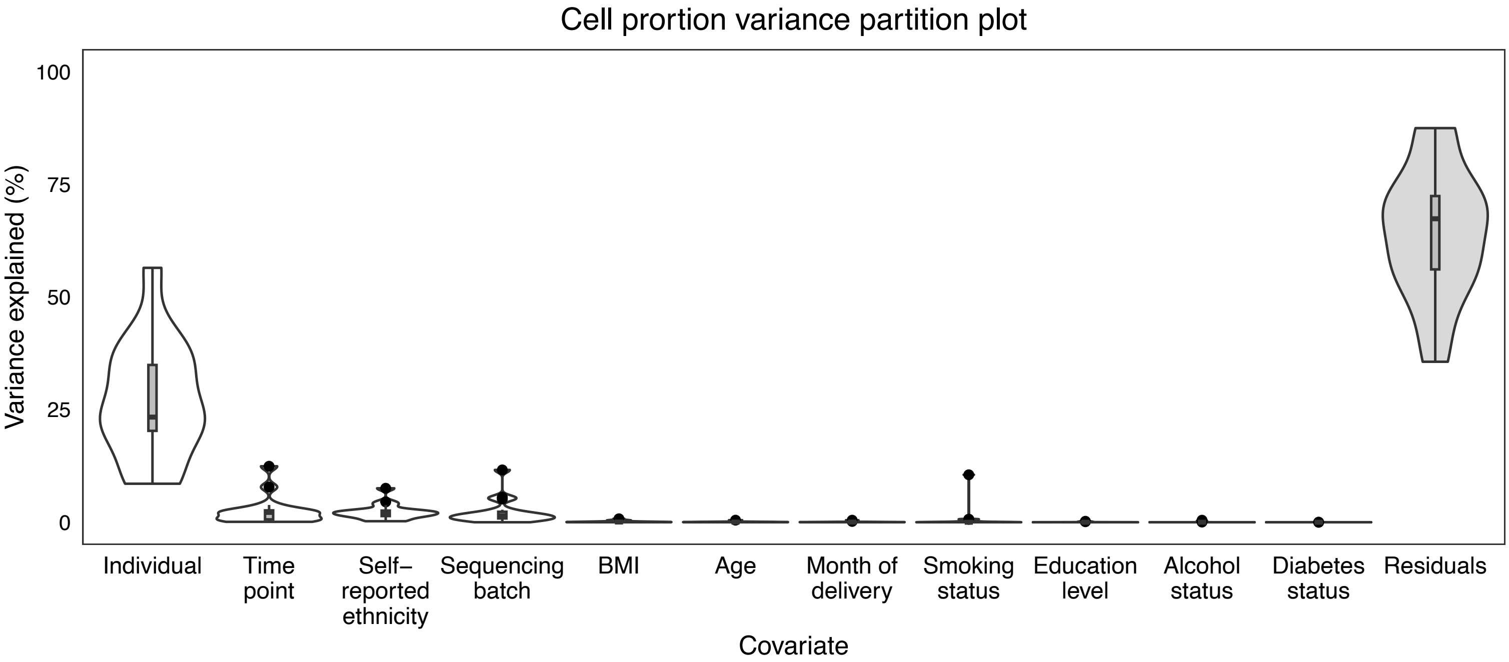


Supplementary Figure 22.
Proportion of imputed cell proportion variance explained (y) by clinical and technical covariates (x).

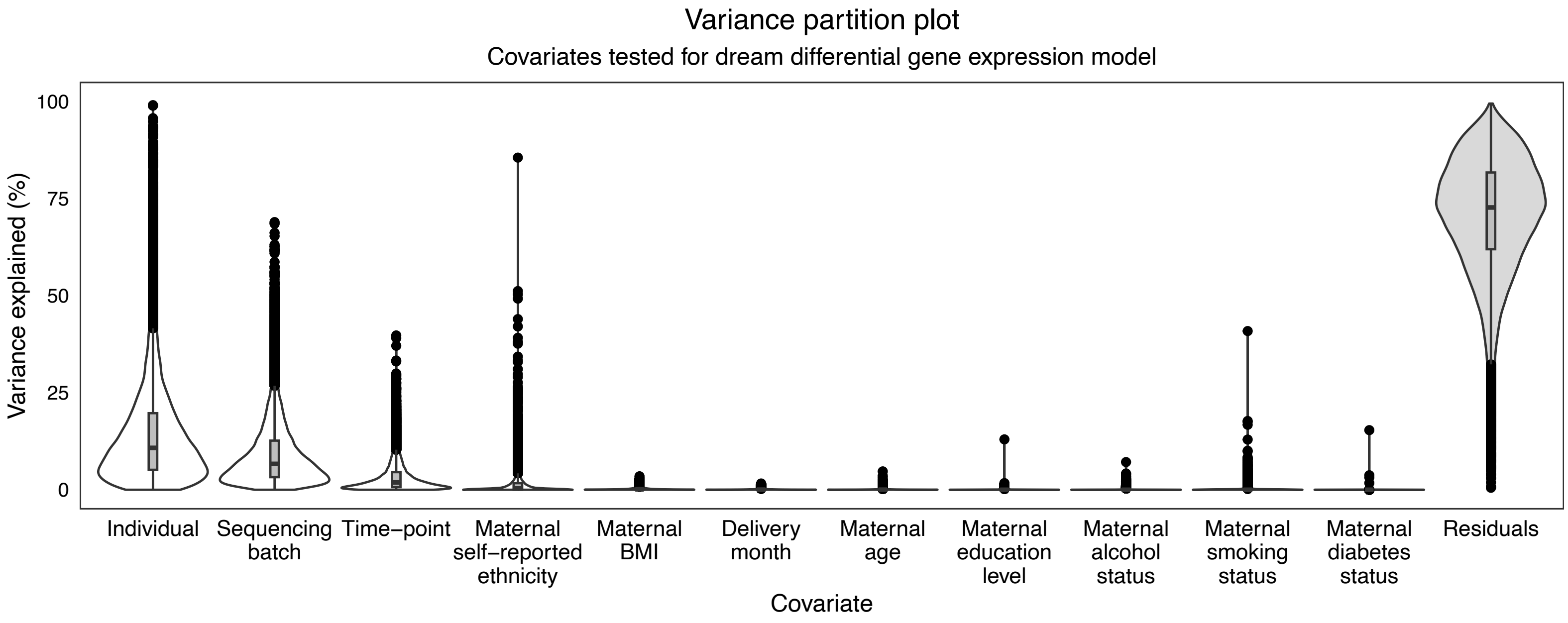


Supplementary Figure 23.
Proportion of gene expression variance explained (y) by clinical and technical covariates evaluated for inclusion in the dream model (x).

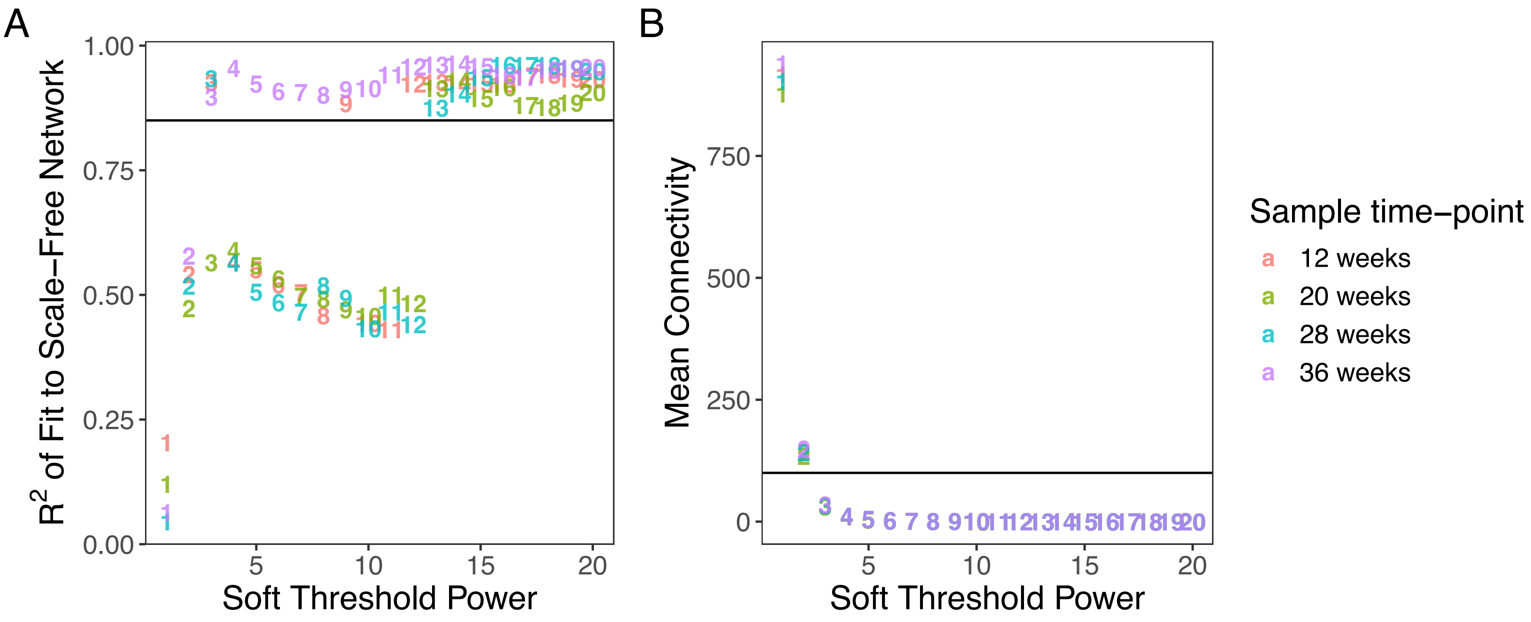


Supplementary Figure 24.

A. Soft threshold power vs. $R^2$ of fit to scale-free network colored by dataset time-point with horizontal line at y=0.85, WGCNA's default cutoff. B. Soft threshold power vs mean connectivity colored by dataset time-point with horizontal line at y=100, WGCNA's recommended max value for mean connectivity.

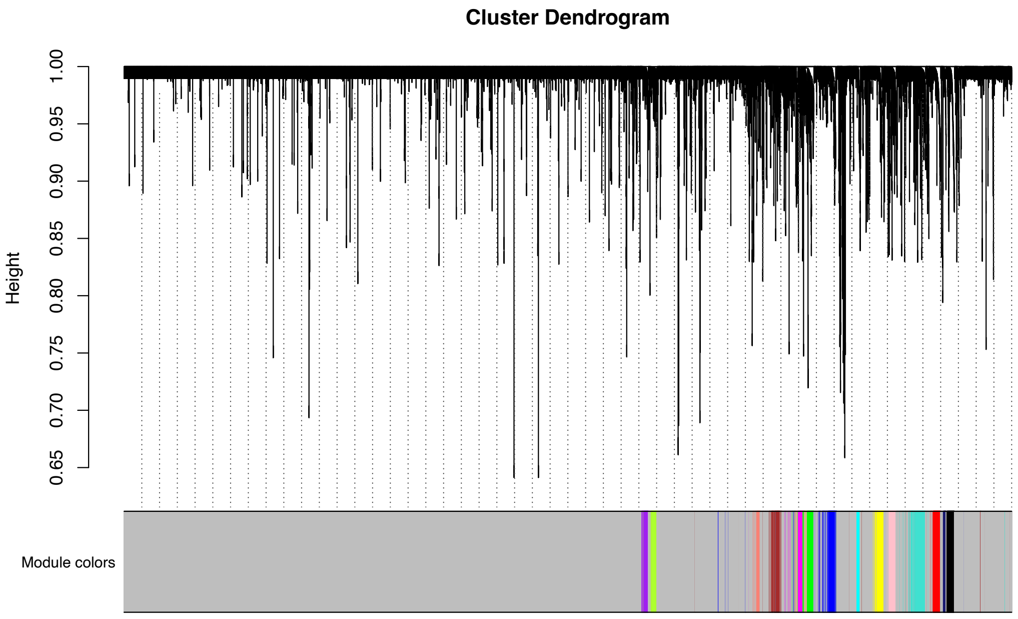


Supplementary Figure 25.
A.  Soft threshold power vs. $R^2$ of fit to scale-free network colored by dataset time-point with horizontal line at y=0.85, WGCNA's default cutoff. B. Soft threshold power vs mean connectivity colored by dataset time-point with horizontal line at y=100, WGCNA's recommended max value for mean connectivity.

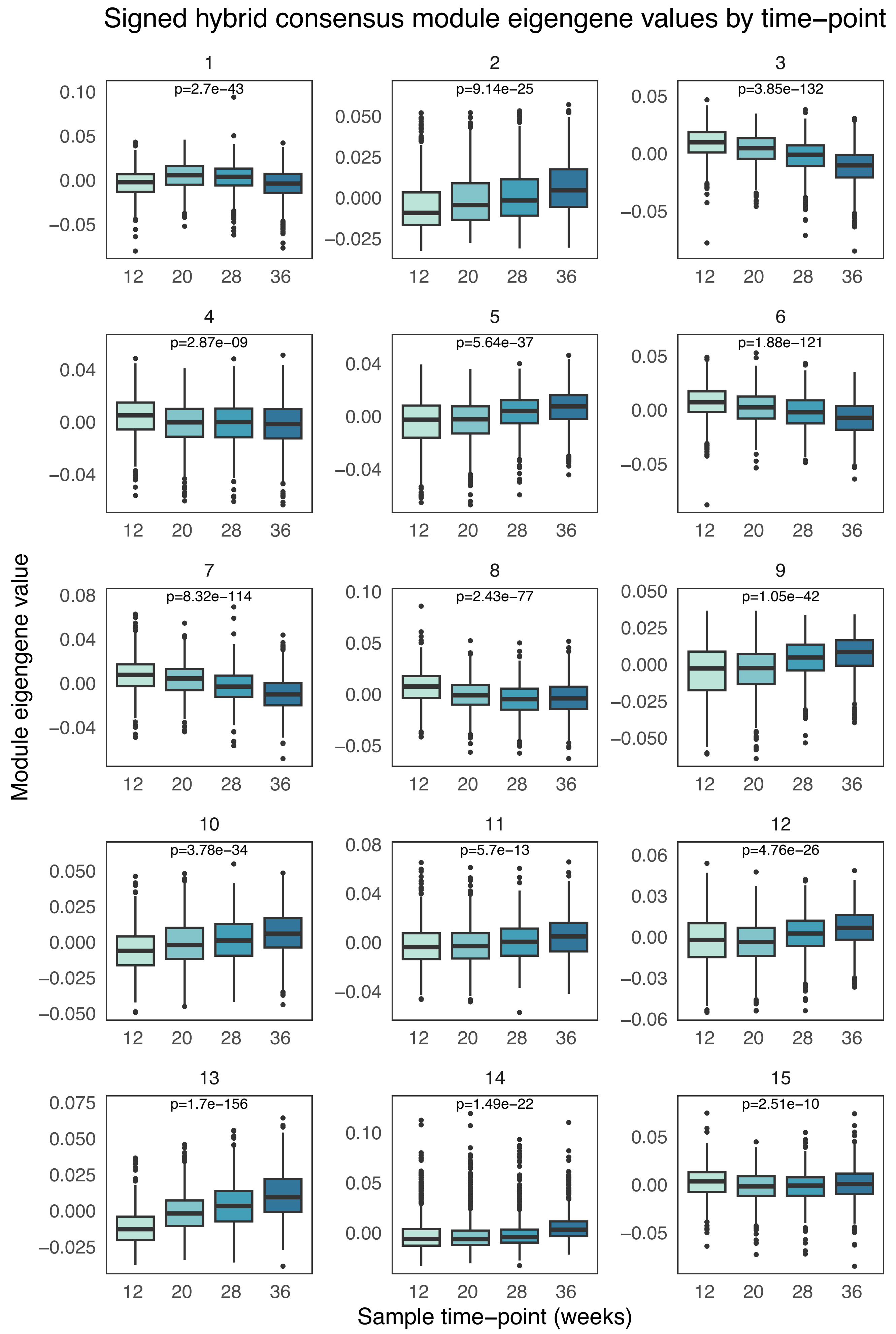


Supplementary Figure 26.
Boxplot of eigengene values by discrete time-point for each module. Annotated with adjusted p-value from repeated measures ANOVA test for association between module eigengene value and time-point.

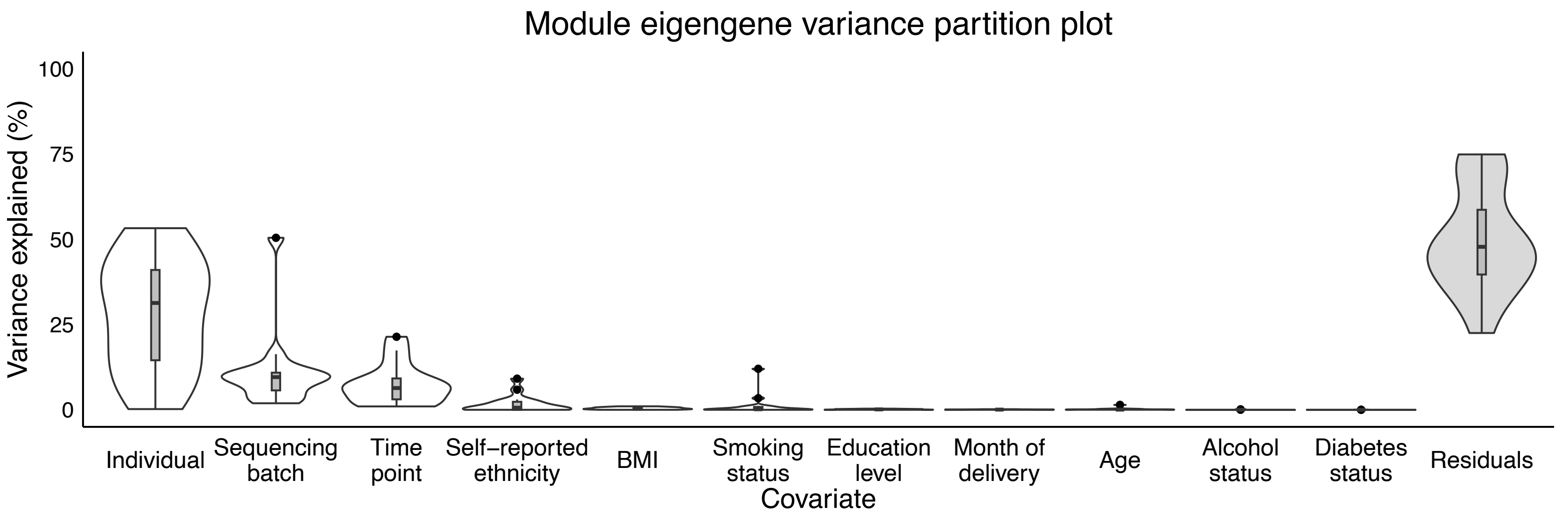


Supplementary Figure 27.
Proportion of module eigengene variance explained (y) by clinical and technical covariates (x).

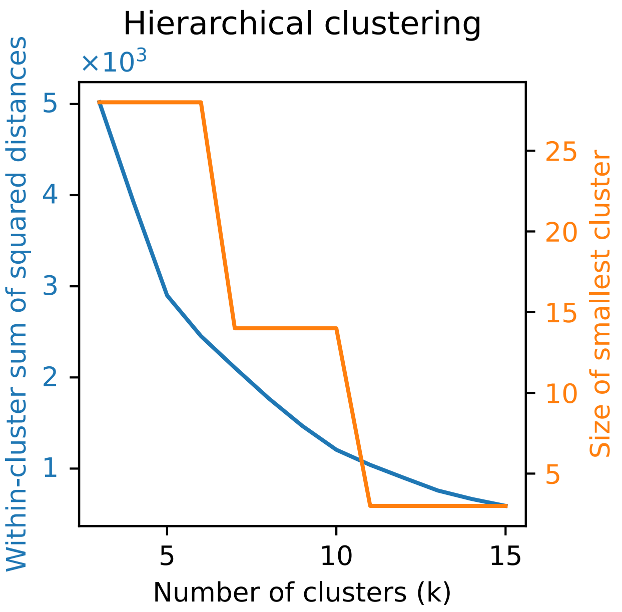


Supplementary Figure 28.
Variance clustering metrics: Within-cluster sums of squared distances and size of smallest cluster for k = 3-15.

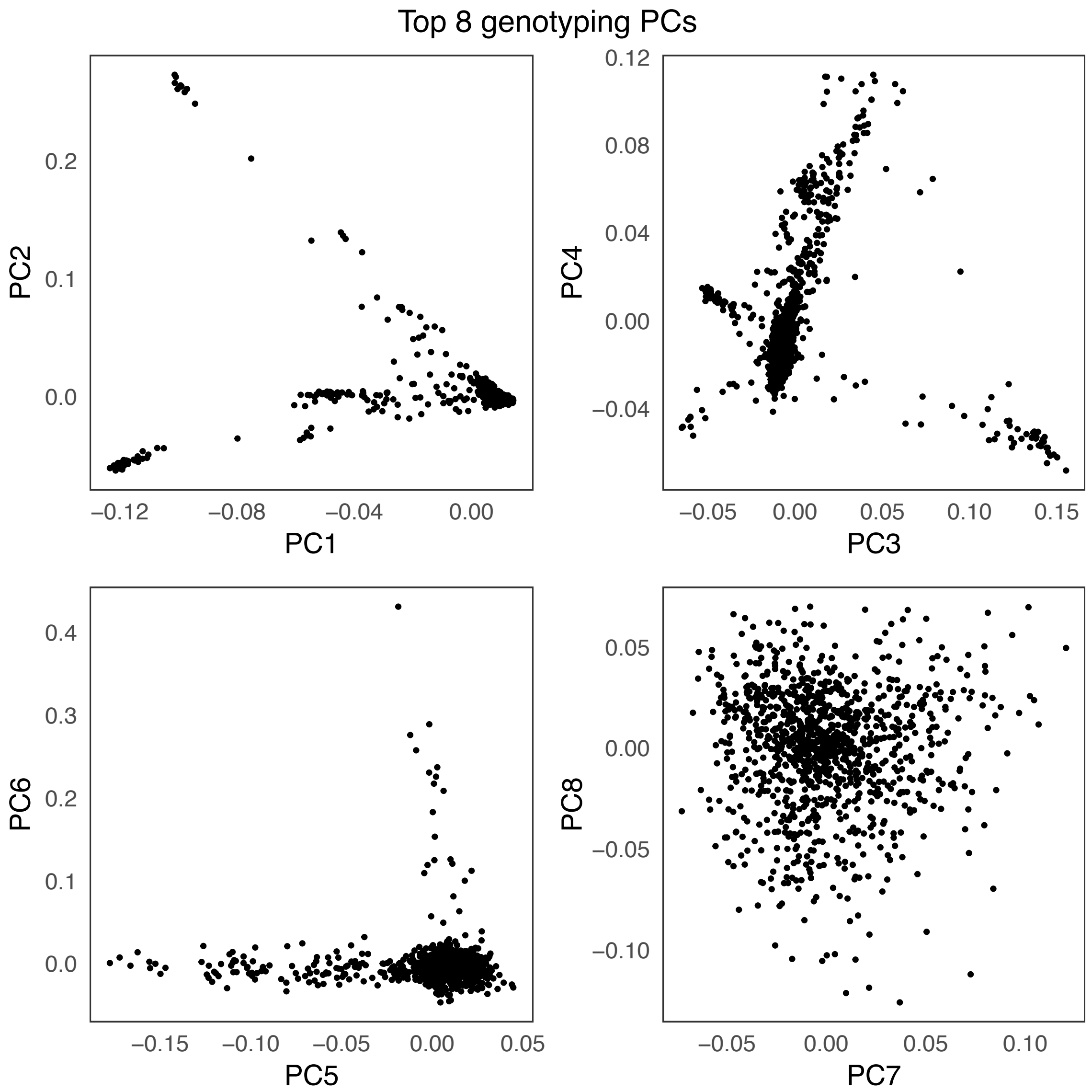


Supplementary Figure 29.
Top 8 genotyping PCs.

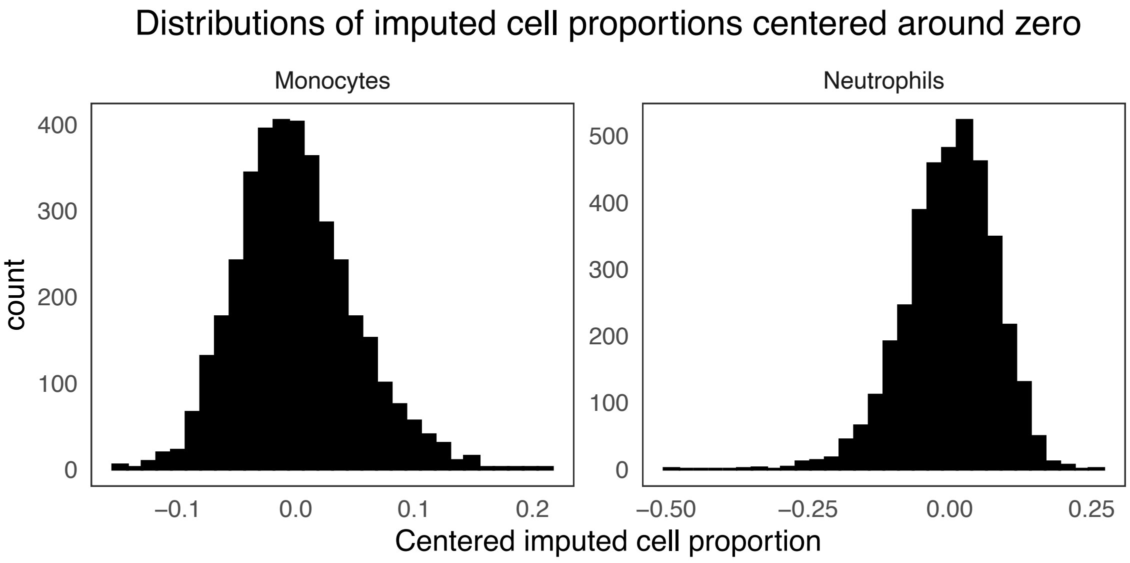


Supplementary Figure 30.
Histogram of imputed neutrophil and monocyte proportions centered around 0.

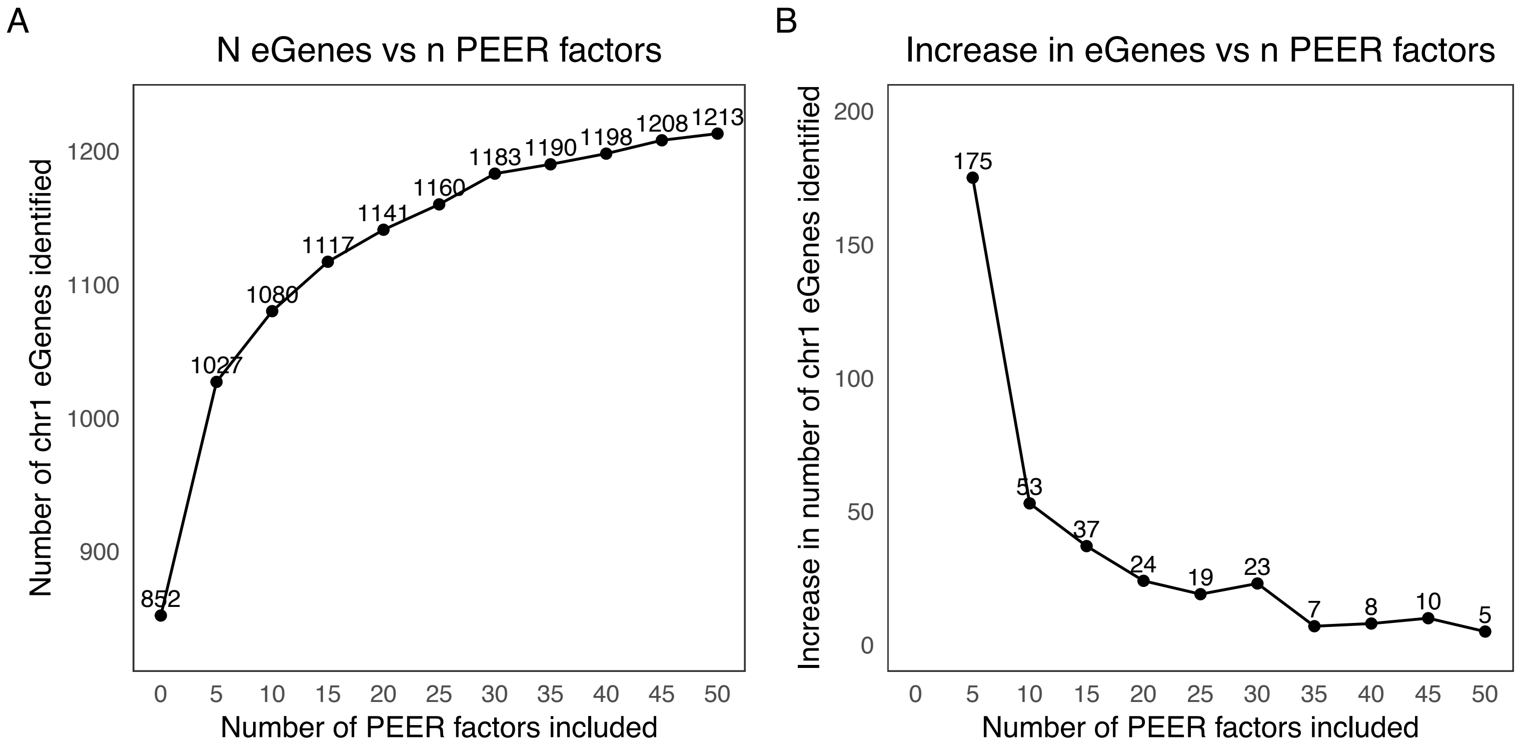


Supplementary Figure 31.
A. Number of eGenes identified vs number of PEER factors included in eQTL model of chromosome 1. B. Increase in number of eGenes identified vs number of PEER factors included in eQTL model of chromosome 1.

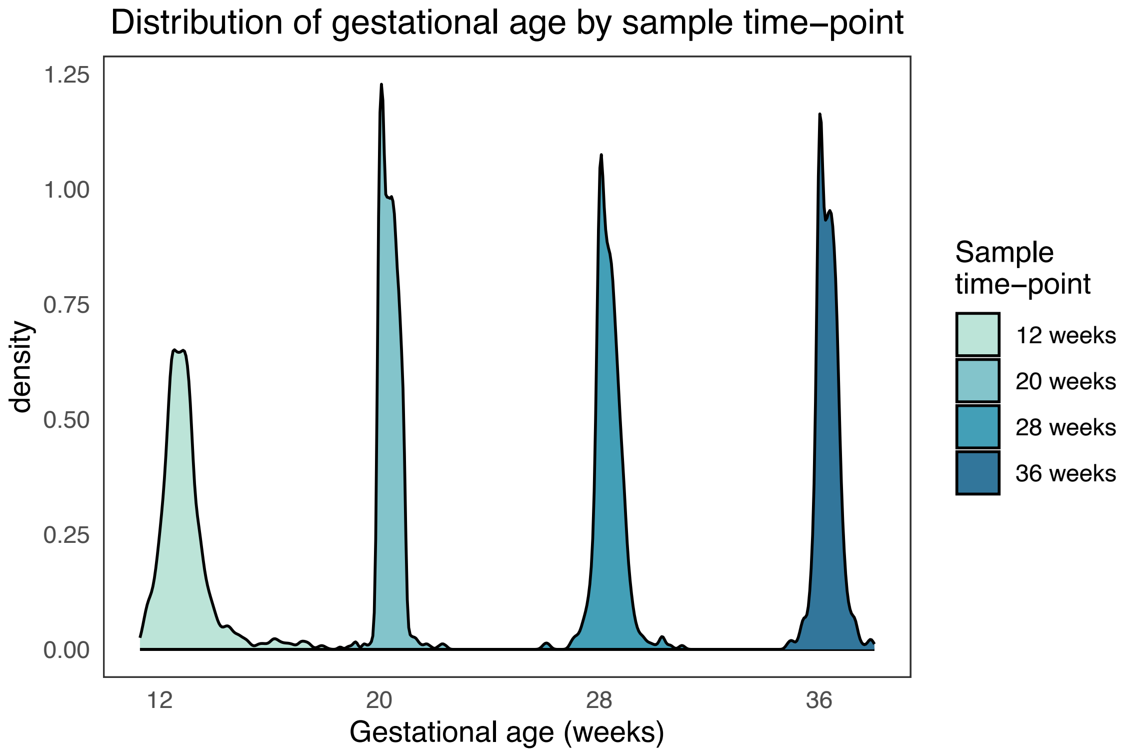


Supplementary Figure 32.
Distribution of gestational age by sample time-point in the full RNA-seq cohort.

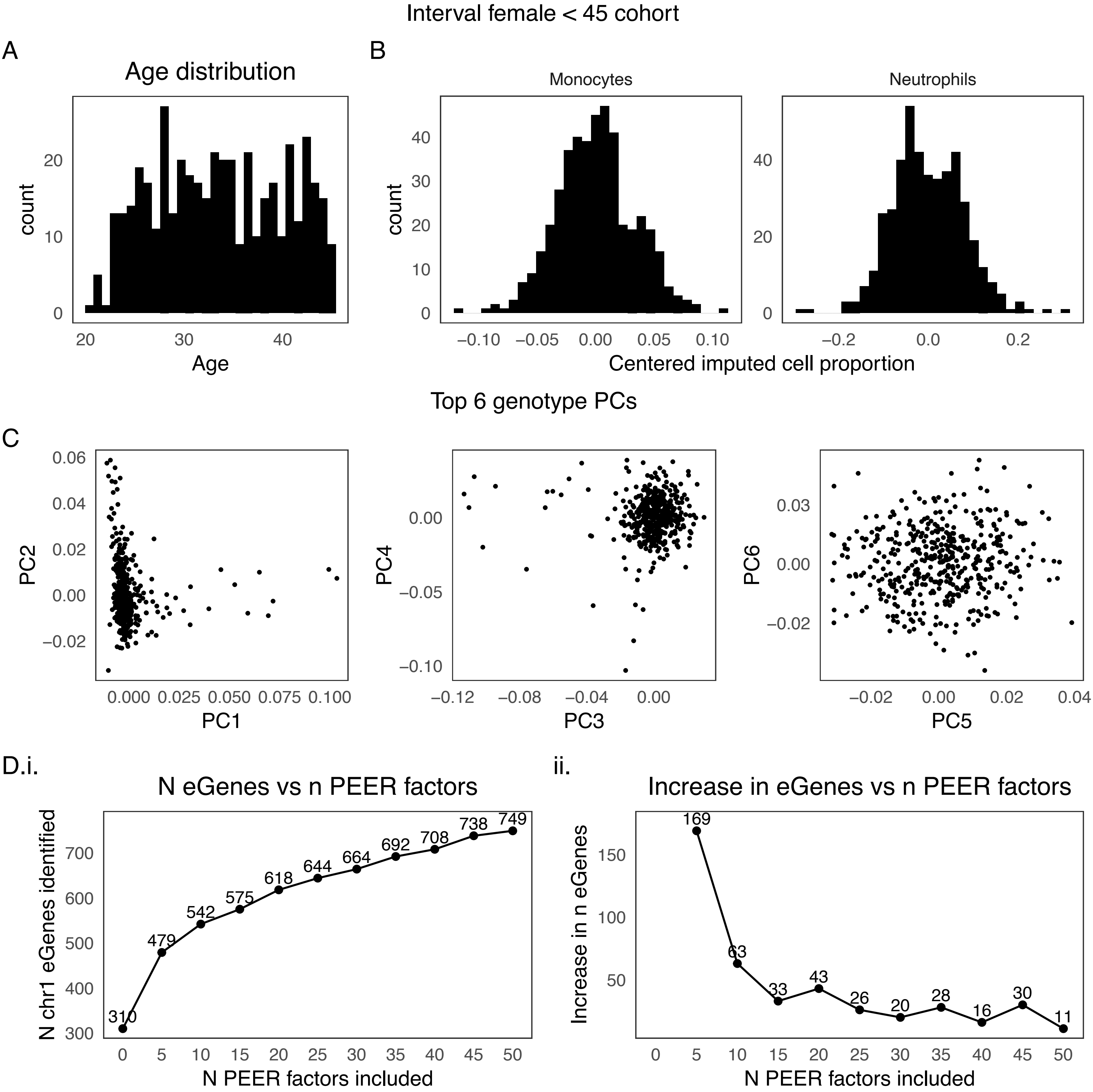


Supplementary Figure 33.

A. Age distribution in INTERVAL females <45 years of age. B. Histogram of imputed neutrophil and monocyte proportions centered around 0. C. Top six genotyping PCs. D.i. Number of eGenes identified vs number of PEER factors included in eQTL model of chromosome 1. D.ii. Increase in number of eGenes identified vs number of PEER factors included in eQTL model of chromosome 1.

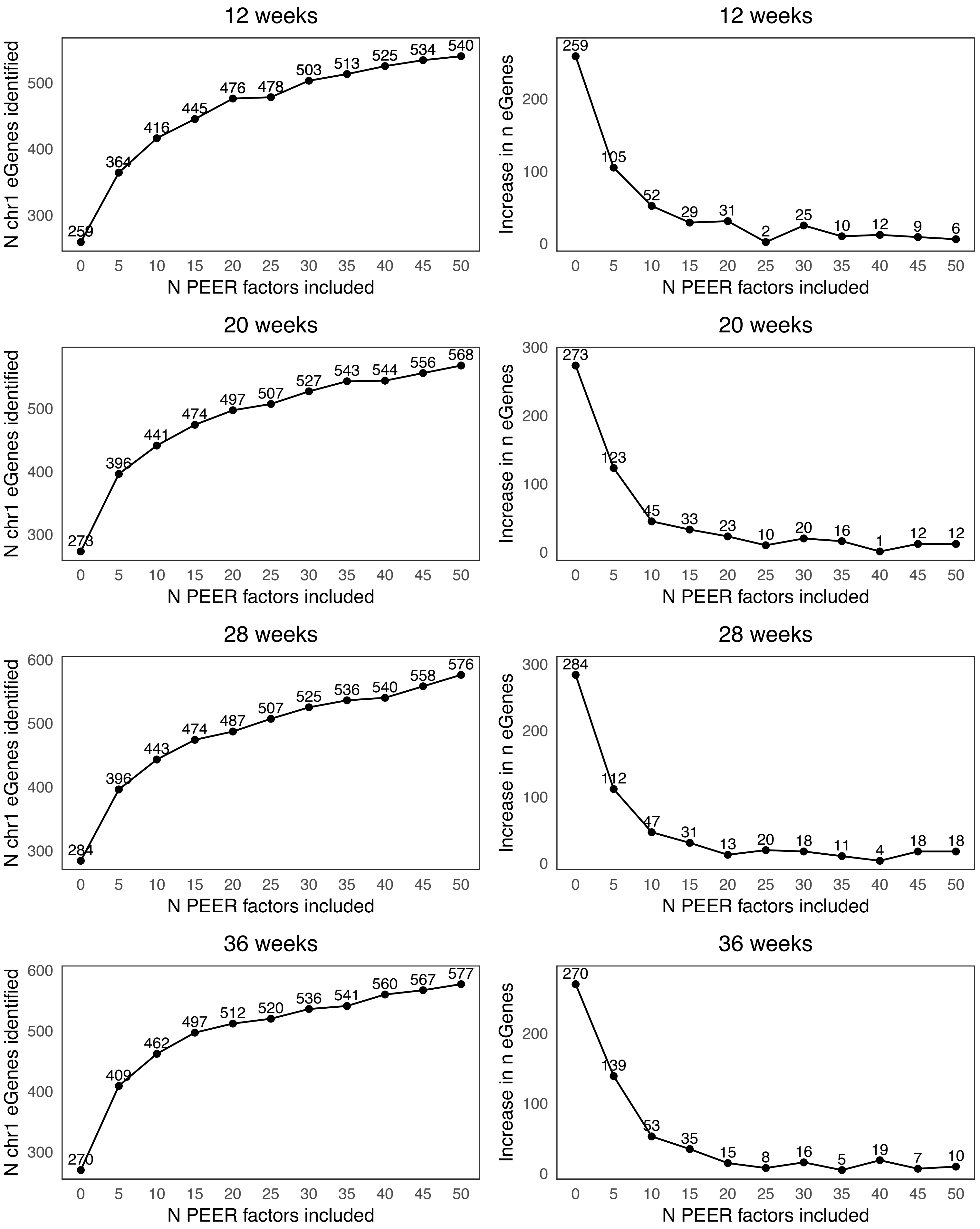


Supplementary Figure 34.
Number of eGenes identified vs number of PEER factors included in eQTL model of chromosome 1 and increase in number of eGenes identified vs number of PEER factors included in eQTL model of chromosome 1 in each time-point subset.

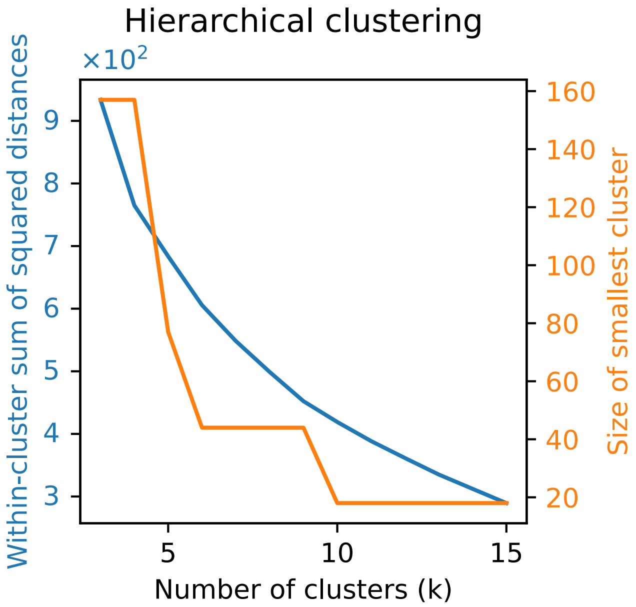


Supplementary Figure 35.
Time interaction eQTL clustering metrics: Within-cluster sums of squared distances and size of smallest cluster for k = 3-15.
